# MRSL: A phenome-wide causal discovery algorithm based on GWAS summary data

**DOI:** 10.1101/2022.06.29.22277051

**Authors:** Lei Hou, Zhi Geng, Xu Shi, Chuan Wang, Hongkai Li, Fuzhong Xue

## Abstract

Causal discovery is a powerful tool to disclose underlying structures by analyzing purely observational data. Genetic variants can provide useful complementary information for structure learning. Here, we propose a novel algorithm MRSL (Mendelian Randomization (MR)-based Structure Learning algorithm), which combines the graph theory with univariable and multivariable MR to learn the true structure using only GWAS summary statistics. Specifically, MRSL also utilizes topological sorting to improve the precision of structure learning and provides three adjusting categories for multivariable MR. Results of simulation reveal that MRSL has up to two-fold higher F1 score than other eight competitive methods. Additionally, the computing time of MRSL is 100 times faster than other methods. Furthermore, we apply MRSL to 26 biomarkers and 44 ICD10-defined diseases from UK Biobank. The results cover most of expected causal links which have biological interpretations and several new links supported by clinical case reports or previous observational literatures.

## Introduction

Causal discovery aims to infer causal structure by analyzing purely observational data [1-2]. It can be widely applied in the social and natural sciences, and it is a powerful tool for discovering biological networks [3-4] and disease diagnostic purposes [5-6], etc. Graphical models reveal the generating process of the observed data and they can be identified under three assumptions [1-2, 7-8]: (1) the causal Markov condition, (2) the causal faithfulness assumption and (3) the causal sufficiency assumption. The causal Markov condition means that all nodes are independent of their non-descendants when conditionally on their parents. The causal faithfulness assumption requires all conditional independences in true underlying distribution 𝕡 are represented in the graph and are invariant to changes in parameterization. The causal sufficiency assumption states that any pair of nodes in the graph has no common external cause, and it implies there is no unobserved confounding variable. Various algorithms have been developed and can learn causal structures from purely or mostly observational data [9]. Constraint-based methods start with a fully connected graph and carry out a series of marginal and conditional independence tests to decide which edges should be removed, such as PC [10] and Markov blanket detection algorithm (e.g. Grow-Shrink [11] and Incremental Association algorithm [12-13]), etc. The outputs of such algorithms are equivalence classes. Score-based methods find the most plausible Directed Acyclic Graph (DAG) by optimizing a properly defined score function, such as the hill climbing (HC) greedy search algorithm [14], etc. The hybrid algorithms have become widely used because they combine the advantages of the constraint-based and score-based algorithms [15]. One popular strategy is to use constraint-based algorithms to determine an initial network structure and then to use score-based algorithms to find the highest-scoring network structure. For example, Max-Min Hill-Climbing (MMHC) [16] and Restricted Maximization algorithm [17], etc. All of these methods require the causal sufficiency assumption and require that the input data sets must be individual data. Recently several algorithms have been developed to learn the structure with latent variables, that is, relax the causal sufficiency assumption, such as Fast Causal Inference (FCI) algorithm [18-19] and Greedy FCI [20], etc. However, they output Partial Ancestral Graphs (PAG), but not complete structure information. All of the above algorithms are for structural learning but not for parameter learning

Utilization of genetic variants which are robustly associated with a risk factor provides a directional causal anchor for causal discovery. Thus the combination of Mendelian Randomization (MR) and causal discovery becomes popular. MR methods use genetic variants as instrumental variables (IVs) to infer the causal relationship from the exposure to the outcome [21]. These IVs can be used to remove confounding biases and to avoid reversed causal relationships. A valid IV must satisfy three assumptions: (1) Relevance – IV is robustly associated with the exposure; (2) Exchangeability – IV is not associated with any confounder of the exposure–outcome relationship; (3) Exclusion restriction – IV is independent of the outcome conditional on the exposure and all confounders of the exposure-outcome relationship. Richard et al. [22-23] concluded that Bayesian network (BN) incorporating genetic anchors is a useful complementary method to conventional MR for exploring causal relationships in complex omic data sets. A novel machine learning algorithm named MRPC incorporates the Principle of Mendelian randomization (PMR) in the PC algorithm, to learn causal graphs [24]. Nevertheless the algorithm also requires causal sufficiency assumption, i.e. no unobserved confounders among all the variables. Actually, this method only uses the information of genetic variants but not the idea of MR. Afterwards, David et al. [25] presented a pipeline, named causal Graphical Analysis Using Genetics (cGAUGE), using IV filters with provable properties to perform univariable MR (UVMR) [26-27] then to obtained a causal graph. This algorithm allows the unobserved confounders among all the variables and requires individual genetic and phenotypic data. A flexible two stage procedure called bidirectional mediated Mendelian randomization (BIMMER) can be used to infer sparse networks of direct causal effects (DCEs) from phenome-scale GWAS summary statistics [28]. However, this process is implemented by inverse sparse regression under the assumption that the DCE matrix is sparse. Furthermore, BIMMER is very time consuming.

Multivariable MR (MVMR) [27, 29-30] is able to compute DCEs when there are multiple potential exposures and a single outcome. In MVMR, each genetic variant must satisfy the following criteria: (1) the variant is associated with at least one of the exposures; (2) the variant is independent of all confounders between exposures and outcomes; and (3) the variant is independent of the outcome conditional on the exposures and confounders. In this paper, we propose a novel algorithm called MRSL based on UVMR and MVMR for structural learning using summarized genetic data without requiring individual data. MRSL starts with an empty graph. For the first step, a marginal causal graph can be obtained by using bi-directional MR [31-32] in pairs. This process is a UVMR analysis, which estimates all total causal effects for each pair of variables. For the second step, we find the topological sorting for the marginal causal graph. For the third step, based on the above topological sorting [33], MVMR is performed to estimate the direct effects for each pair of variables by adjusting for the genetic associations with the phenotypes in a sufficient separating set. After an iteration process of step 2 and step 3, MRSL outputs a true causal graph. We apply MRSL to 26 biomarkers and 44 ICD10-defined diseases in 337,198 European from UK Biobank using summarized genetic data.

## Results

### Method overview

We present a novel algorithm MRSL for structural learning based on summarized genetic data. An illustration diagram of MRSL is displayed in Figure 1. Consider a DAG 𝒢 with *d* phenotypes {*X*_1_, *X* _2_, …, *X*_*d*_ }. U represents a set of unobserved confounders among *d* phenotypes. GWAS summary data for these *d* phenotypes are available. Generally, for continuous phenotypes, beta coefficients and their standard errors can be obtained from linear regression; for binary phenotypes, log(OR) coefficients and their standard errors can be obtained from logistic regression. Firstly, MRSL initializes the target causal graph with an empty graph and then obtains a marginal causal graph 𝒢_M_, using bi-directional MR in pairs of variables. The marginal causal graph 𝒢_M_ includes all the edges in the true causal graph 𝒢, but may add extra edges and spurious colliders. At the second step, we find the topological sorting of the nodes in the marginal causal graph using Depth First Search (DFS) algorithm [33-34]. The order of the topological sorting for the true causal graph 𝒢and the marginal causal graph 𝒢_M_ are the same. Based on this order, MVMR is performed to remove extra edges in 𝒢_M_. For each edge *X* _*p*_ → *X*_*q*_ in the marginal causal graph 𝒢_M_, we search a sufficient separating set 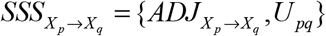 and adjust for genetic associations with 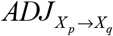 using MVMR to detect whether an edge is extra. The marginal causal graph is updated after each edge’s test. 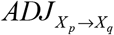can be searched in three ways: (1) all variables on the open paths from *X* _*p*_ to *X* _*q*_ ; (2) minimal sufficient adjustment set [1-2] and all the mediators from *X* _*p*_ to *X* _*q*_ ; (3) V\{ *X* _*p*_, *X* _*q*_ and *S*^*d*^ }, where *S*^*d*^ refers to the colliders where *X* _*p*_ and *X* _*q*_ have direct edges on them. For each edge *X* _*p*_ → *X*_*q*_ pointing to them in the marginal causal graph 𝒢_M_, if there is a sufficient separating set 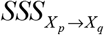 such that 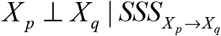 using MVMR, we delete the extra edge *X* _*p*_ → *X*_*q*_ in the true causal graph 𝒢. In addition, adjusting for the nodes in 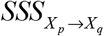 cannot unlock any blocked pathways in the true causal graph 𝒢. Thus the third step removes the extra edges in the graph 𝒢_M_. For the fourth step, we add an iteration step to perform MVMR in step 2 and step 3 again, using the graph obtained by step 3 as the initialization, until this graph converges. Detailed illustration of MRSL is shown in the Methods section.

**Figure 1.**
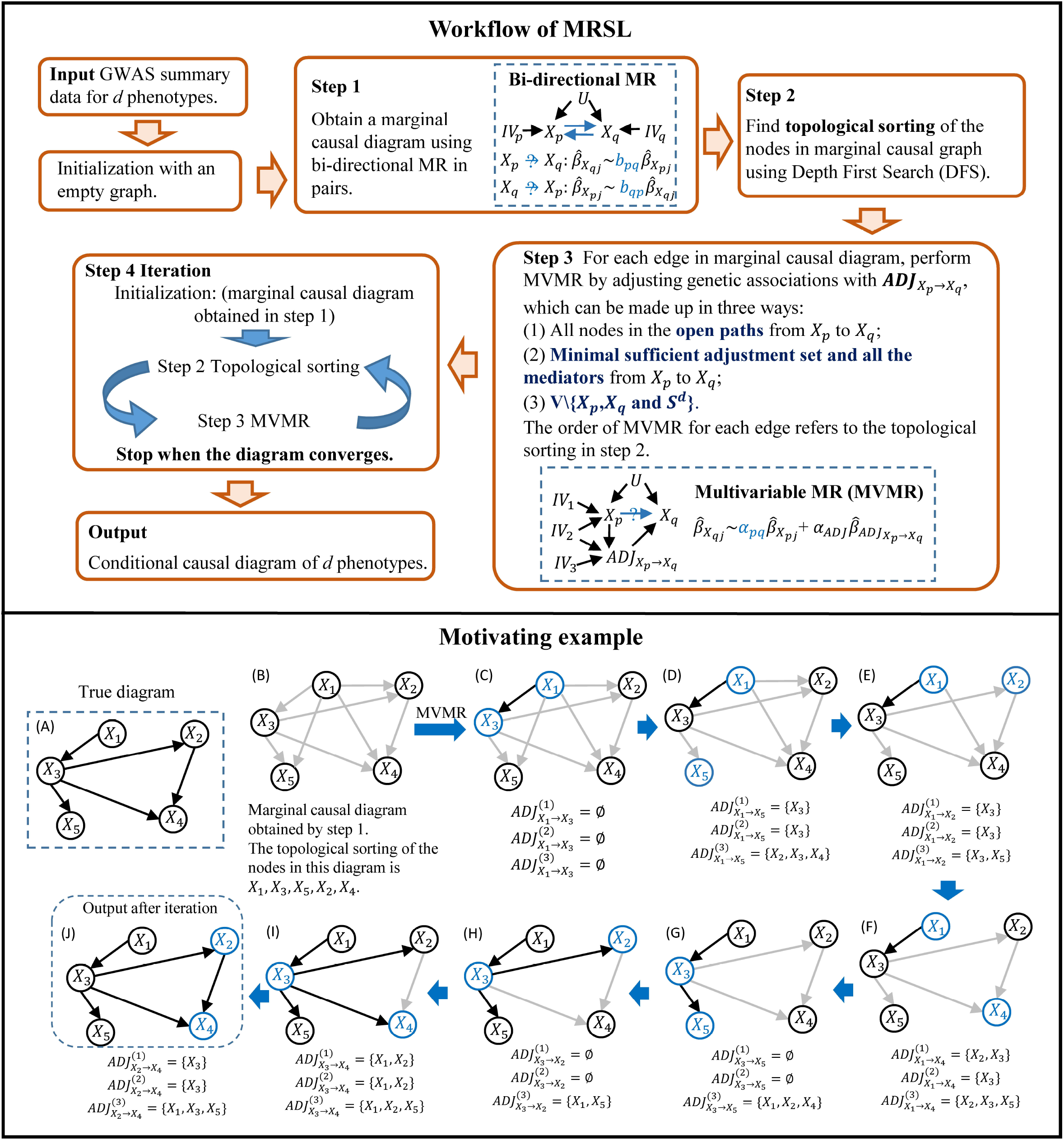
Workflow and the motivating example of MRSL algorithm. Confounders of *d* phenotypes are omitted. The input data is GWAS summary data for each phenotype. Initialization with an empty graph. For the Step 1, a marginal causal graph can be obtained using bi-directional MR in pairs. For the Step 2, the topological sorting of marginal causal graph should be found using Depth First Search (DFS). For the Step 3, MVMR is performed to remove extra edges in the marginal causal graph by adjusting for the genetic associations with phenotypes of three strategies of sufficient separating sets. The in the Step 4, iteration for step 2 and step 3 is performed until the graph converges. Finally MRSL outputs a conditional causal graph. **(A-J)** Motivating example with five nodes. (A) The true causal graph. (B) Marginal causal graph obtained by step 1. (C-J) Perform MVMR for each edge in graph (F) based on its topological sorting. Blue nodes denote the exposure and outcome we are interested in. MRSL outputs the graph (J). 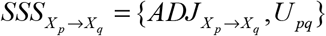 denotes the sufficient separating set from 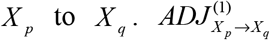 includes all nodes on the open paths from *X* _*p*_ to 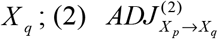 includes the elements in the minimal sufficient adjustment set and all the mediators from *X*_*p*_ to 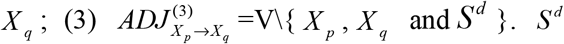 refers to the colliders where *X* _*p*_ and *X* _*q*_ have direct edges on them. In the motivating example, we omit the unobserved confounders U and the instrumental variables for each phenotypes used in MR.

We provide a motivating example to illustrate the workflow of MRSL (Figure 1 A-F). The true causal diagram is Figure 1 (A). The input are the GWAS summary datasets of five phenotypes. Firstly, bidirectional MR in pairs of five variables are performed to obtain a marginal causal graph (Figure 1 (B)). We find that its topological sorting is {*X* _1_, *X* _3_, *X* _5_, *X* _2_, *X* _4_ }. Then we perform MVMR varying across each edge in Figure 1 (B) to detect whether the edge is extra. In this stage, we adjust for the genetic associations with phenotypes in 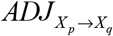 for each MVMR. We firstly focus on the edge *X*_1_ → *X* _3_. The other three nodes are not included in 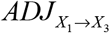 thus this edge retains. Then we are interested in the edge *X*_1_ → *X* _5_, and 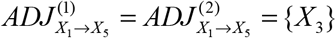 and 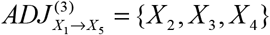. MVMR is performed by adjusting for the genetic associations with phenotypes in 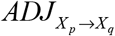 and the result reveals a null direct causal relationship between *X*_1_ and *X* _5_. Thus the edge *X*_1_ → *X* _5_ is removed. The rest edges are tested by the same ways (Figure 1 C-J). After all the edges in Figure 1 (B) are tested once, we obtained Figure 1 (J). An iteration for step 2 and step 3 are performed using this graph and stop when the causal graph converges. Finally MRSL output the target causal diagram.

### Simulations

Firstly we conducted a simulation study to evaluate the performance of MVMR to estimate the direct causal effect of an exposure (X) on an outcome (Y) when adjusting for a collider (S), a mediator (M) or a measured confounder (C), respectively (Figure 2 A-C). We considered seven kinds of available SNPs as IVs: (1) *G*_1_ : SNPs only associated with X; (2) *G* _2_ : SNPs associated with X and adjusting variable; (3) *G* _3_ : SNPs only associated with the adjusting variable; (4) *G*_1_ + *G*_3_ ; (5) *G*_1_ + *G*_2_ ; (6) *G*_2_ + *G*_3_ ; (7) *G*_1_ + *G*_2_ + *G*_3_. Details of data generation are shown in Methods section and Supplementary Notes.

**Figure 2.**
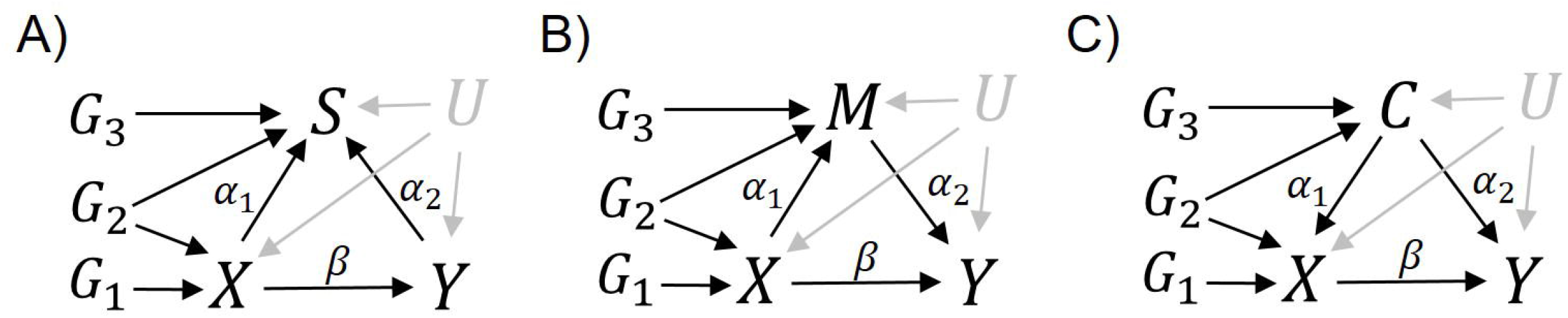
Diagrams for simulation study 1. X, exposure; Y, outcome; S, collider; M, mediator; C, measured confounder; U, unobserved confounder. *β* is the causal effect of X on Y. *α*_1_ is the causal effect of X on S/M or C on X. *α* _2_ is the causal effect of C/M on Y or Y on S. *G*_1_ are SNPs only associated with X. *G* _2_ are SNPs associated with X and adjusting variable. *G* _3_ are SNPs only associated with the adjusting variable

Figure 3 shows the results of MVMR when there are 100 IVs. When adjusting for the collider (S), the causal estimation is biased whatever IVs are used, and this bias becomes larger as the increasing of other edges’ effects. When adjusting for the mediator (M), causal estimation is unbiased when the IVs are *G* _2_ only, *G*_1_ + *G*_3_ or *G*_1_ + *G*_2_ + *G*_3_. The type I error rates of causal estimations are stable around 0.05 under these three kinds of IV selection. The causal estimations under *G*_1_ + *G*_3_ or *G*_1_ + *G*_2_ + *G*_3_ have higher power than that under *G* _2_ only when the causal effect is 0.1. *G* _3_ are terrible IVs with large biased estimations whatever variables adjusting for. When adjusting for the measured confounder (C), *G*_1_ only, *G* _2_ only, *G*_1_ + *G*_2_ and *G*_1_ + *G*_2_ + *G*_3_ are good choices for IVs in MVMR with unbiased estimations. When *G* _2_ only and *G*_1_ + *G*_2_ are IVs, the type I error rates of causal estimations are stable around 0.05, whereas when *G*_1_ only and *G*_1_ + *G*_2_ + *G*_3_ are IVs, the type I error rates are a little bit inflated. The power of causal estimation is high whatever kinds of IVs are selected except *G* _2_ only. The simulation results of 6, 20 and 60 IVs are shown in the Supplementary Fig.1-4. In practice, practitioners always don’t know the roles of the adjusting variables. Consider the above three graphs together, *G*_1_ + *G*_2_ + *G*_3_ is the best choice of IVs when performing MVMR.

**Figure 3.**
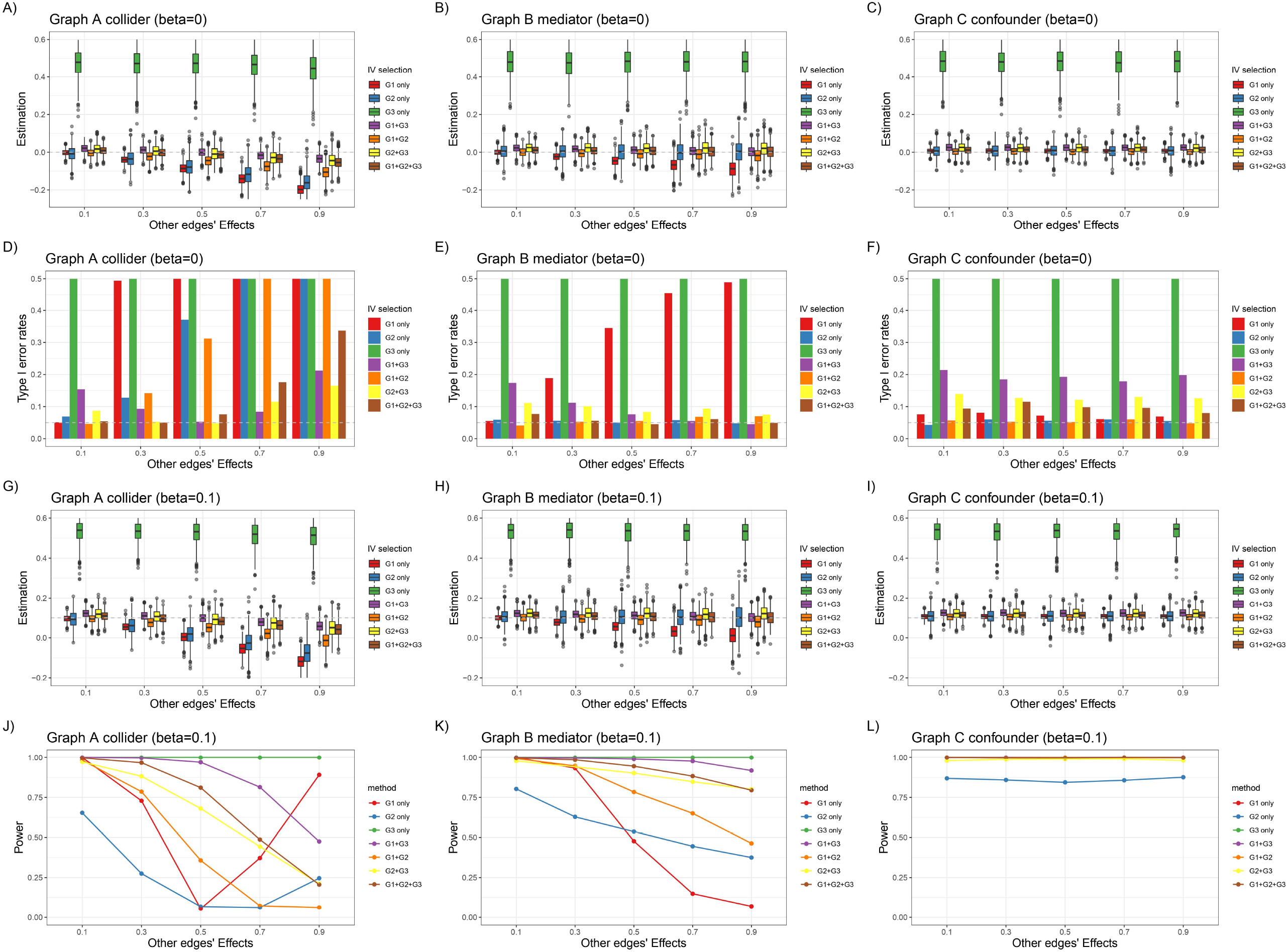
Simulation results of MVMR in simulation study 1. (A-C) Causal effect estimation of X on Y when causal effect *β* = 0; (D-F) Type I error rates of causal effect estimation of X on Y when causal effect *β* = 0 ; (G-I) Causal effect estimation of X on Y when causal effect *β* = 0.1; (J-L) Statistical power of causal effect estimation of X on Y when causal effect *β* = 0.1. The x-axis represent the other edges’ effect (*α*_1_ and *α*_2_ in Figure 2).

For the simulation study 2, we conducted a simulation study for continuous and binary phenotypes to learn the structures of random graphs, respectively. We generated the random graphs with 5, 10 and 15 nodes, respectively. Considering the different complexity of network, we set the probability of each edge to be present in a graph as 0.2, 0.5 and 0.8. The effect of each edge follows uniform distribution with four settings: U(0,0.25), U(0.25,0.5), U(0.5,0.75) and U(0.75,1). We varied across the number of SNPs *g*_1_ and *g* _2_ as 5, 10, 20, 30, 40 and 50, respectively. We compared MRSL with eight published methods: BIMMER [28], cGAUGE based on IVW, MR Egger and MR PRESSO [25], HC algorithm incorporating genetic anchors [23] (based on genetic risk score or the most significant SNP) and MRPC algorithm [24] (based on genetic risk score or the most significant SNP). Details of data generation are shown in Methods section.

Simulation results of 10 continuous nodes are shown in Figure 4-5 and Table 1. Figure 4 demonstrates the F1 score with different edges’ effects and complexity of network. Figure 5 shows the mean of precision and recall when there are 20 IVs. Results of precision and recall when there are 5, 10, 30, 40, 50 IVs are shown in Supplementary Fig. 5-8. When the network is simple (prob=0.2), F1 score of MRSL is the highest and the performance of three adjustment categories are similar. As the network become more complex, the F1 score of MRSL when adjusting for all nodes on the open paths and minimum separated set is decreasing, whereas MRSL when adjusting for V\{ *X*_*p*_, *X*_*q*_, *S*^*d*^ and U} still has the highest F1 score. The recall of the former is smaller than the latter as the edges’ effects and the complexity of graph rising. When the edges’ effects are small, F1 score of MRSL rises as the number of IVs increasing. When the edges’ effects are large, F1 score of MRSL decreases as the number of IVs increasing due to the reduction of precision. This may because in simulation study 1, as the number of IVs increasing, the type I error rate of *G*_1_ + *G*_2_ + *G*_3_ is even more inflated and lead to the increase of false negative rate. Besides, the power of causal estimation using MVMR is decreasing as other edges’ effects increasing. In addition, the number of adjusting variables is increasing as the network become more complex, then the accuracy of causal estimation using MVMR is reducing. Table 1 shows the computing time of MRSL and other eight methods when there are 5, 20 and 50 IVs. MRSL has the fastest computing time among these methods. Computing time of all the methods with 10, 30 and 40 IVs are listed in Supplementary Table 1. The results of MRSL with 10 binary nodes are similar as that with continuous nodes (Supplementary Fig. 9-13 and Supplementary Table 2). As the number of nodes increasing in the network, the F1 score of MRSL is reducing especially when the network is complex. Results of 5 and 15 nodes are shown in Supplementary Fig. 14-33 and Supplementary Table 3-6.

**Table 1.**
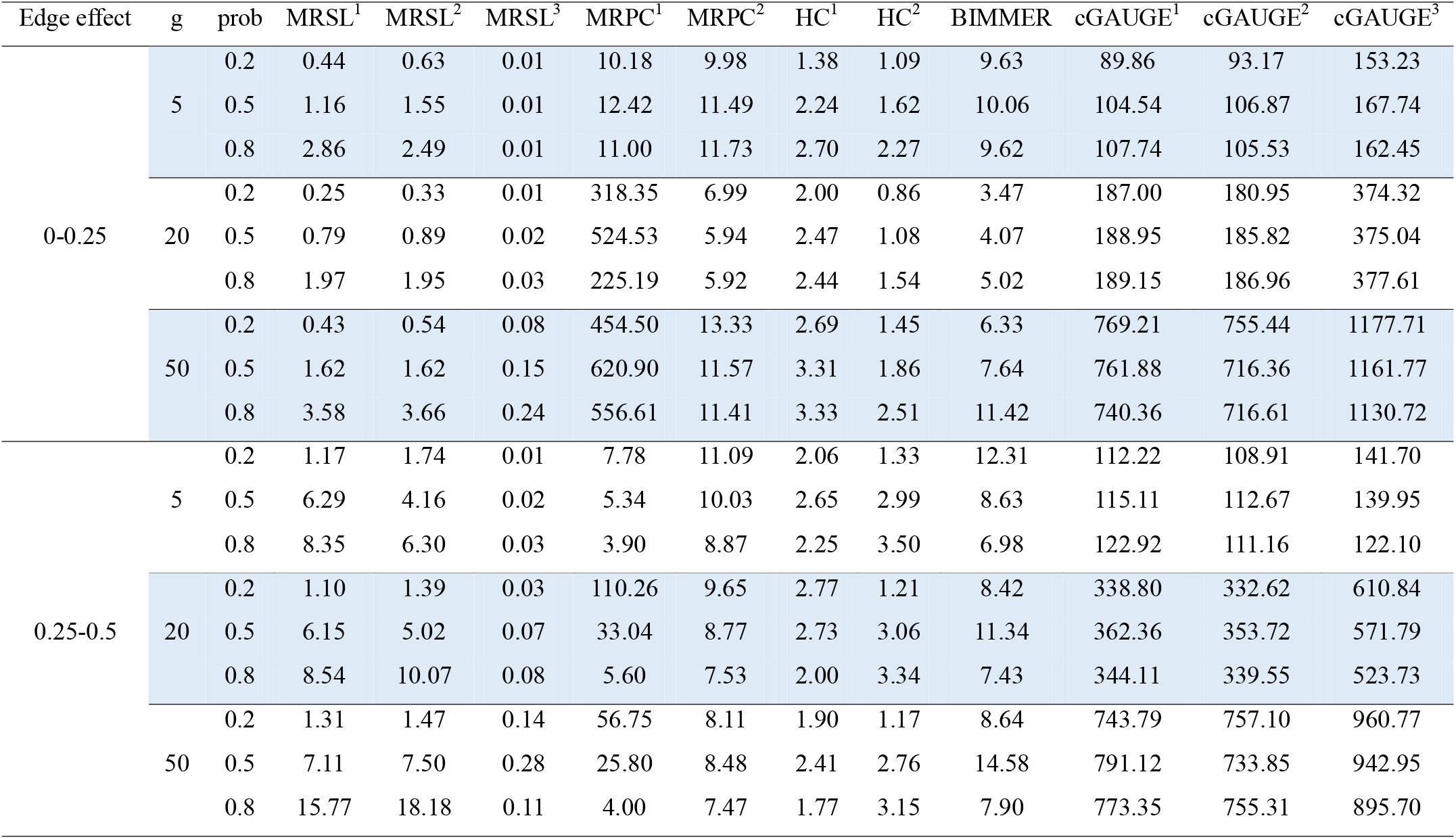
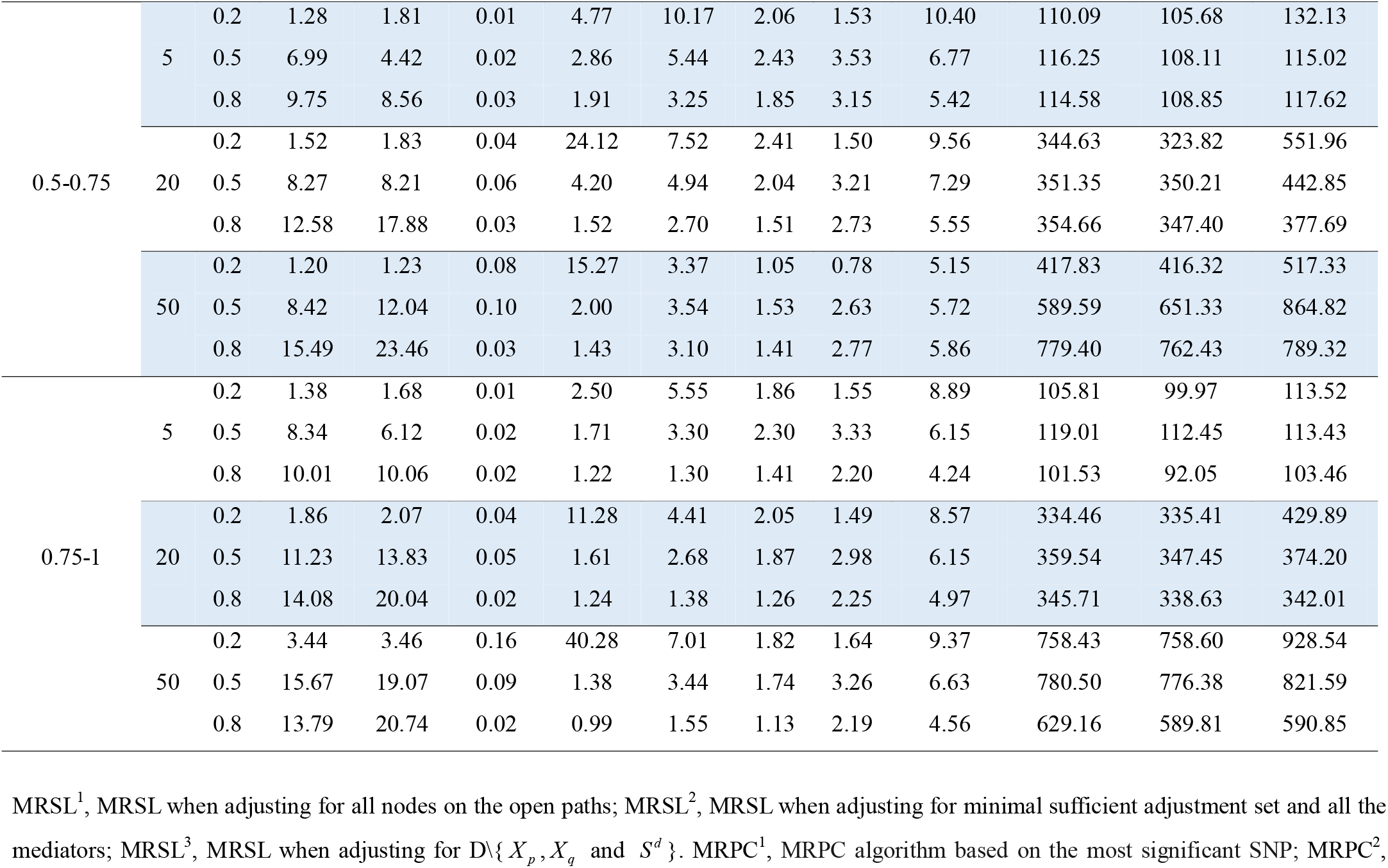
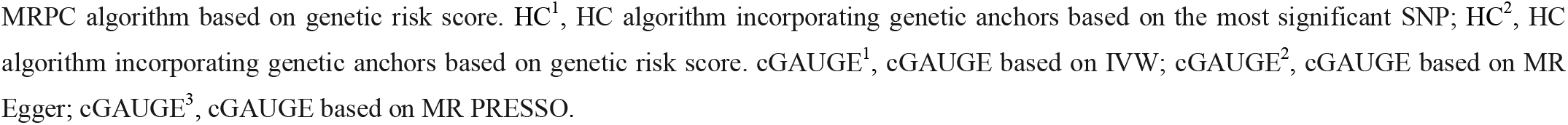
Computing time with network of 10 continuous nodes in simulation study 2 (seconds).

**Figure 4.**
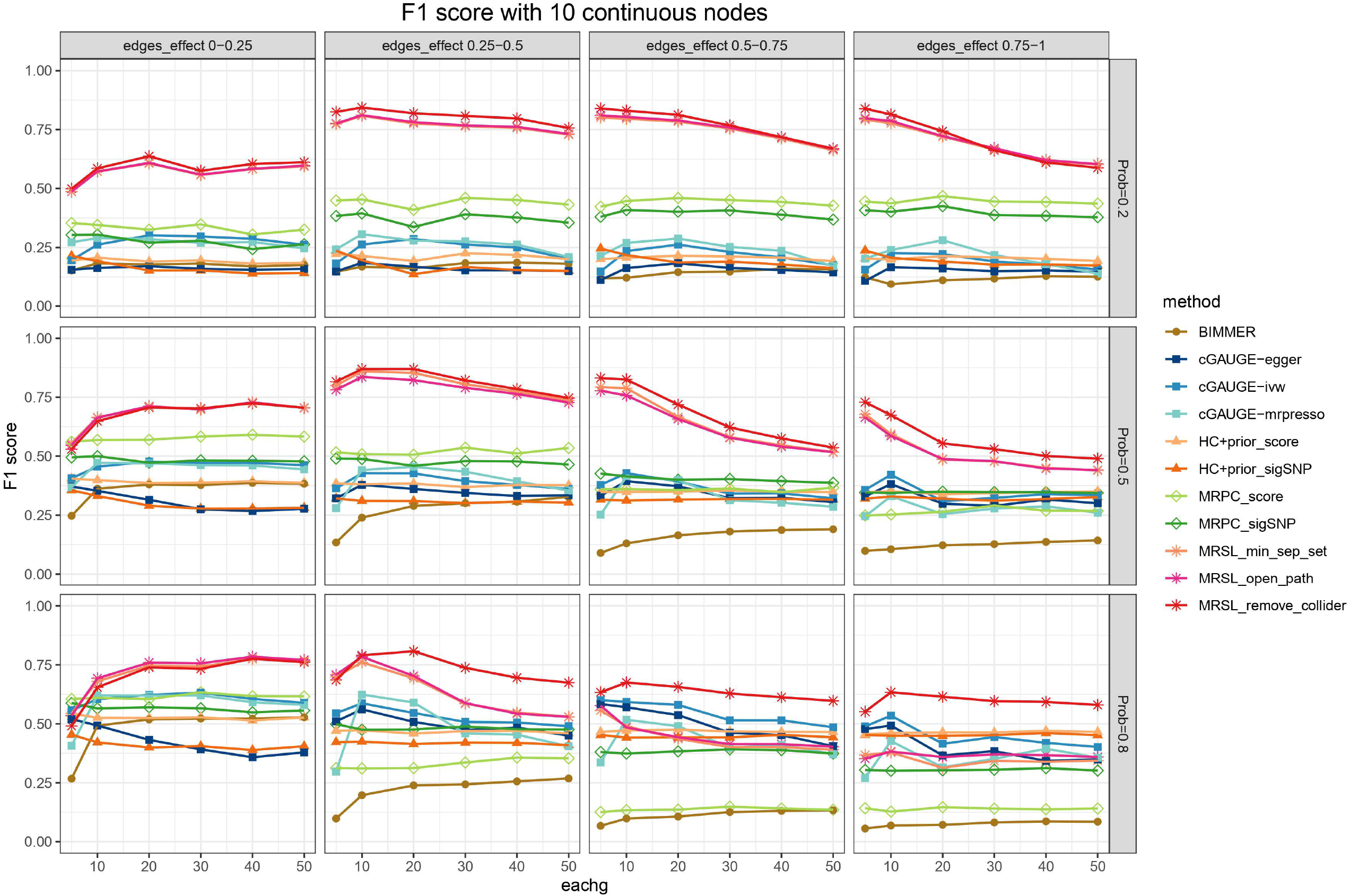
F1 score with 10 continuous nodes. The x axis represents the number of IVs. Considering the different complexity of network, we set the probability of each edge to be present in a graph as 0.2, 0.5 and 0.8. The effects of any two traits *β* follows uniform distribution with four parameter settings: U(0,0.25), U(0.25,0.5), U(0.5,0.75) and U(0.75,1) for continuous nodes. MRSL_min_sep_set indicates the MRSL adjusting for minimal sufficient adjustment set and all the mediators; MRSL_open_path indicates the MRSL adjusting for all the nodes on the open paths; MRSL_remove_collider indicates the MRSL adjusting for V\{ *X* _*p*_, *X* _*q*_ and *S* ^*d*^ }.

**Figure 5.**
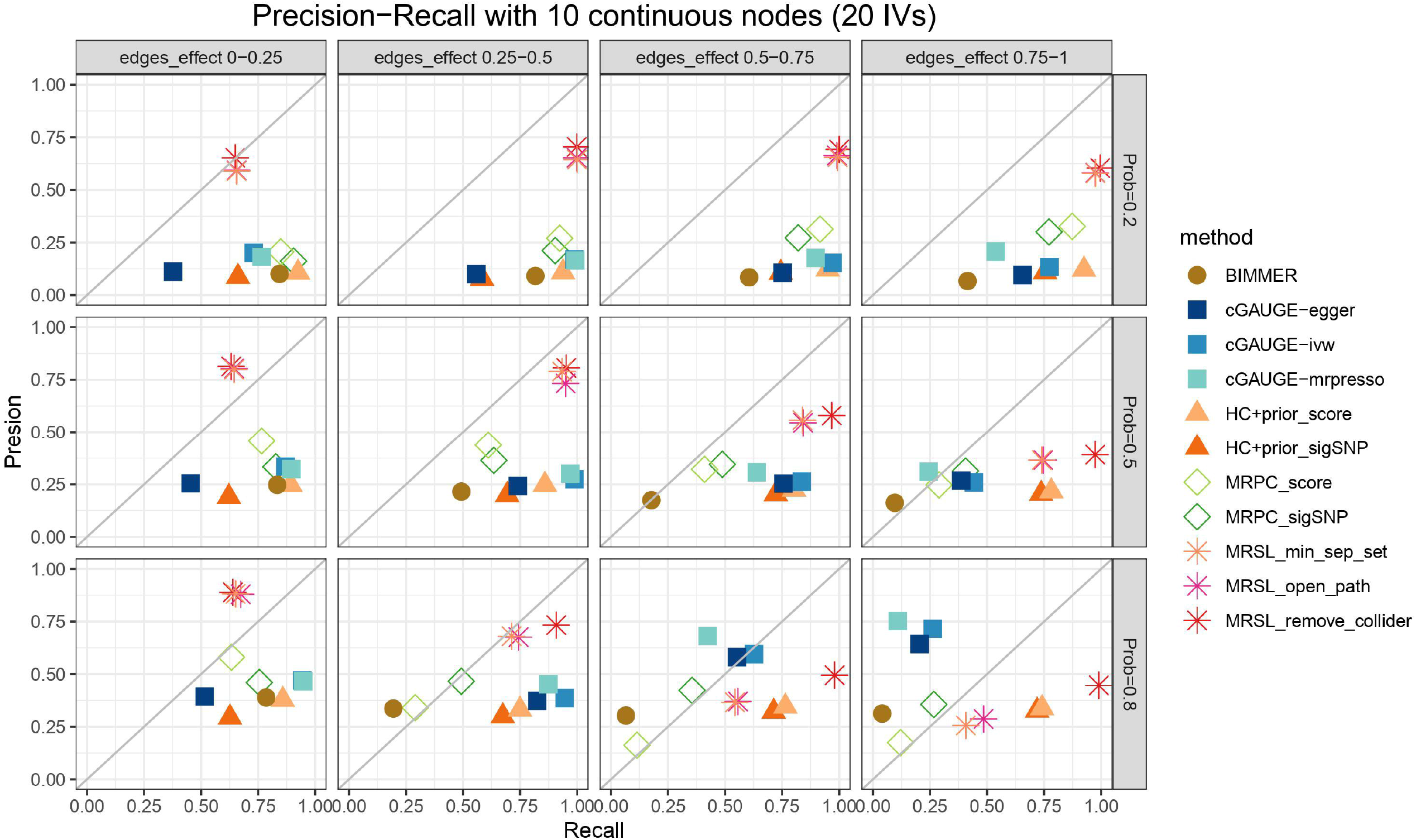
Precision and recall with 10 continuous nodes when there is 20 IVs.

For the simulation study 3, in order to evaluate the performance of MRSL in practical application, we choose three fixed networks (Figure 6), which are representative examples in practice, including MAGIC-NIAB [35], ASIA (lung cancer network) [36] and Healthcare Cost [2], with continuous, binary and mixed nodes, respectively. Details of these three networks are illustrated in the Method section. Figure 7 shows the F1 scores of MRSL and eight methods when learning three networks. MRSL has the best performance among all methods. The performances of ASIA (binary) and MAGIC-NIAB (continuous) are similar as that in simulation study 2. For the mixed variables network Healthcare Cost, F1 score is lower than that of ASIA and MAGIC-NIAB. And when the edges’ effects are larger, MRSL when adjusting for adjusting for all nodes on the open paths and minimum separated set is a little bit larger F1 score than MRSL when adjusting for V\{ *X*_*p*_, *X*_*q*_, *S*^*d*^ and U} because of higher precision. The precision and recall are shown in Supplementary Fig. 34-45.

**Figure 6.**
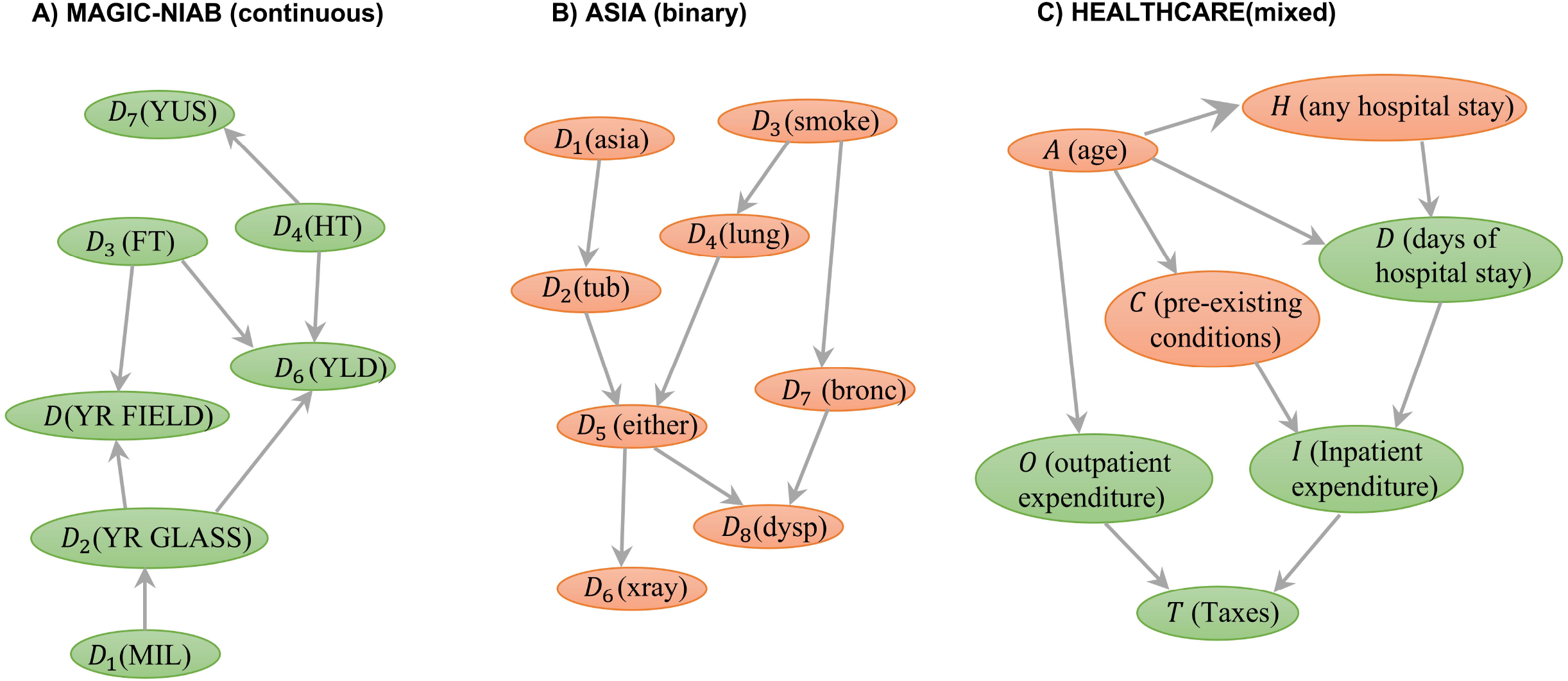
Diagrams of practical example in the simulation study 3. Green and yellow circles denote continuous and binary variables, respectively. MAGIC, Multiparent Advanced Generation Inter-Cross; NIAB, National Institute of Agricultural Botany; YLD, yield; FT, flowering time; HT, height; YR.GLASS, yellow rust in the glasshouse; YR.FIELD, yellow rust in the field; FUS, Fusarium; MIL, mildew.

**Figure 7.**
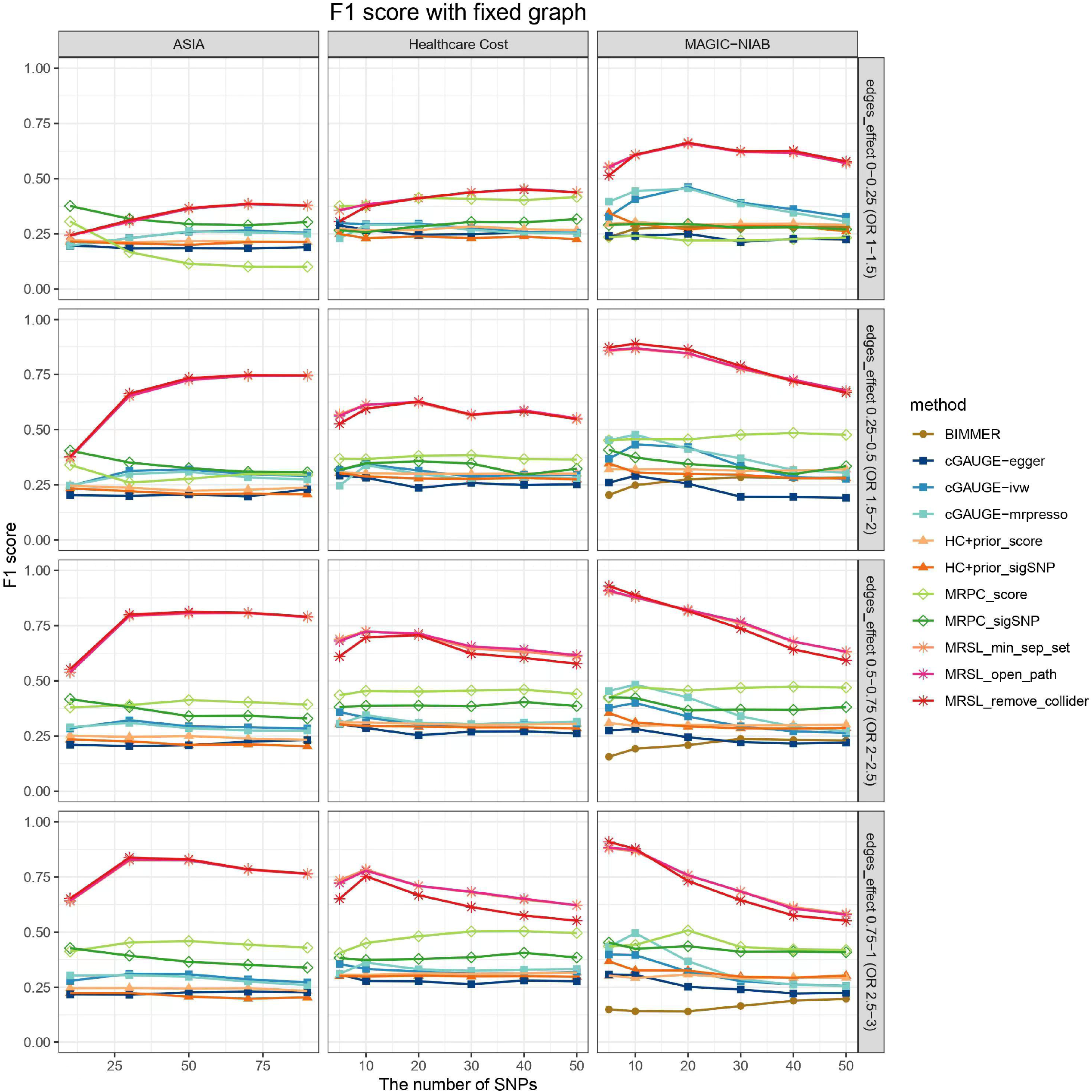
F1 score of MRSL when learning the structure of MAGIC-NIAB, ASIA and Healthcare Cost.

### Applied example: network of 44 diseases and 26 biomarkers

We applied MRSL to learn the network of 44 diseases with ICD 10 coding and 26 biomarkers using GWAS summary data in UK Biobank. The list of these 70 traits are shown in the Supplementary Table 7. Figure 8 A) shows the marginal causal graph by MRSL step 1, resulting in 70 nodes and 388 edges. Figure 8 B) shows the conditional causal graph obtained by MVMR adjusting for V\{ *X*_*p*_, *X*_*q*_, *S*^*d*^ and U}, resulting in 69 nodes and 192 edges. This result was obtained by removing 196 direct edges induced by mediation pathways after bonferroni correction. Figure 8 C) shows the causal mediation pathways from biomarkers on each diseases. Vitamin D, Total protein, Urate and Urea are root causes for nearly all the mediation pathways of diseases [37-39]

**Figure 8.**
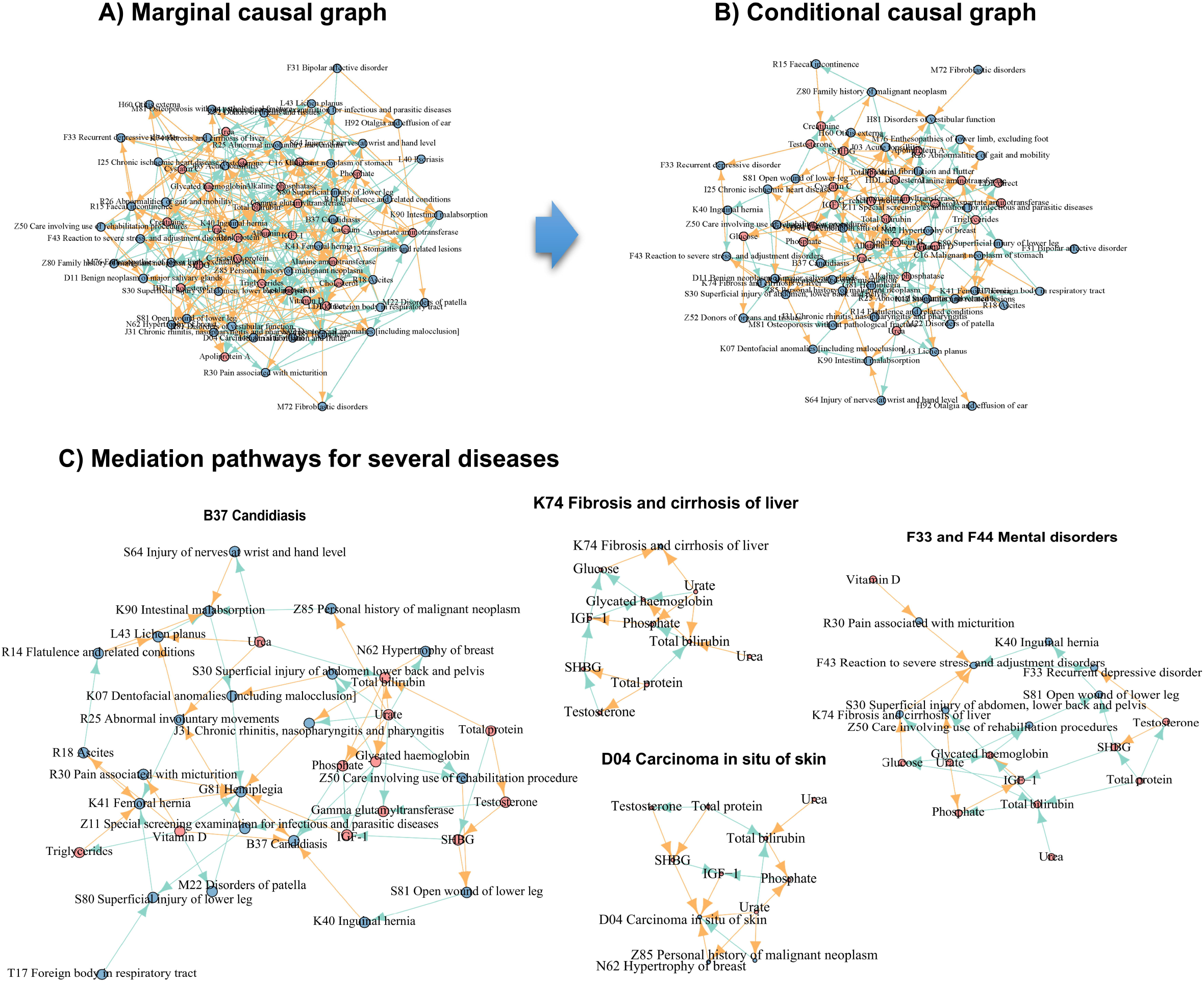
Structures of 26 biomarkers and 44 diseases in applied example 2. A) the marginal causal graph obtained by MRSL-step1; B) the conditional causal graph obtained by MRSL-step2; C) mediation pathways of several diseases. The color of an edge indicates whether the effect is positive or negative. Yellow edges represent the positive effects and green edges represent the negative effects. Blue nodes represent the ICD10-defiend diseases. Pink nodes represent the biomarkers.

Most of causal links are expected and have clear interpretation of biological pathways or have been confirmed by experiments. Such as B37 Candidiasis, Vitamin D [40], K40 Inguinal hernia [41] and G81 Hemiplegia [42] are direct risk factors; Phosphate [43] and Glycated haemoglobin [44] are direct protective factors. For F33 Recurrent depressive disorder, Testosterone have a positive effect on F33 [45]. Other biomarkers affect F33 through F43 Reaction to severe stress, and adjustment disorders. IGF-1 [46] directly influence the risk of D04 Carcinoma in situ of skin. Vitamin D directly affect G81 Hemiplegia with a protective effect [47]. Glucose [48] and Urate [49] are risk factors of K74 Fibrosis and cirrhosis of liver. Biomarkers have causal effects on K90 Intestinal malabsorption through R14 Flatulence [50] and related conditions and L43 Lichen planus [51].

Several novel causal links are founded and supported by clinical case report or observational studies. For example, for C16 Malignant neoplasm of stomach, Urate [52], T17 Foreign body in respiratory tract [53-54], F31 Bipolar affective disorder [55] and K41 Femoral hernia [56] have negative causal effects on the Malignant neoplasm of stomach. Urate [57] positively affects the risk of Carcinoma in situ of skin. IGF-1 [58-59] and K07 Dentofacial anomalies [including malocclusion] directly increase the risk of G81 Hemiplegia. For H60 Otitis externa, Glycated haemoglobin [60] is protective factor, J03 Acute tonsillitis is a risk factor of Otitis externa. For J03 Acute tonsillitis, IGF-1 [61], HDL cholesterol [62], total protein [63] and total bilirubin [64] positively affect J03 Acute tonsillitis. Urate [65] is a risk factor for K12 Stomatitis and related lesions. Urate negatively affects the risk of M81 Osteoporosis without pathological fracture [66] whereas R25 Abnormal involuntary movements is a risk factor [67].

## Discussion

In this work, we present a novel algorithm called MRSL based on UVMR and MVMR for structural learning. Our method is flexible as it requires only summarized genetic data. Besides, MRSL relaxes the causal sufficiency assumption and can be implemented with fast computing speed and outputs a conditional causal graph with directed causal effects. MRSL consists of four steps: step 1 outputs a marginal causal graph using bi-directional MR in pairs, step 2 find the topological sorting of marginal causal graph, step 3 outputs a conditional causal graph by MVMR and step 4 is an iteration process of step 2 and 3. The marginal causal graph reveals the total causal relationships of each pair of variables and the conditional causal graph identifies the mediation pathways then discloses the direct effects of each pair of variables. The application to 26 biomarkers and 44 ICD10-defined diseases cover lots of expected causal links which have biological interpretations and several new links supported by clinical case reports or previous observational literatures.

The core of MRSL is MR analysis, thus how well MRSL performs depends on the performance of MR decides. The first point we need to focus on is the selection of IVs. For the bi-directional MR, an alternative choice is the SNPs only associated with the exposure but not associated with other variables in the network. To some extent, this can block nearly all pleiotropic pathways. For MVMR, we firstly conducted a simulation study to choose the most valid IVs. Here we should consider the valid IVs which makes the MVMR perform best when adjusting for the collider, mediator and confounder simultaneously. From the results of simulation 1, *G*_1_ + *G*_2_ + *G*_3_ is the best choice. This is supported by previous literature [68-69]. It is necessary to have as many IVs as possible, that is, including genetic variables that are associated with at least one exposure and removing the instruments that only strong with one exposure will lead to a loss of precision in the estimation or other potential bias. We only focus on the effect of particular exposure on the outcome using MVMR each time, thus we only force positive association with respect to the exposure we are interested in [70]. It has no influence on our results although this may change the sign of the association with respect to the adjusting variables. In this work, we use univariable and multivariable IVW as the main methods. MRSL can be extend to use other UVMR methods, such as pleiotropy-robust methods (e.g. MR-Egger [71], the weighted median method [72], the mode-based estimate method [73], MR-RAPS [74] and contamination mixture method [75], etc), and MVMR methods (e.g. MVMR-Egger, MVMR-Robust, MVMR-Median, and MVMR-Lasso [70], etc) instead of IVW. However, combination of these methods in MRSL is a time consuming process and may lose precision due to the low accuracy of methods themselves.

In the second step of MRSL, we present three strategies for adjusting variables in MVMR with the complement of graph theory in causal inference. Because MR overcome the influence of unobserved confounding, we exclude U in the three sets of adjusting variables. Another point we pay attention to is that whether these three sets of adjusting variables are the same in the marginal causal graph and the true causal graph, or in other words, we have two questions: does adjusting these variables in the marginal causal graph unlock the blocked pathways in the true causal graph? or not completely block the mediation pathways in the true causal graph? For these two questions, we proposes Lemma 1-3 and Theorem 2. The first way is adjusting for all nodes on the open paths in the marginal causal graph. This enables all the open paths between two variables can be blocked, whether mediation pathways or confounding pathways. This adjusting set doesn’t include the spurious colliders in the marginal causal graph. For the second way, minimal separating set may include spurious colliders at the cost of include other confounders or mediators to ensure the separation of two variables. This not only block the pathways in the true causal graph, but also block all the pathways include spurious pathways in the marginal causal graph. The third adjusting set is the most conservative set, that is, adjusting for all the variables excluding colliders, which are particular colliders that must have direct edges on the two variables we are interested in. In all, the second step of MRSL is a process to remove extra edges in marginal causal graph, and obtain a conditional causal graph.

Combination of graph theory and MVMR is a unique property of our algorithm, and we utilize this novel property into causal discovery to improve the precision and recall. Our method can be easily implemented only using GWAS summary data, which is public available for the most phenotypes as the emergence of a large number GWAS studies with huge sample size. Published MR-based algorithm such as cGAUGE, requires the individual-level data and are thus not as easily available as the GWAS summary statistics, and is very time consuming [25]; BIMMER are implemented based on the complex inverse sparse regression and obtained an approximately estimation of DCE matrix, this require time roughly 𝒪(*κd*^4^) for *d* phenotypes [28]. In the result of simulation study 2-3, we found that the computing time of MRSL is only around 1/100 of BIMMER, and 1/1000 of cGAUGE, respectively. MRSL has two-fold higher F1 score than other eight methods when the network is simple. Also, MRSL outputs the unbiased direct effect of each pair of variables. Moreover, MRSL can be applied into the structure with feedback loops between any two variables, because our main MR method IVW can powerfully deal with the case of bi-directional causal relationship between two variables [28,76]. Similar to MR analysis, GWAS summary data of *d* phenotypes should from the homogenous population. We also need to pay more attention to other issues, such as measurement error, selection bias and missing data, etc, in the future.

In conclusion, we proposed a novel algorithm, utilizing the combination of graph theory and MR into causal discovery to learn the conditional causal graph. We look forward to offer constructive suggestions for disease diagnostic and apply our method beyond the scope considered here.

## Methods

### MRSL

We consider an algorithm MRSL for structural learning based on summarized genetic data. An illustration diagram of MRSL is displayed in Figure 1. Assume a DAG 𝒢 =<*V, E* > with unobserved confounders, where V is a set of nodes and E is a set of pairs of nodes. Assume we are interested in *d* phenotypes {*X*_1_, *X* _2_, …, *X*_*d*_ }. U represents unobserved confounders among *d* phenotypes. E_𝒢_ and E_𝒢M_ denote all the pairs of nodes for directed edges in the true causal graph 𝒢 and the marginal causal graph 𝒢_M_, respectively. S_𝒢_ and S_𝒢M_ denote all the colliders in the true causal graph 𝒢 and the marginal causal graph 𝒢_M_, respectively. For convenience, the unobserved confounders among phenotypes in Figure 1 are omitted. GWAS summary data for these *d* phenotypes are available. Generally, for a continuous phenotype, beta coefficient and its standard error can be obtained from linear regression; for a binary phenotype, log(OR) coefficient and its standard error can be obtained from logistic regression. MRSL initializes with an empty graph. The first step of MRSL is to obtain a marginal causal graph, denoted by 𝒢_M_, using bi-directional MR in pairs. Here, we choose the univariable inverse-variance weighted (IVW) method as the main method because of its high accuracy.

#### Assumption 1.

For an exposure and an outcome, the independent IVs in UVMR must satisfy (**Relevance**) IVs are strongly associated with the exposure; (**Exchangeability**) IVs are independent with unobserved confounders between the exposure and the outcome; (**Exclusion restriction**) IVs affect the outcome only through the exposure.

For a pair of phenotypes *X* _*p*_ and *X* _*q*_, we firstly focus on the causal effect of *X* _*p*_ on *X* _*q*_. We select *J* _*p*_ IVs for *X* _*p*_ to perform weighted regression:

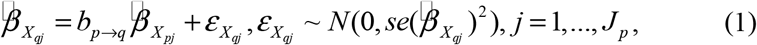

Where 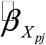 and 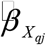 are genetic associations for *X* _*p*_ and *X* _*q*_ on j-th IVs in linear regression in GWAS, respectively. 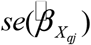 is the standard error of 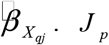 IVs for *X* _*p*_ must satisfy the Assumption 1. For the reverse direction, we select *J*_*q*_ IVs for *X* _*q*_ to perform weighted regression:

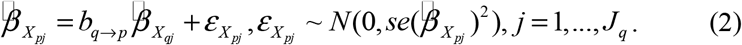

*b*_*p*→*q*_ or *b*_*q*→ *p*_ represents the total effect of *X* _*p*_ on *X* _*q*_ or *X* _*q*_ on *X* _*p*_, respectively. Similarly, *J*_*q*_ IVs for *X* _*q*_ must satisfy the Assumption 1. Wald tests for the total effect estimations can be used to test whether there are causal pathways from *X* _*p*_ to *X* _*q*_ or *X* _*q*_ to *X* _*p*_, respectively.

#### Assumption 2 (Causal Markov condition)

Each variable is independent of its non-descendants given its parents in graph 𝒢.

#### Assumption 3 (Faithfulness assumption)

All independencies embedded in the observed distribution 𝕡 are stable and are invariant to changes in parameterization. Thus, it implies (together with d-separation) that (*X* ⊥ *Y* | *Z*) _𝕡_ ⟺(*X* ⊥*Y* | *Z*)_𝒢_.

#### Lemma 1.

For the true causal graph 𝒢 and the marginal causal graph 𝒢_M_, E_𝒢_⊆E_𝒢M_ and S_𝒢_⊆S_𝒢M_.

For the second step, we find the topological sorting *T*_𝒢M_ in marginal causal graph 𝒢_M_ using DFS [33-34]. Topological sorting for a DAG is a linear ordering of vertices such that for every directed edge *X* _*p*_ → *X*_*q*_, vertex *X* _*p*_ comes before *X* _*q*_ in the ordering. This ensures that parent nodes will be ordered before their child nodes, and honors the forward direction of edges in the ordering. The DFS algorithm loops through each node of the graph, in an arbitrary order. DFS terminates when it hits any node that has already been visited since the beginning of the topological sort or the node has no outgoing edges. Each node X gets prepended to the output list only after considering all other nodes which depend on X (all descendants of X in the graph).

#### Lemma 2 (Topological sorting invariance)

The topological sorting of the true causal graph 𝒢 and the marginal causal graph 𝒢_M_ are the same *T*_𝒢_*=T*_𝒢M_.

In the third step, MVMR is performed to remove E_𝒢M_\E_𝒢_ in 𝒢_M_, and obtain the true causal graph. For each edge in 𝒢_M_ (e.g. *X* _*p*_ → *X*_*q*_), we detect whether this edge exists after adjusting for the genetic associations with the phenotypes in a sufficient separating set using MVMR.

#### Assumption 4.

For an exposure, a set of covariates and an outcome, the independent IVs in MVMR must satisfy (**Relevance**) IVs are strongly associated with the exposure and covariates; (**Exchangeability**) IVs are independent with any unobserved confounders among the exposure, adjusting variables and the outcome; (**Exclusion restriction**) IVs affect the outcome only through the exposure or covariates.

For an edge *X* _*p*_ → *X*_*q*_ in 𝒢_M_, 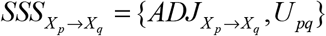denotes the sufficient separating set. Then we can perform multivariable IVW by following weighted regression of *X* _*q*_ on *X* _*p*_ adjustment for 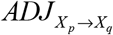:

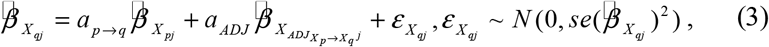

where *a*_*p*→*q*_ is the direct effect of *X* _*p*_ on *X* _*q*_ not through mediators and confounders in 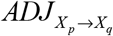. The Wald test for estimation of *a*_*p*→*q*_ can be used to test whether there is a direct edge from *X* _*p*_ to *X* _*q*_. IVs for the regression (3) above must satisfy the Assumption 4. This kind of IV can be obtained by statistical filtering criteria (see application examples) or ImpIV filter and ExSep test [25].

For MVMR, the sufficient adjustment set 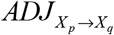 can be made up of three ways: (1) all nodes on the open paths from *X* _*p*_ to *X* _*q*_ ; (2) minimal sufficient adjustment set for confounders and all the mediators from *X* _*p*_ to *X* _*q*_ ; (3) V\{ *X* _*p*_, *X* _*q*_ and *S*^*d*^ }. *S*^*d*^ refers to a set of colliders where the two interested nodes have direct edges on them, e.g. for two nodes *X* _*p*_ and *X* _*q*_, the collider *S*_1_ in *X* _*p*_ → *S*_1_ ← *X*_*q*_ is included in *S* ^*d*^ but the collider *S*_2_ in *X* _*p*_ → *S*_2_ ← *C* → *X*_*q*_ is not included in *S*^*d*^. *S*^*d*^ in the graph 𝒢_M_ includes the colliders and the nodes not on the pathway from *X* _*p*_ to *X*_*q*_, but not includes any mediators, confounders or the nodes which are both mediators and confounders on the pathways from *X* _*p*_ to *X*_*q*_ in the true causal graph 𝒢. Note that one node can be a mediator and a collider simultaneously. For example, a graph 𝒟 with edges *X*_1_ → *X* _3_, *X* _3_ → *X* _2_, *X* _4_ → *X* _2_ and *X* _4_ → *X* _3_, *X*_3_ is a mediator on the path *X*_1_ → *X* _3_ → *X* _2_ and also a collider on the path *X*_1_ → *X* _3_ ← *X* _4_. This kind of node is included in the sufficient separating set 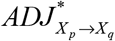 by including other nodes (e.g. *X* _4_) to ensure *X* _*p*_ and *X*_*q*_ are sufficiently separated.

We use 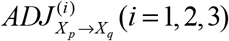 to denote that adjusting variables 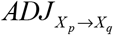 obtained by the *i*-th way. The order of MVMR varying across each edge is the same as the topological sorting in step 2. The topological sorting avoids the case in Supplementary Fig. 46, in which we show the MVMR adjusting for the genetic associations with the phenotypes in 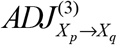 with and without topological sorting. For the latter, when we perform MVMR to test whether the edge *X* _4_ → *X*_1_ exists, the genetic association with *X* _3_ is adjusted in MVMR. However, in A3, *X* _3_ is a collider but is not included in *S*^*d*^, and *X* _2_ is included in *S*^*d*^. Then we perform MVMR 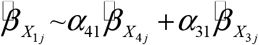 and the estimation for direct effect of *X* _4_ on *X* _1_ is biased because *X* _3_ is a collider. The bias formula of causal estimation when adjusting for a collider using MVMR are shown in Supplementary Notes. For the former, this kind of problem can be avoided and the process of removing edges is more accurate and faster after topological sorting.

#### Theorem 1

Under the Assumptions 2-4, for each edge *X* _*p*_ → *X*_*q*_ in the marginal causal graph 𝒢_M_, given a sufficient separating set 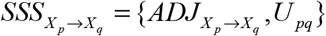 such that 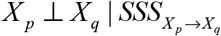, which can be tested by adjusting for genetic associations with 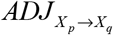 using MVMR,

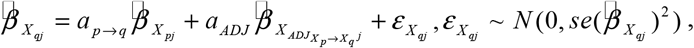

where *a*_*p*→*q*_ determines the existence of edge from *X* _*p*_ to *X* _*q*_ in the true causal graph 𝒢, then there is no direct edge from *X* _*p*_ to *X* _*q*_ in the true causal graph 𝒢.

Based on Theorem 1, the edges E_𝒢M_\ E_𝒢_ in the graph 𝒢_M_ are removed. After the third step, we add an iteration step to perform step 2 and 3 again, using the graph obtained in the previous step 3, until the graph is convergence. The aim of this step is to eliminate the random error and statistical test error in the UVMR and MVMR and increase the precision of MRSL.

### Simulation

We conduct three simulation studies to evaluate the performance of MRSL. Firstly, we conduct a simulation study to choose the optimal selection strategy of IVs for MVMR. Secondly, we evaluate the performance of MRSL and other eight published methods for structure discovery, when learning the structure of random graph with continuous and binary nodes, respectively. Thirdly, we select three representative graphs from previous literatures and access the performance of MRSL and other eight published methods.

### Simulation study 1 on IVs selection in MVMR

The basis of MRSL is MR, thus it is vital to select valid IVs. We firstly conduct a simulation study to evaluate the performance of MVMR when estimate the causal effect of an exposure (X) on an outcome (Y). We consider three roles of adjusting variables in MVMR: a collider (S), a mediator (M) or a measured confounder (C) in the causal pathway from X to Y (Figure 2 A-C). Based on the three figures, there are seven kinds of available SNPs as IVs: (1) *G*_1_: SNPs only associated with X; (2) *G* _2_ : SNPs associated with X and adjusting variable; (3) *G* _3_ : SNPs only associated with the adjusting variable; (4) *G*_1_ + *G*_3_ ; (5) *G*_1_ + *G*_2_ ; (6) *G*_2_ + *G*_3_ ; (7) *G*_1_ + *G*_2_ + *G*_3_. When the adjusting variable is a confounder, *G* _3_ is also associated with X, and it may be selected as instrumental variable because practitioners don’t know the true roles of the adjusting variables. The process of data generation is shown in Supplementary Notes. We generate 10,000 individuals and 1,000 repeated datasets. To access the performance of MVMR, we plot a boxplot to evaluate the estimation of causal effect of X on Y, and calculate the type I error rate for null causal effect and statistical power to detect the non-zero causal effect. The nominal level is set to 0.05.

### Simulation study 2 on MRSL with random graphs

To valid the utility of the MRSL method for learning structures, we conduct a simulation study for continuous and binary variables, respectively. Genetic IVs are generated from binomial distribution *B*(2,0.3). Let *Y* denote the N×*d* matrix of *d* phenotypes and *G* denote a N×J matrix of J SNPs. For continuous variables, *d* phenotypes are generated from the following model

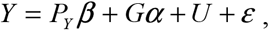

where *P*_*Y*_ represents the parent nodes of *Y, β* are the effects of *P*_*Y*_ on *Y* and generated from uniform distribution, *α* is a *d*×J matrix of effects of SNPs on phenotypes, *U* represents the confounding factors among *d* phenotypes and *ε* is the residual term following normal distribution *N*(0,1). For binary variables, *d* nodes are generated from the following model

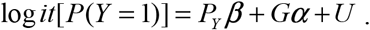

We generate the random graphs with 5, 10 and 15 nodes, respectively. Considering the different complexity of network, we set the probability of each edge to be present in a graph as 0.2, 0.5 and 0.8. In practice, there may be effects of different magnitude between traits, thus we consider *β* follows uniform distribution with four parameter settings: U(0,0.25), U(0.25,0.5), U(0.5,0.75) and U(0.75,1) for continuous nodes, odd ratio (OR) U(1,1.5), U(1.5,2), U(2,2.5) and U(2.5,3) for binary nodes. The IVs are assumed uncorrelated, and subdivided into two categories: (1) *g*_1_ SNPs that only predict one phenotype; (2) *g* _2_ SNPs that predict all the phenotypes simultaneously. The variance of each phenotype explained by all the SNPs is around 10%. We vary across the number of SNPs *g*_1_ and *g* _2_ with 5, 10, 20, 30, 40 and 50, respectively.

We compared our method with eight published methods: BIMMER [28], cGAUGE based on IVW, MR Egger and MR PRESSO [25], HC algorithm incorporating genetic anchors [23] (based on genetic risk score or the most significant SNP) and MRPC algorithm [24] (based on genetic risk score or the most significant SNP). BIMMER can be implemented using GWAS summary data whereas other seven methods need individual genetic and phenotypic data. To access the performance of algorithm, we calculate the mean of F1 score, recall, precision and computing time across 100 data sets with 10,000 individuals for each method. Recall (i.e., power, or sensitivity) measures how many edges from the true causal graph a method can recover, whereas precision (i.e., 1-FDR) measures how many correct edges are recovered in the inferred graph, and F1 score is a combined index of recall and precision. Details of calculation formula are shown in Supplementary Notes.

### Simulation study 3 on MRSL with fixed graphs

In order to evaluate the performance of MRSL in practical application, we choose three representative examples (Figure 3 A-C): A) MAGIC-NIAB, a network based on the Multiparent Advanced Generation Inter-Cross (MAGIC) winter wheat population produced by the UK National Institute of Agricultural Botany (NIAB), seven continuous traits were measured: yield (YLD), flowering time (FT), height (HT), yellow rust in the glasshouse (YR.GLASS) and in the field (YR.FIELD), Fusarium (FUS), and mildew (MIL). Such a scheme is designed to produce a mapping population from several generations of intercrossing among eight founders and has the potential to improve quantitative trait loci (QTL) mapping precision. B) ASIA, also called lung cancer network, consists of eight binary variables. Lauritzen and Spiegelhalter (1988) [36] motivate this example as follows: “Shortness-of-breath (dyspnoea) may be due to tuberculosis, lung cancer or bronchitis, or none of them, or more than one of them. A recent visit to Asia increases the chances of tuberculosis, while smoking is known to be a risk factor for both lung cancer and bronchitis. The results of a single chest X-ray do not discriminate between lung cancer and tuberculosis, as neither does the presence or absence of dyspnea.” C) Healthcare Cost, is a recurring theme in most countries’ public discourse due to the combination of an ageing population and the availability of more advanced (read, expensive) treatments. We use the simple example in Marco et al. (2021) [77], which modeled an individual’s yearly medical expenditure by seven mixed variables: age, pre-existing conditions, outpatient expenditure, inpatient expenditure, any hospital stay, days of hospital stay and taxes.

The data generation process and parameters’ settings of these three networks are similar with that in simulation study 2. We also use F1 score, recall, precision and computing time to evaluate the performance of MRSL.

### Applied example

We apply MRSL to learn the network of 26 biomarkers and 44 ICD10-defined diseases using GWAS summary data in UK Biobank. For quantitative phenotypes, we use the phenotypes that have been inverse rank normalized.

For MRSL, we first clumped the UK Biobank summary statistics to p<5×10^−8^ for 26 biomarkers and 44 diseases, with r^2^ < 0.0001 and distance 10000 kilobases using the European reference panel in mrbase (https://www.mrbase.org/). To avoid the selection bias, we choose the IVs in the male population and use the summarized statistics in female population. For step 1 in Figure 1, in order to obtain a marginal causal graph using bi-directional MR, for each MR analysis, we select the SNPs associated with the exposure but not associated with other phenotypes (except exposure and outcome) as IVs. For example, if we perform MVMR *X*_3_ ∽*X*_1_ + *X* _4_, SNPs associated with *X* _1_ but not associated with *X* _4_ are selected as IVs. Next we perform MVMR using three adjustment strategies to obtain the true graph. We select SNPs associated with at least one phenotype of exposure and the adjusting variables as IVs. For example, if we perform MVMR *X*_3_ ∽*X*_1_ + *X* _4_, SNPs associated with at least one of *X* _1_, *X* _3_ and *X* _4_ but not associated with *X* _2_ are selected as IVs. For each MVMR, we also need to filter out the SNPs with linkage disequilibrium (r^2^ > 0.0001).

## Data Availability

All data produced are available online at http://www.nealelab.is/uk-biobank.

## Data and code availability

The GWAS summary data in UK Biobank are publicly available at http://www.nealelab.is/uk-biobank. All the analysis in our article were implemented by R software. MRSL can be implemented by https://github.com/hhoulei/MRSL. BIMMER were implemented using R packages *bimmer*. MRPC were implemented using R packages *MRPC*. HC algorithm were implemented using R packages *bnlearn*. cGAUGE were implemented using functions in https://github.com/david-dd-amar/cGAUGE and R packages *MendelianRandomization, MRPRESSO*. All the networks were plotted using R packages *igraph*.

## Ethics approval and consent to participate

Ethical approval was not sought, because this study involved analysis of publicly available summary-level data from GWASs, and no individual-level data were used.

## Authors’ contributions

HL and FX conceived the study. LH contributed to theoretical derivation with assistance from HL and ZG. LH contributed to the data simulation. LH and CW contributed to the application. LH, ZG, XS and HL wrote the manuscript with input from all other authors. All authors reviewed and approved the final manuscript.

## Acknowledgements

None.

