## Supplementary Notes for "MRSL: A phenome-wide causal discovery algorithm based on GWAS summary data"

**Content**

### Proof of Lemma 1

**Lemma 1.** For the true causal graph 𝒢 and the marginal causal graph 𝒢_M_, E_𝒢_⊆E_𝒢M_ and S_𝒢_⊆S_𝒢M_.

**Proof**: If
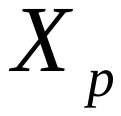
 has a causal relationship with
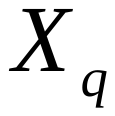
 in the true causal graph 𝒢, whether a direct or an indirect causal relationship, there exists an edge
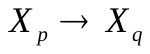
 in the marginal causal graph 𝒢_M_. There is no edge linking
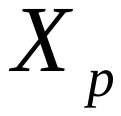
 and
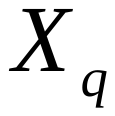
 in 𝒢_M_ if they are not causally related in 𝒢. Thus the collider in 𝒢 is also a collider in 𝒢_M_ but the extra edge may induce spurious collider in 𝒢_M_. Taking the edge
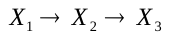
 as an example, the marginal causal graph 𝒢_M_ will add an extra edge
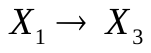
. This extra edge induces a spurious collider
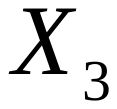
 in 𝒢_M_, that is,
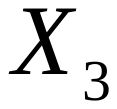
 is not a collider in the graph 𝒢 but is a collider in 𝒢_M_ (
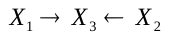
). ∎

### Proof of Lemma 2

**Lemma 2 (Topological sorting invariance).** The topological sorting of the true causal graph 𝒢 and the marginal causal graph 𝒢_M_ are the same *T*_𝒢_*=T*_𝒢M_.

**Proof**: Based on Lemma 1, comparing with the true causal graph 𝒢, the marginal causal graph 𝒢_M_ may add several extra edges and induce spurious colliders but this cannot change the topological sorting. For a pair of nodes
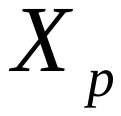
 and
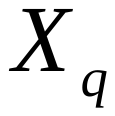
, if
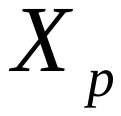
 is ordered before
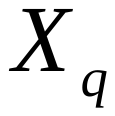
 in the graph 𝒢,
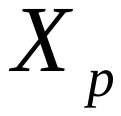
 is a parent node of
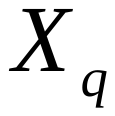
, that is, there is a directed path from
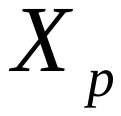
 to
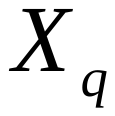
. However, if the topological sorting changes in the graph 𝒢_M_,
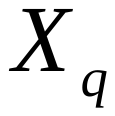
 is ordered before
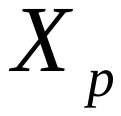
, that is,
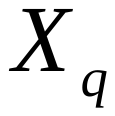
 is a parent node of
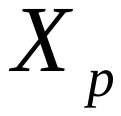
 and a cycle appears. ∎

### Proof of Theorem 1

**Theorem 1:** Under the Assumptions 2-4, for each edge
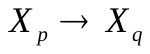
 in the marginal causal graph 𝒢_M_, given a sufficient separating set
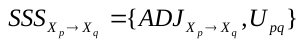
 such that
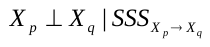
, which can be tested by adjusting for genetic associations with
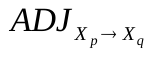
 using MVMR,

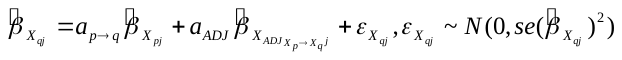
,

where
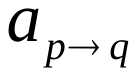
 determines the existence of edge from
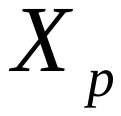
 to
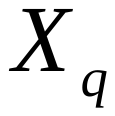
 in the true causal graph 𝒢, then there is no direct edge from
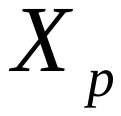
 to
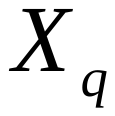
 in the true causal graph 𝒢.

**Proof**: Firstly, given a number of valid IVs satisfying Assumption 4, MVMR overcomes the influence of unobserved confounders on the causal estimation, and estimates the direct effect of
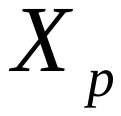
 on
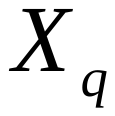
 after adjusting for the genetic associations with
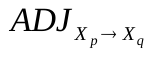
. Taking two continuous phenotypes as an example, the linear model of
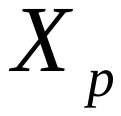
,
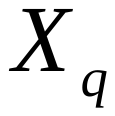
 and
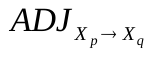
 is

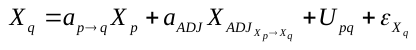
,

where
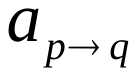
 is the direct causal effect of
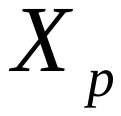
 on
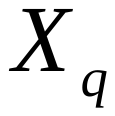
 after adjusting for
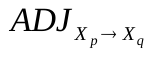
 and U. Then we have

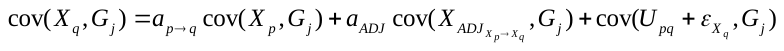
.

If
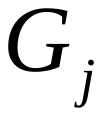
 is a valid IV satisfying Assumption 4,
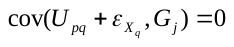
. Divide both sides of this equation by
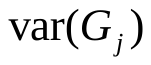
,

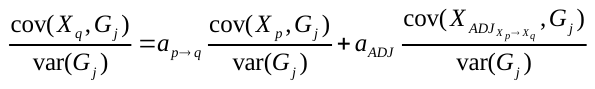
,

which can be simplified to

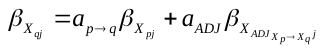
.

Thus the direct effect of
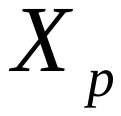
 on
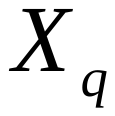
 can be estimated by adjusting for the genetic associations with
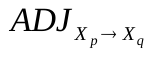
 using MVMR (model (3)). For two nodes
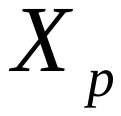
 and
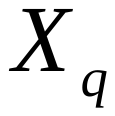
, there is a sufficient separating set
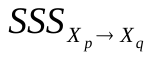
 satisfying
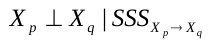
, then
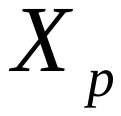
 and
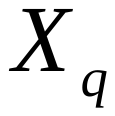
 are not directly linked in graph 𝒢_M_. If directed edge
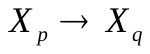
 exists in the true causal graph 𝒢, E_𝒢_ is no more a subset of E_𝒢M_, which is contrary to Lemma 1. ∎

### Data generation of simulation study 1.

| Figures | Data generation | Parameters’ setting |
| --- | --- | --- |
| Figure 2 A) | 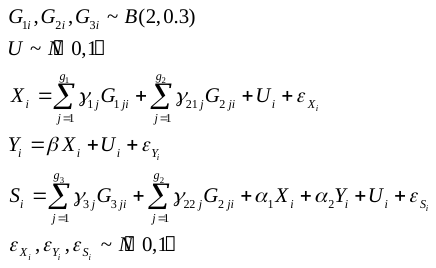 | *i*: *i*-th individual  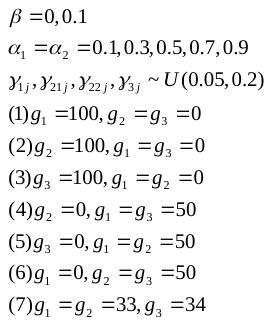 |
| Figure 2 B) | 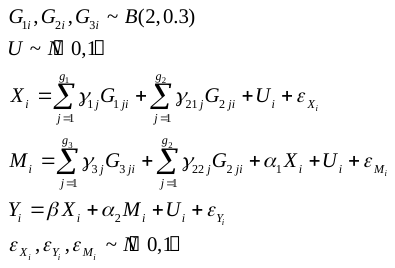 |  |
| Figure 2 C) | 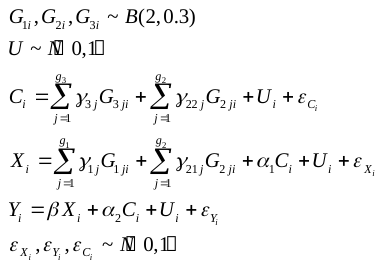 |  |

### Calculation formula of F1score, recall and precision

|  | True | | | |
| --- | --- | --- | --- | --- |
| Predict |  | True | False | All |
|  | Positive | TP | FP | P |
|  | Negative | FN | TN | N |
|  | All | T | F | P+N=T+F |

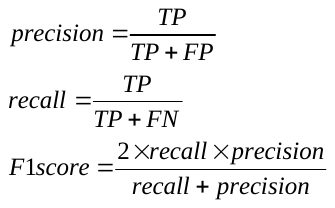

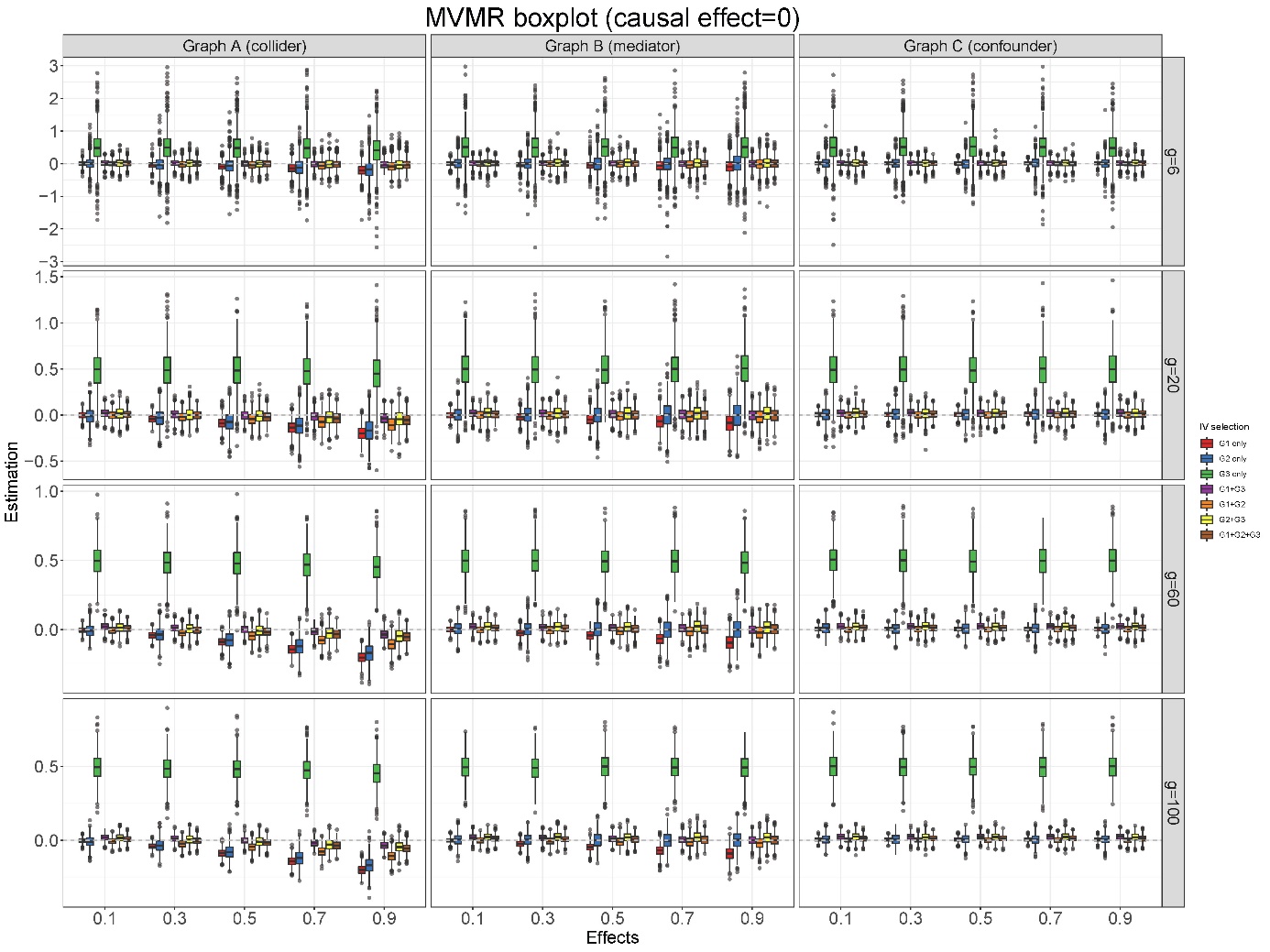

### Supplementary Figure 1. Estimation of causal effect using MVMR in simulation study 1 (causal effect=0)

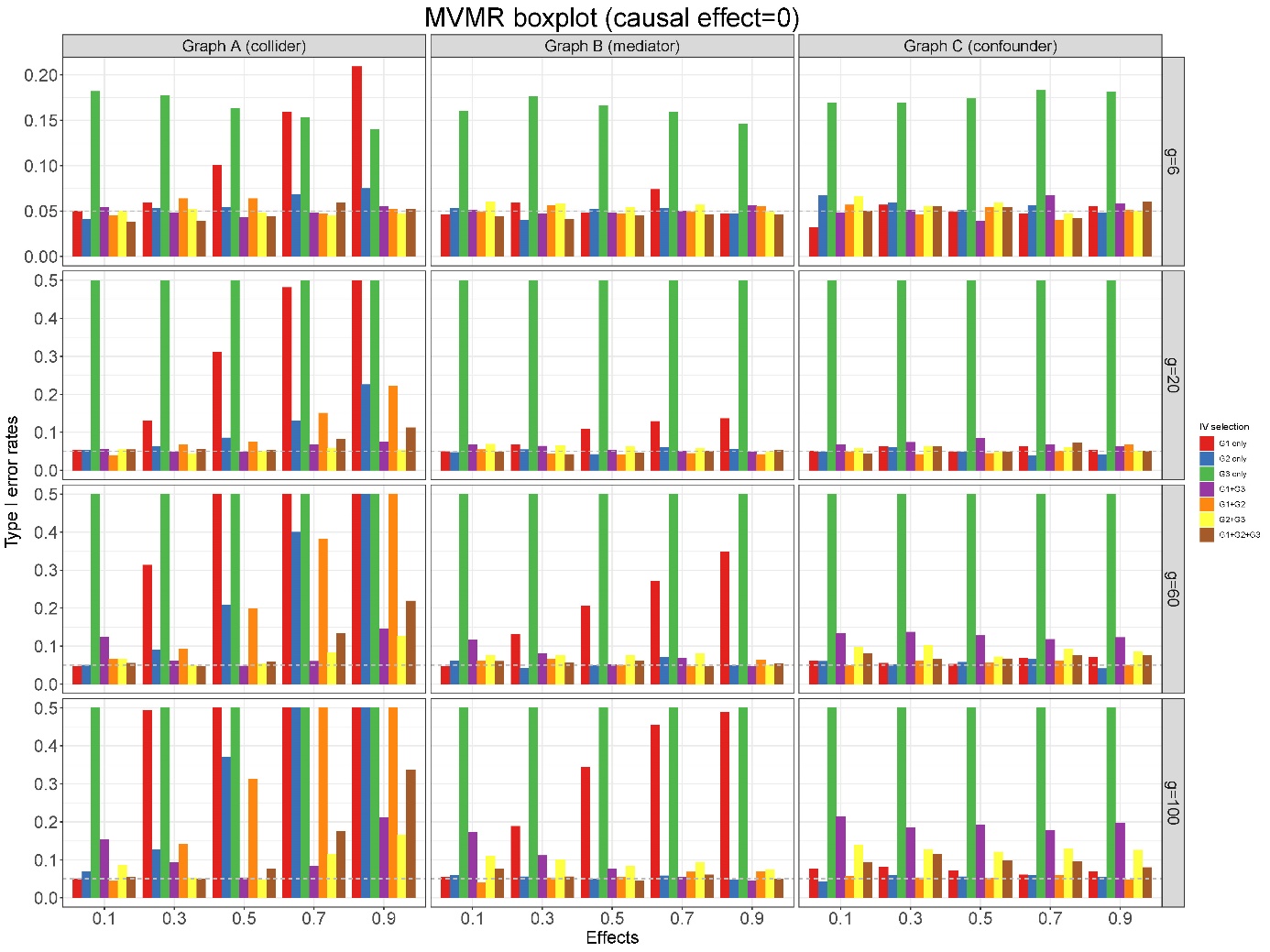

### Supplementary Figure 2. Type I error rate of causal effect using MVMR in simulation study 1 (causal effect=0)

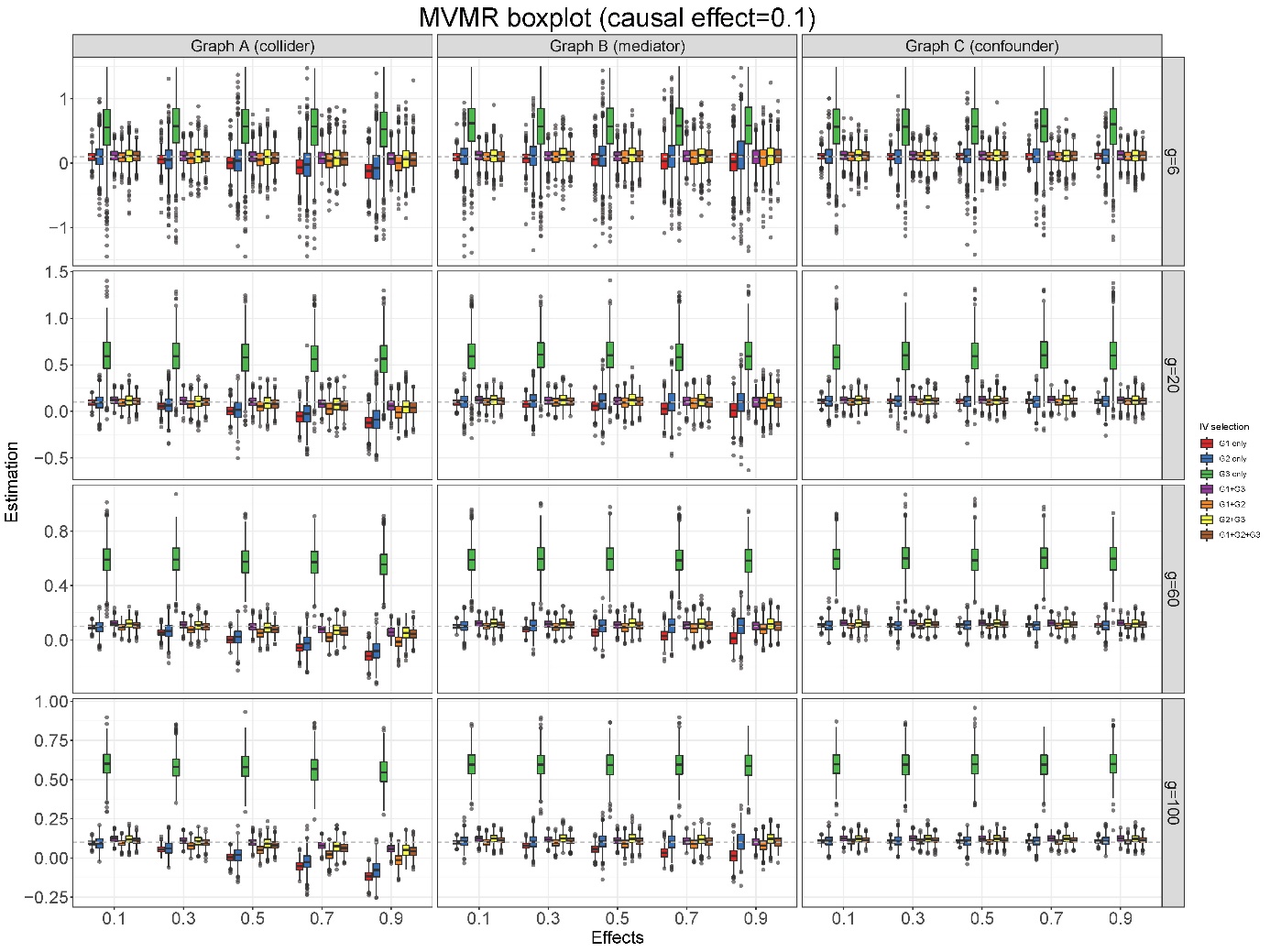

### Supplementary Figure 3. Estimation of causal effect using MVMR in simulation study 1 (causal effect=0.1)

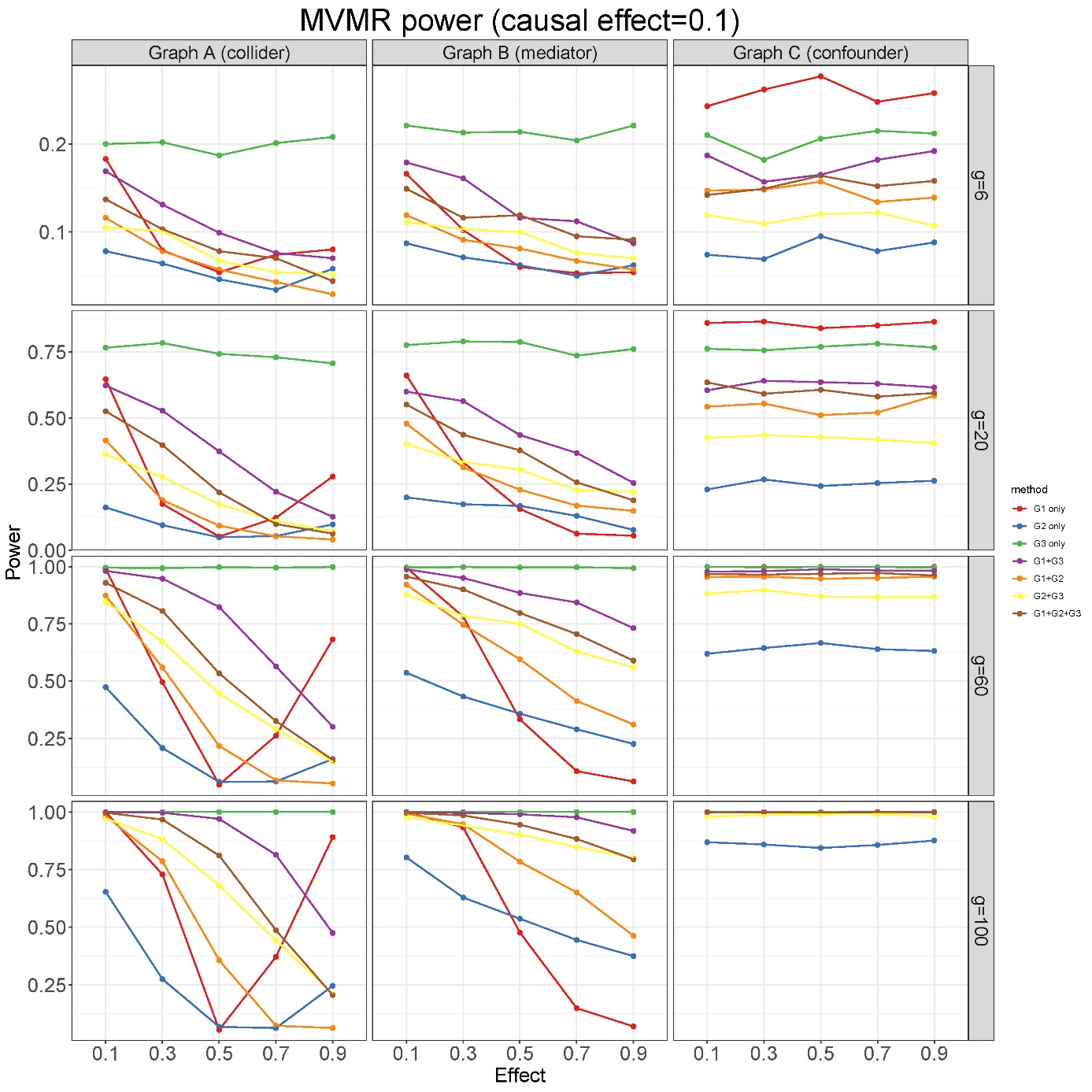

### Supplementary Figure 4. Statistical power of causal effect using MVMR in simulation study 1 (causal effect=0.1)

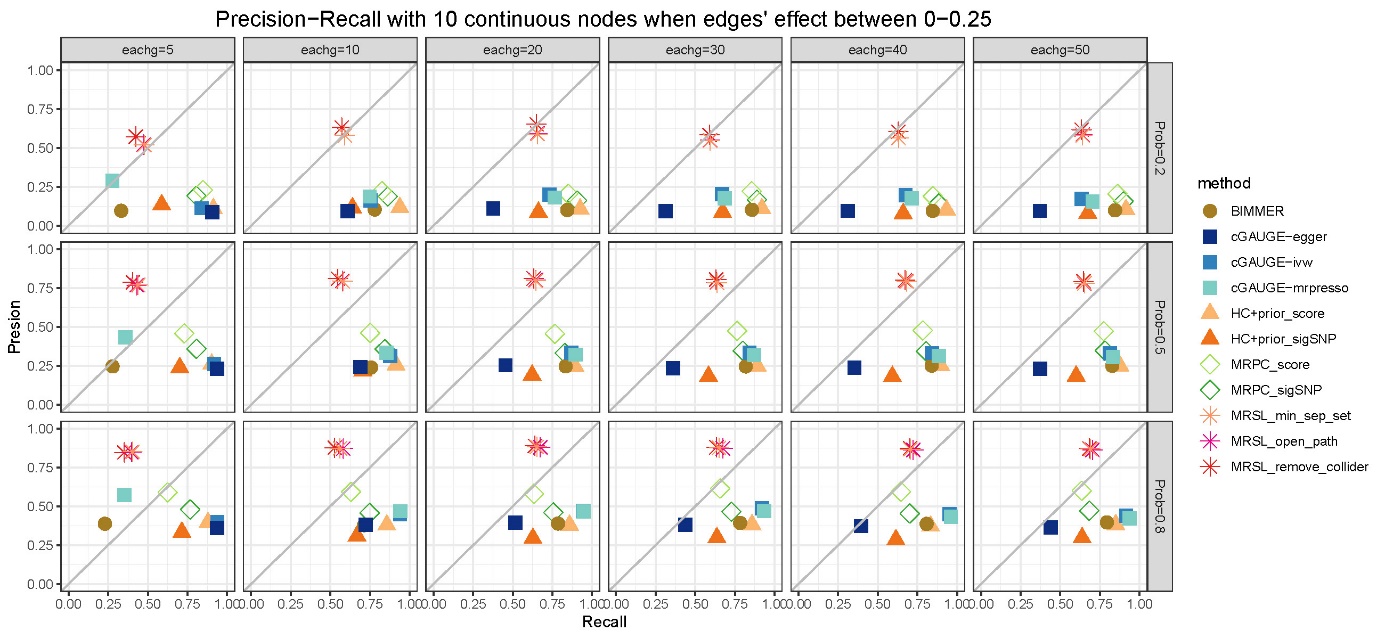

### Supplementary Figure 5. Precision−Recall with 10 continuous nodes when edges' effect between 0−0.25 in simulation study 2

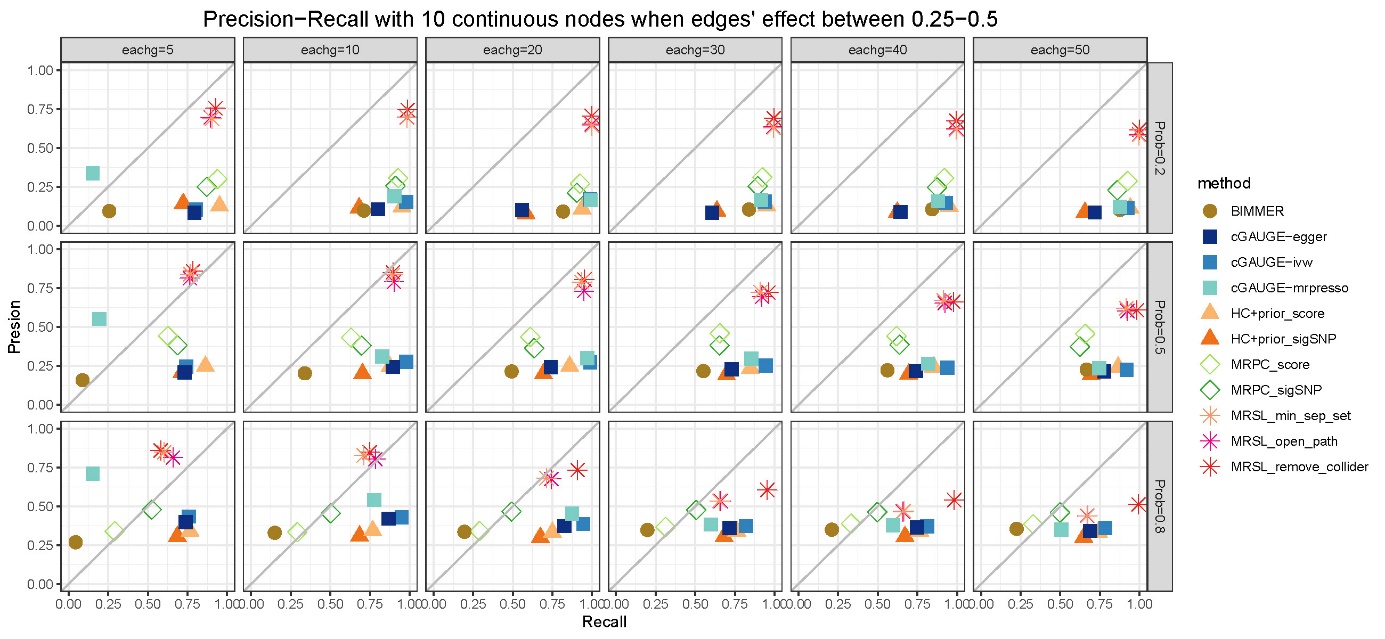

### Supplementary Figure 6. Precision−Recall with 10 continuous nodes when edges' effect between 0.25−0.5 in simulation study 2

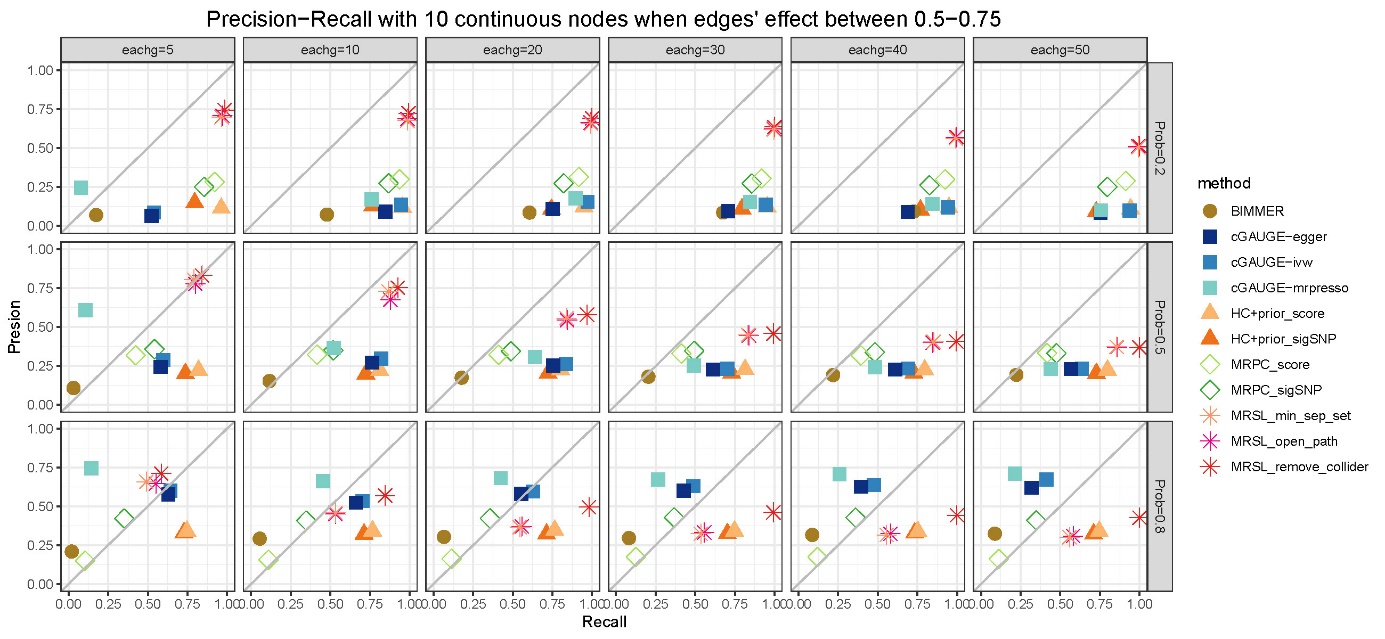

### Supplementary Figure 7. Precision−Recall with 10 continuous nodes when edges' effect between 0.5−0.75 in simulation study 2

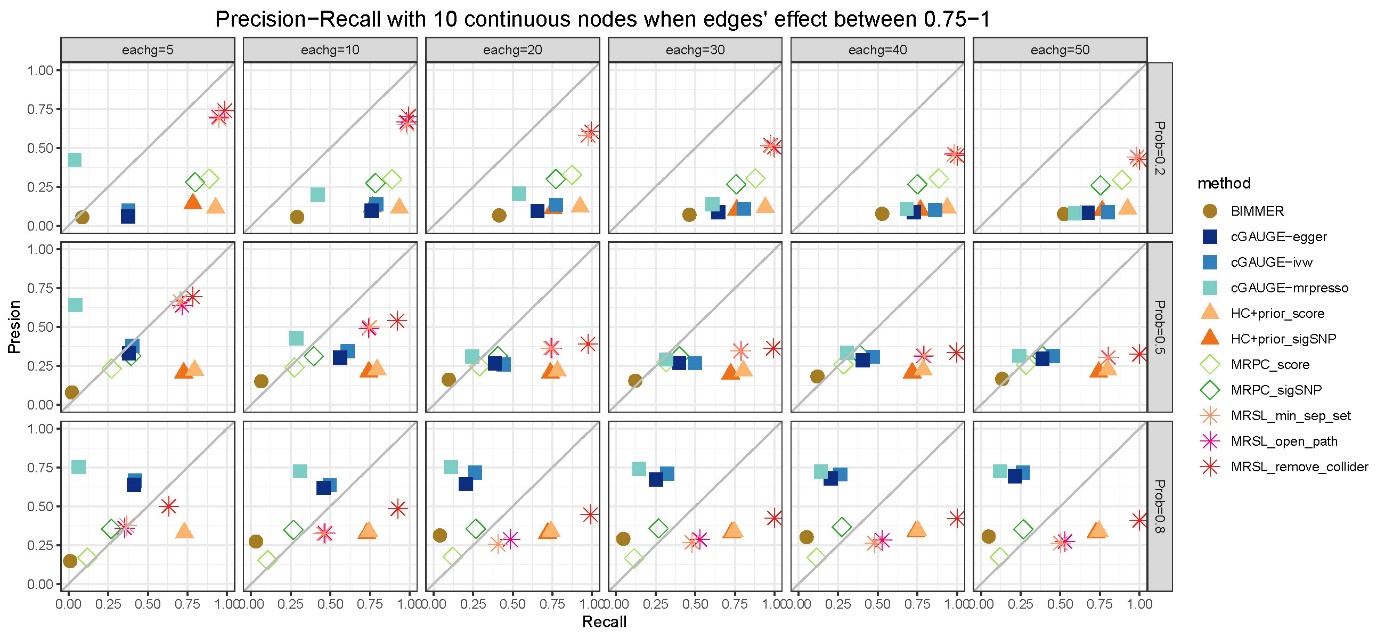

### Supplementary Figure 8. Precision−Recall with 10 continuous nodes when edges' effect between 0.75−1 in simulation study 2

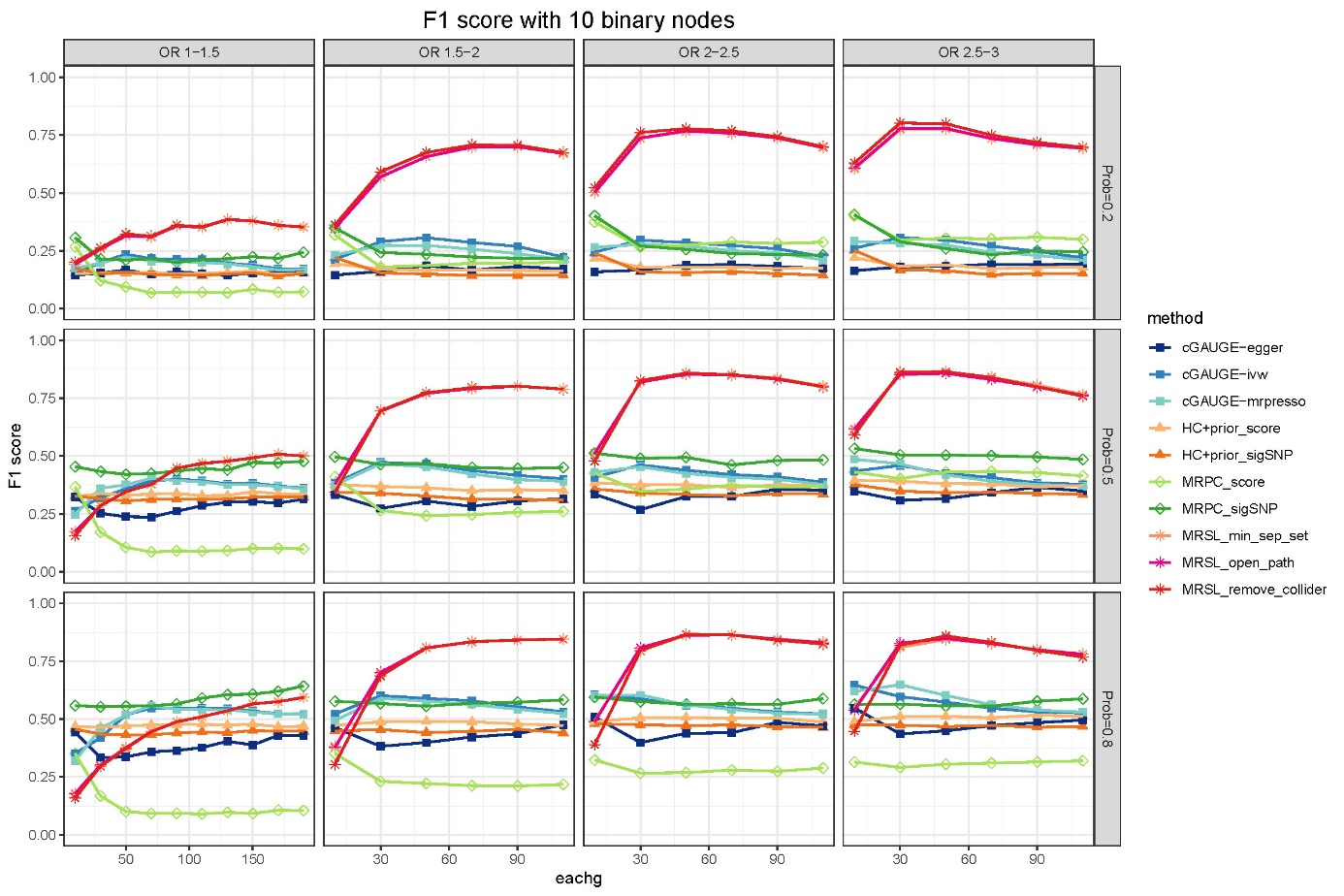

### Supplementary Figure 9. F1 score with 10 binary nodes

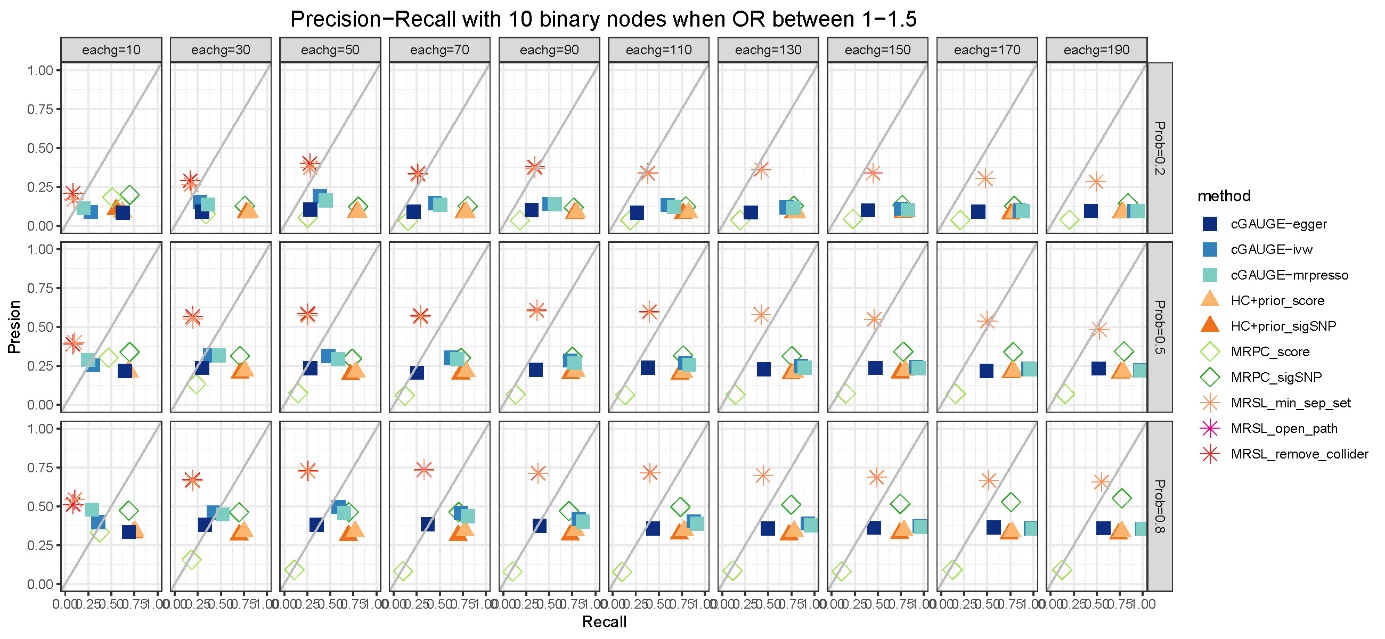

### Supplementary Figure 10. Precision−Recall with 10 binary nodes when OR between 1−1.5 in simulation study 2

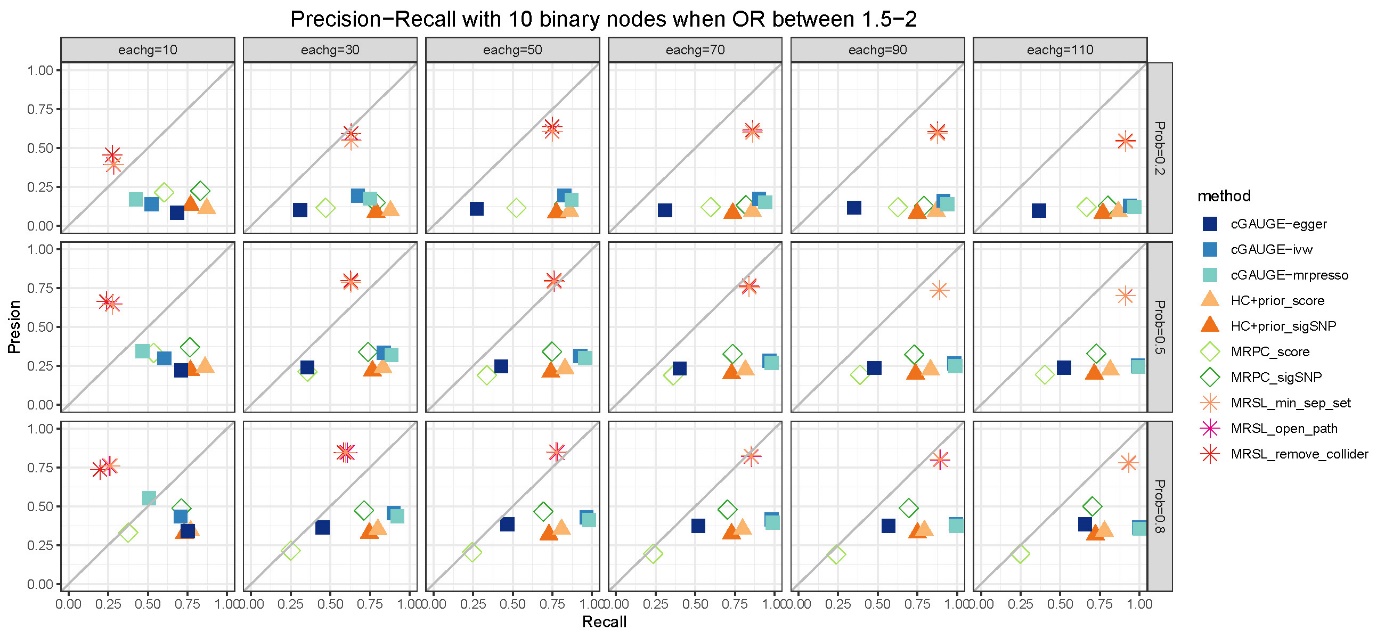

### Supplementary Figure 11. Precision−Recall with 10 binary nodes when OR between 1.5−2 in simulation study 2

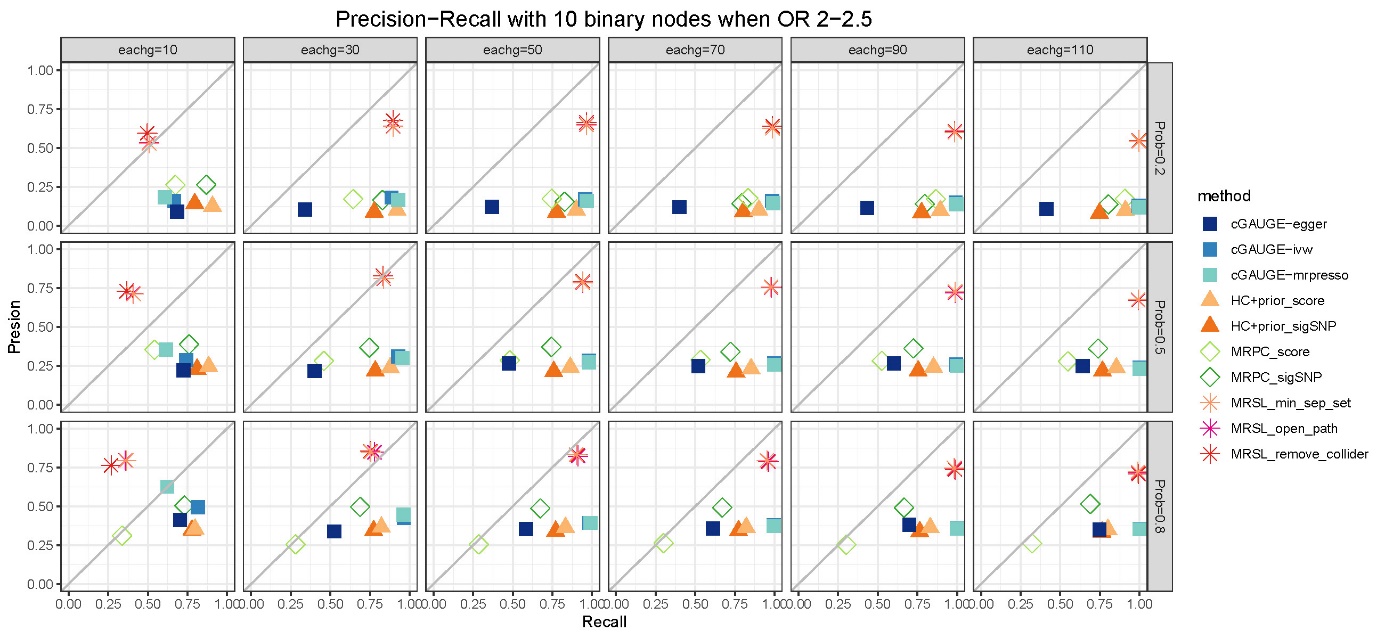

### Supplementary Figure 12. Precision−Recall with 10 binary nodes when OR between 2−2.5 in simulation study 2

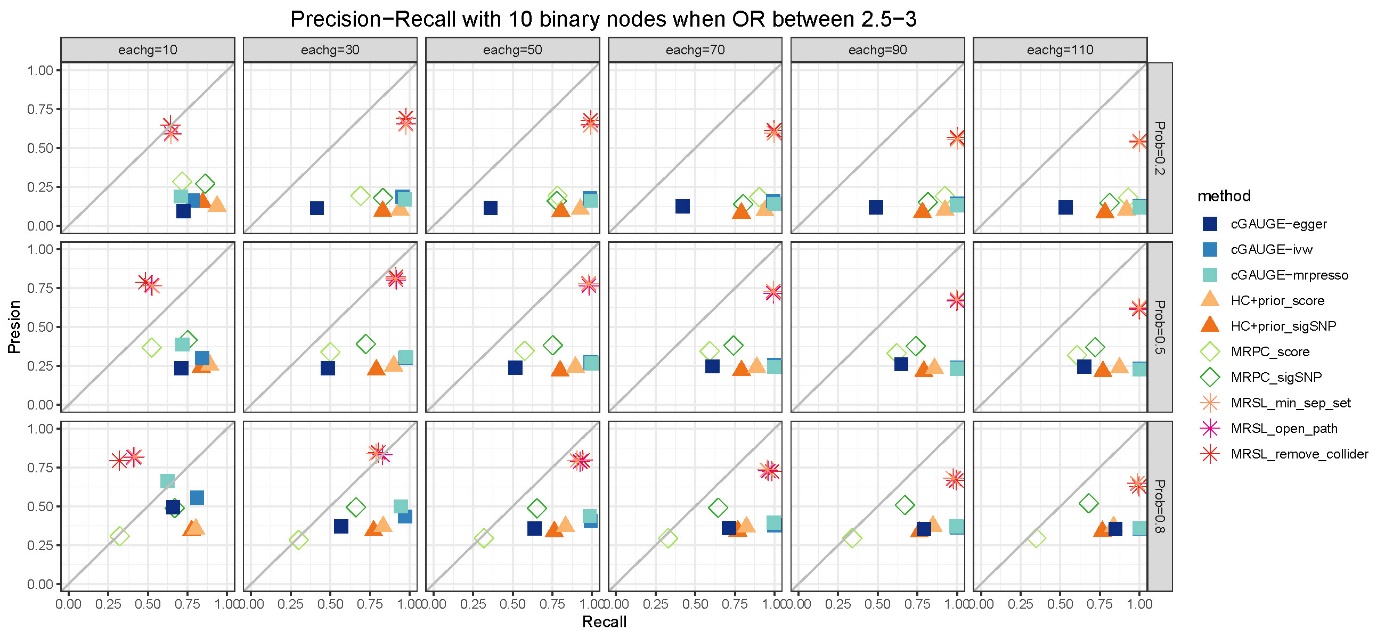

### Supplementary Figure 13. Precision−Recall with 10 binary nodes when OR between 2.5−3 in simulation study 2

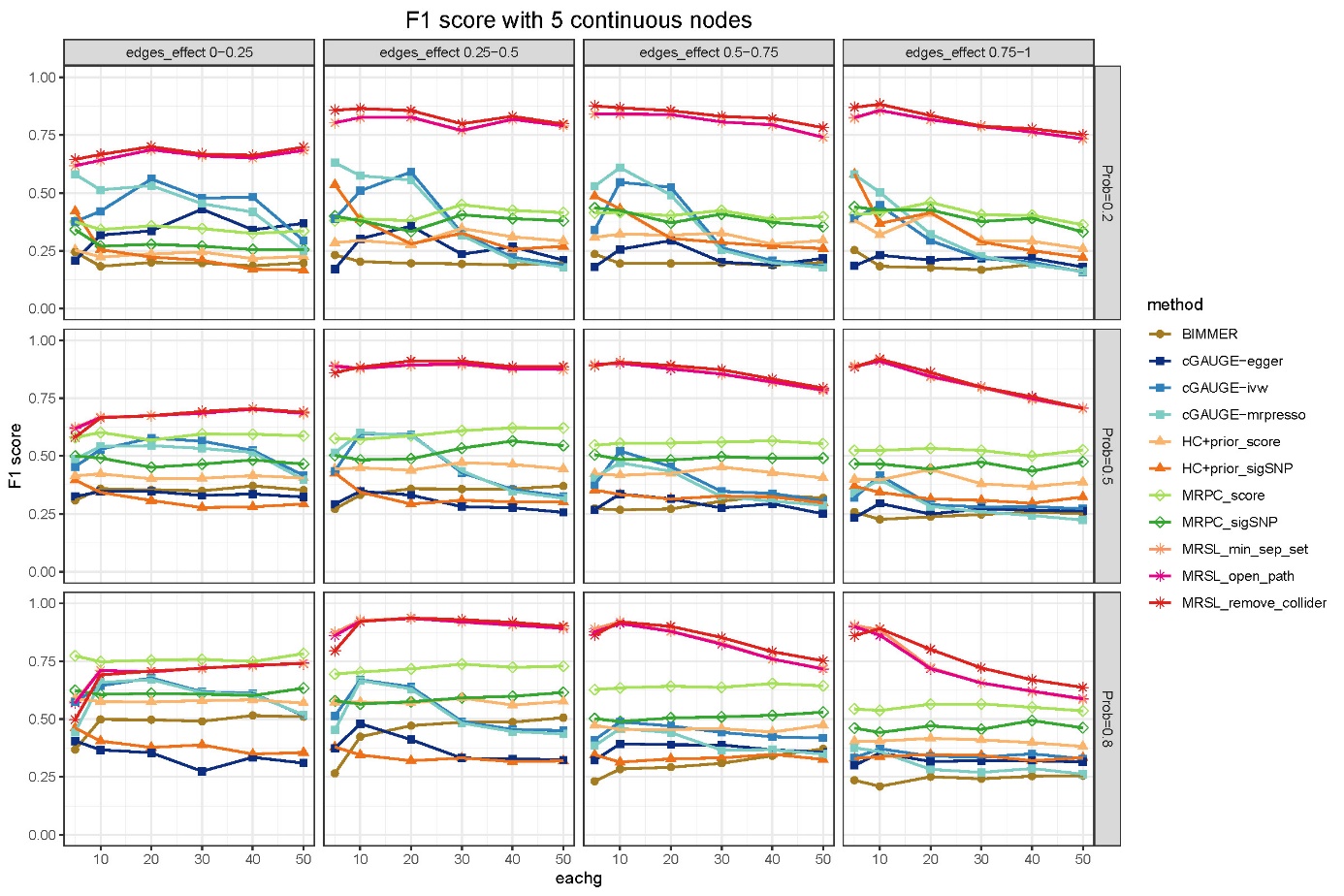

### Supplementary Figure 14. F1 score with 5 continuous nodes in simulation study 2

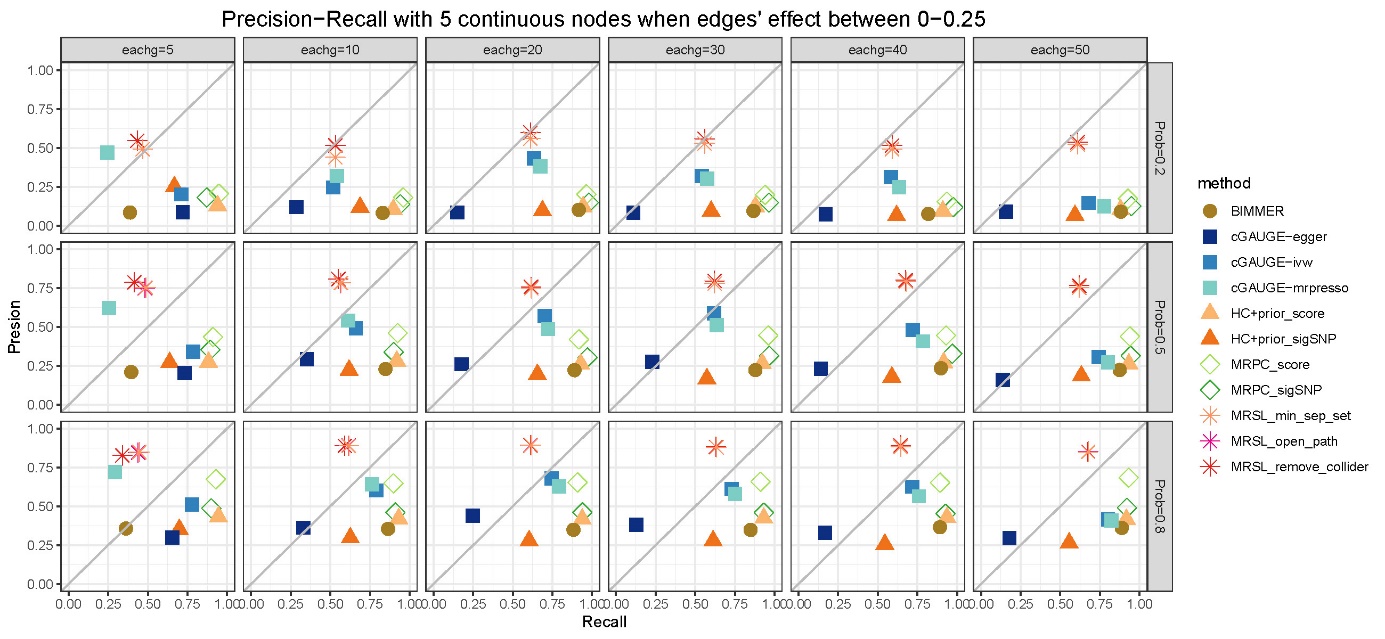

### Supplementary Figure 15. Precision−Recall with 5 continuous nodes when edges' effect between 0−0.25 in simulation study 2

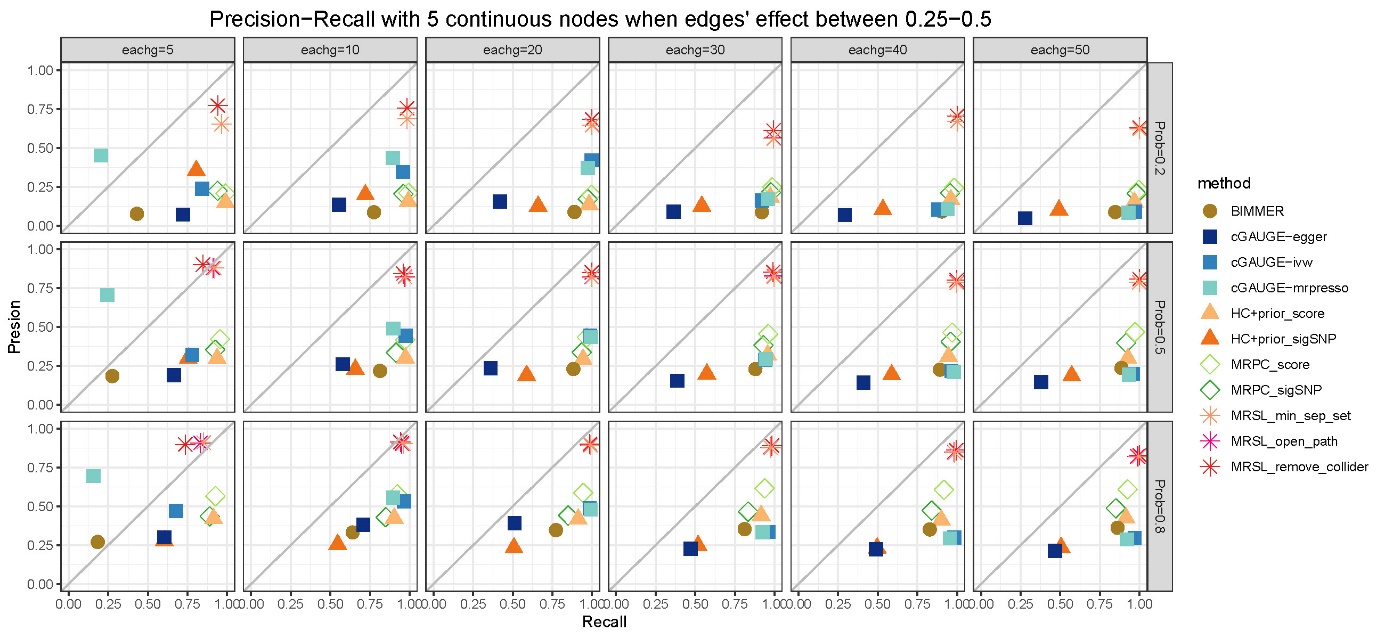

### Supplementary Figure 16. Precision−Recall with 5 continuous nodes when edges' effect between 0.25−0.5 in simulation study 2

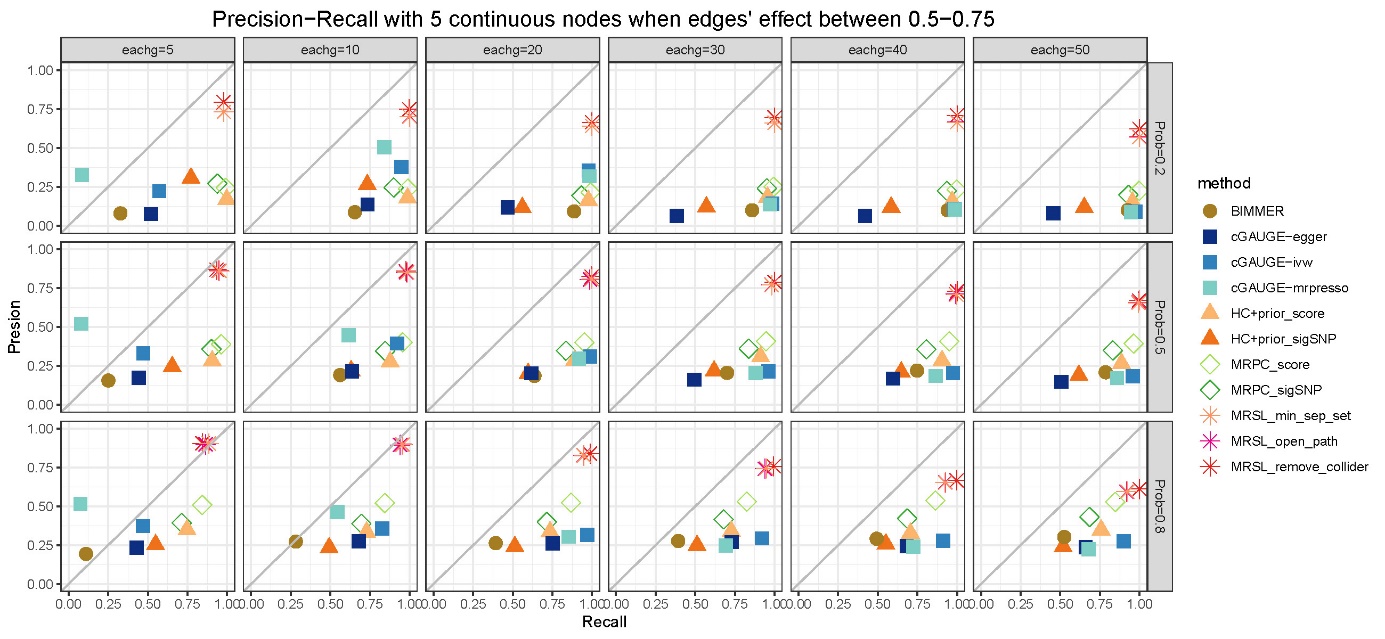

### Supplementary Figure 17. Precision−Recall with 5 continuous nodes when edges' effect between 0.5−0.75 in simulation study 2

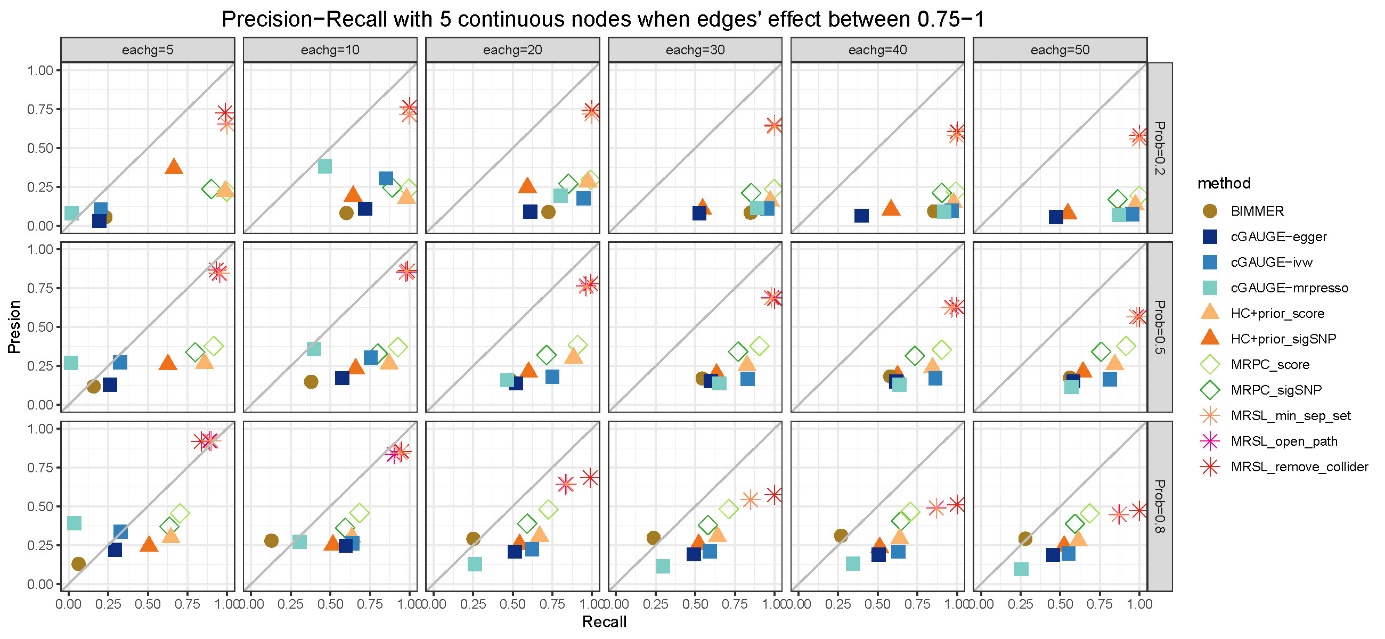

### Supplementary Figure 18. Precision−Recall with 5 continuous nodes when edges' effect between 0.75−1 in simulation study 2

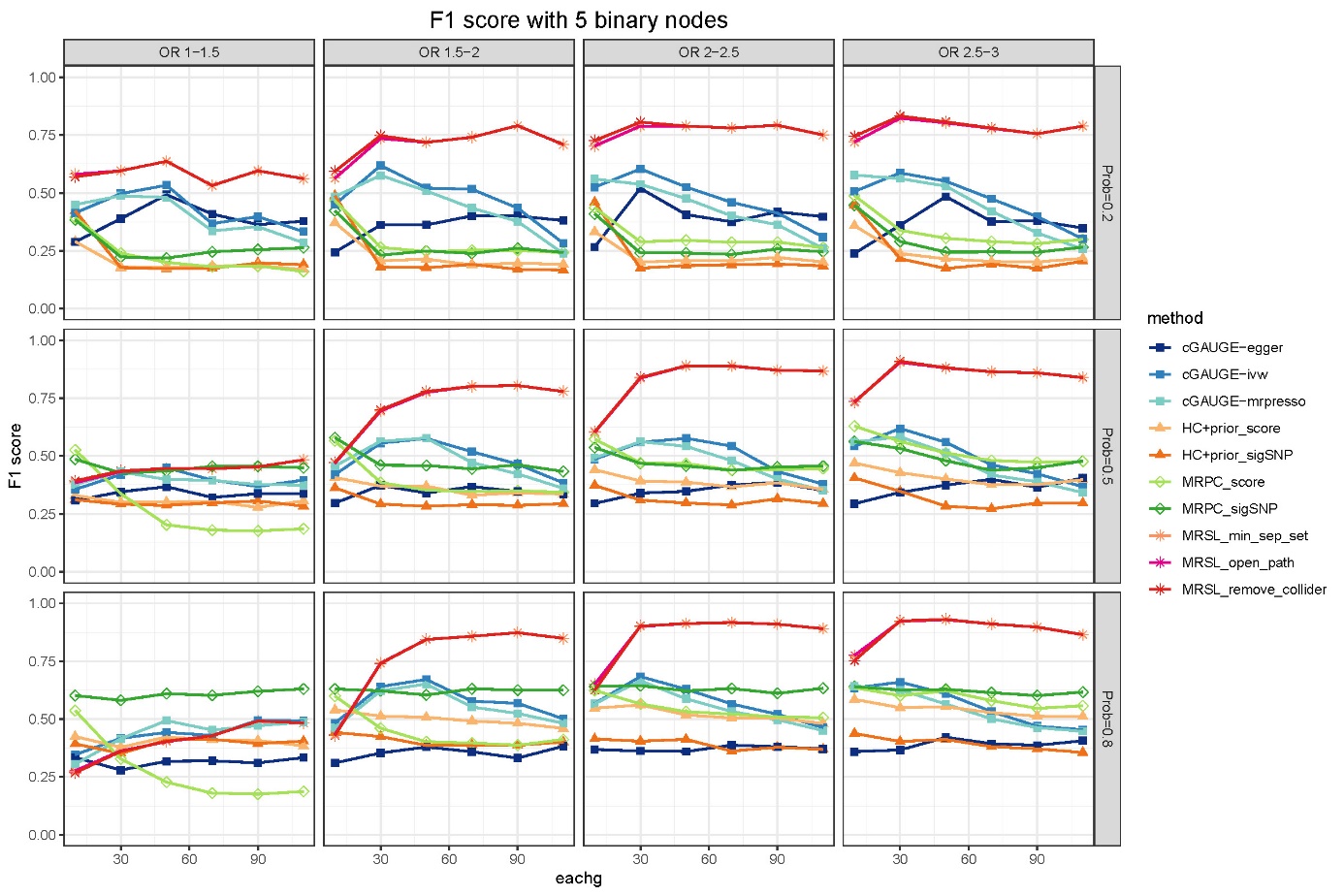

### Supplementary Figure 19. F1 score with 5 binary nodes in simulation study 2

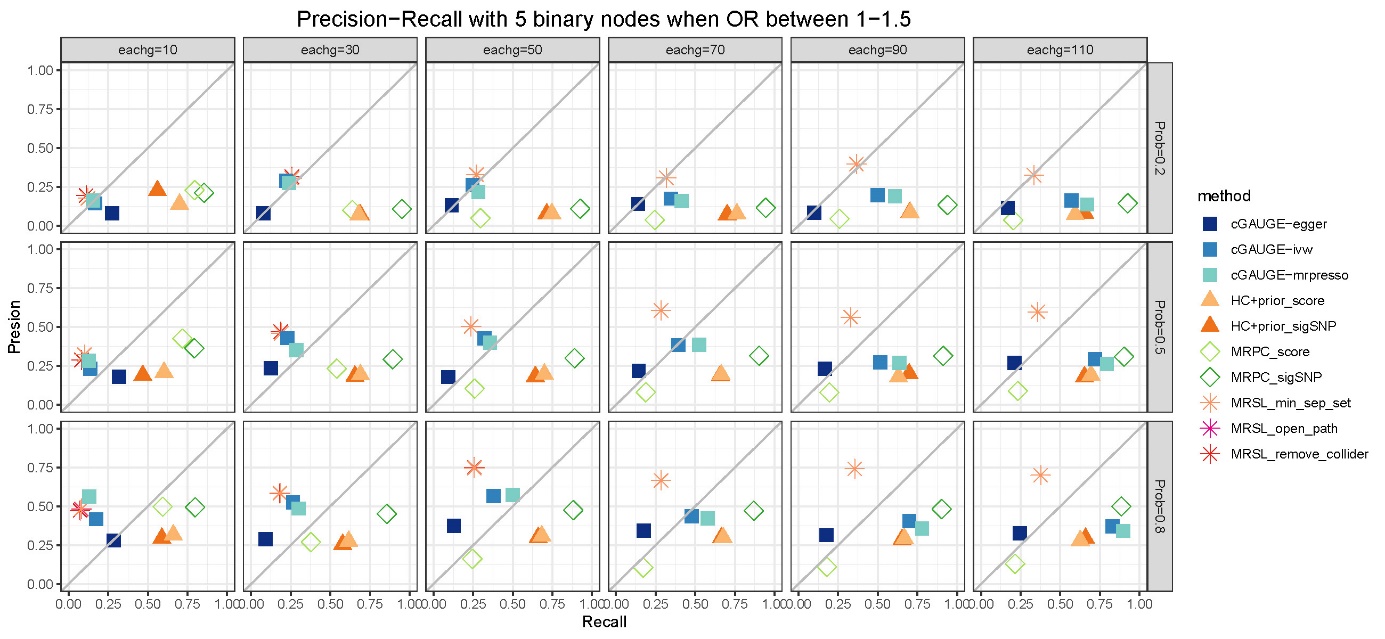

### Supplementary Figure 20. Precision−Recall with 5 binary nodes when OR between 1−1.5 in simulation study 2

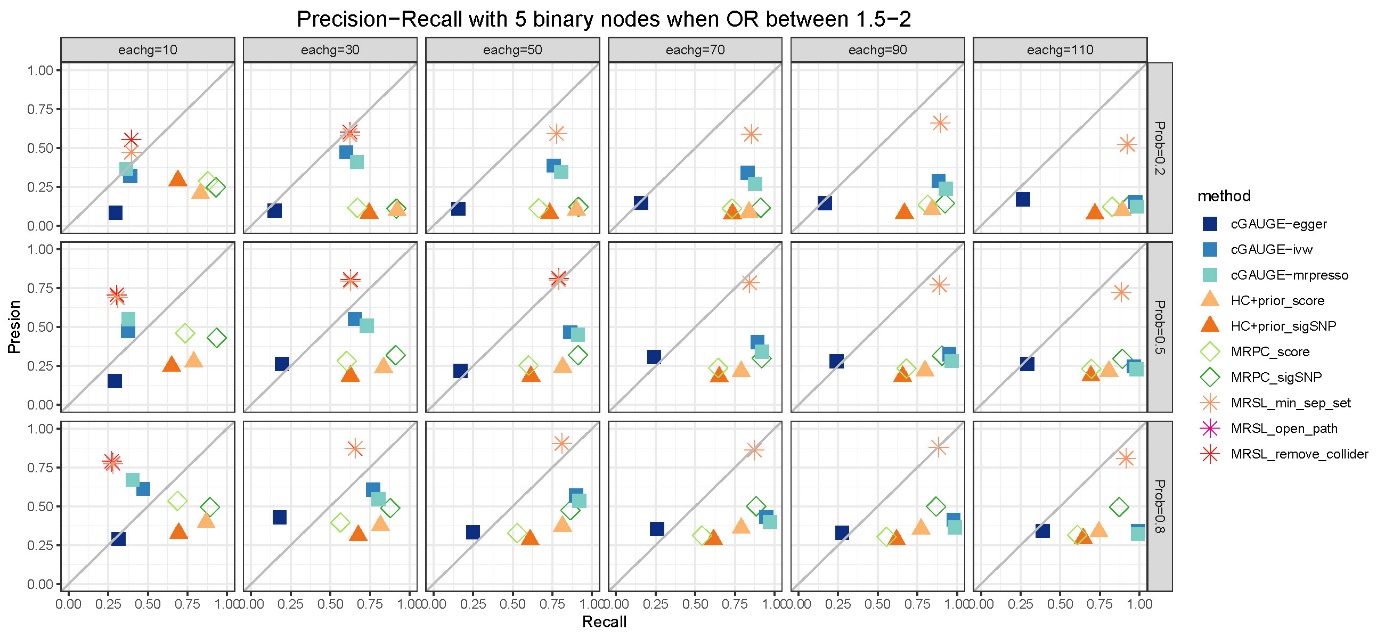

### Supplementary Figure 21. Precision−Recall with 5 binary nodes when OR between 1.5−2 in simulation study 2

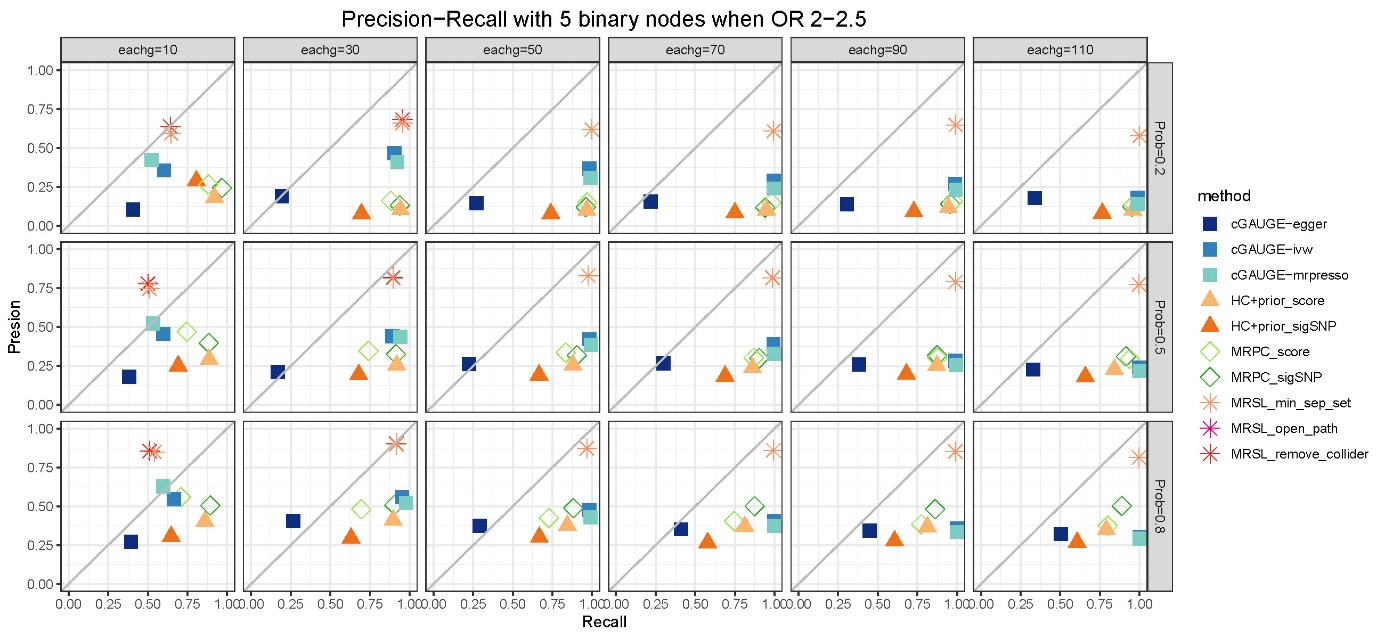

### Supplementary Figure 22. Precision−Recall with 5 binary nodes when OR between 2−2.5 in simulation study 2

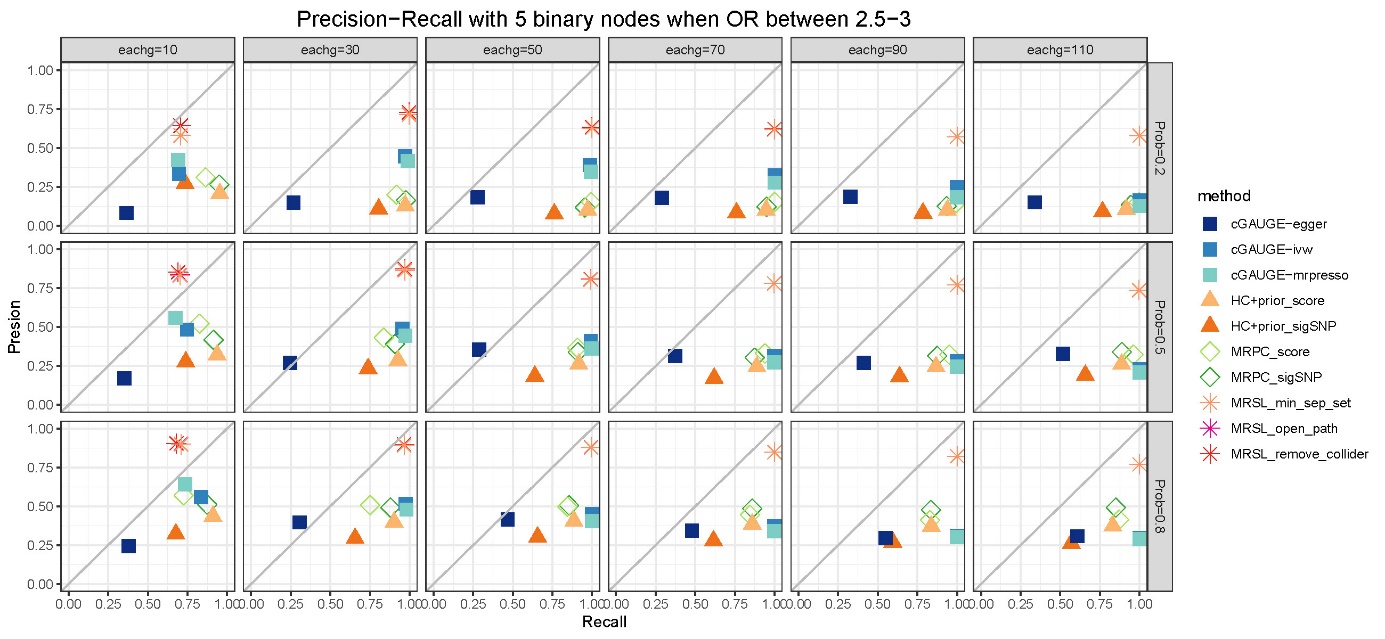

### Supplementary Figure 23. Precision−Recall with 5 binary nodes when OR between 2.5−3 in simulation study 2

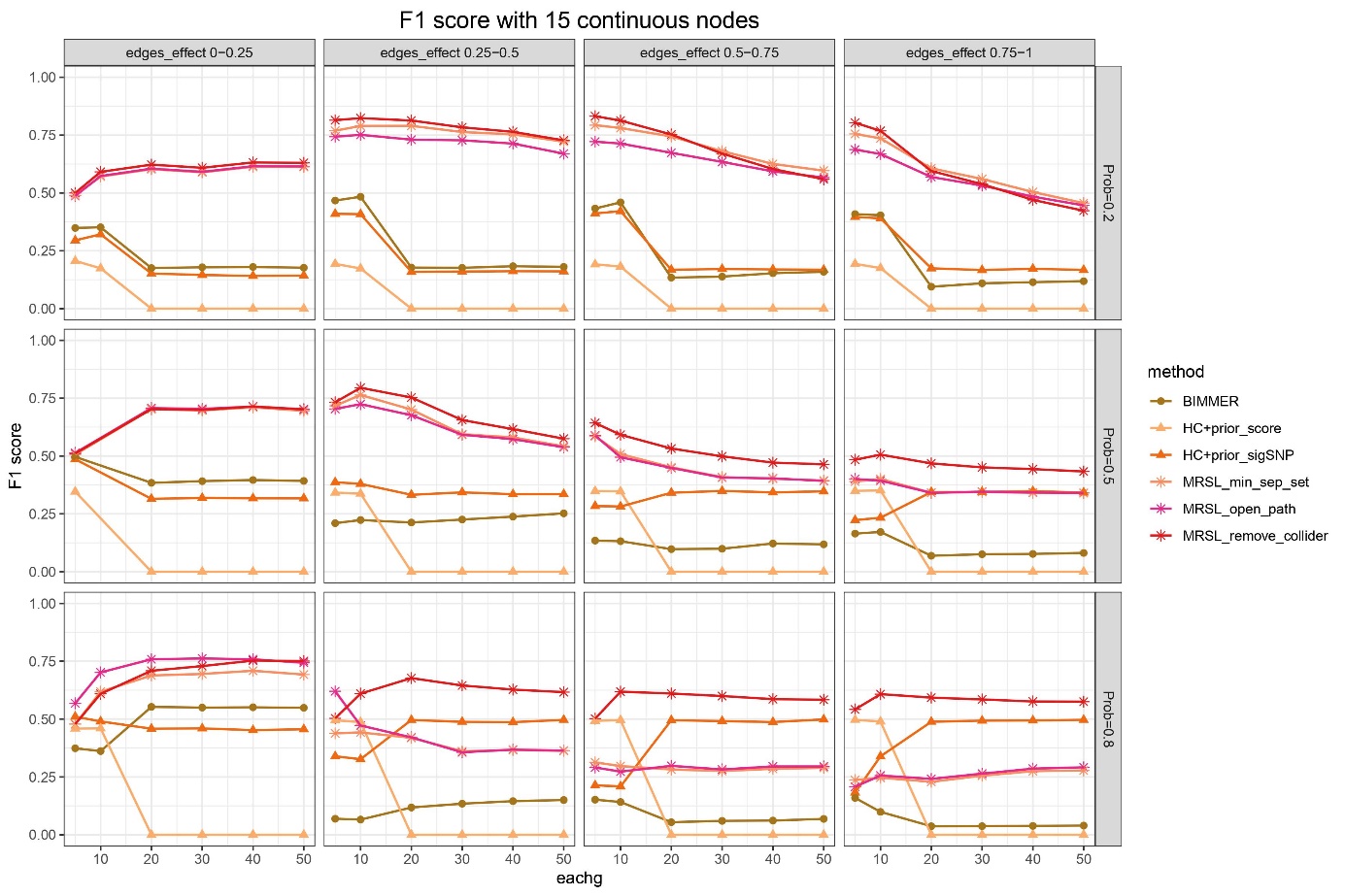

### Supplementary Figure 24. F1 score with 15 continuous nodes in simulation study 2

MRPC and cGAUGE are not listed due to their huge time consuming.

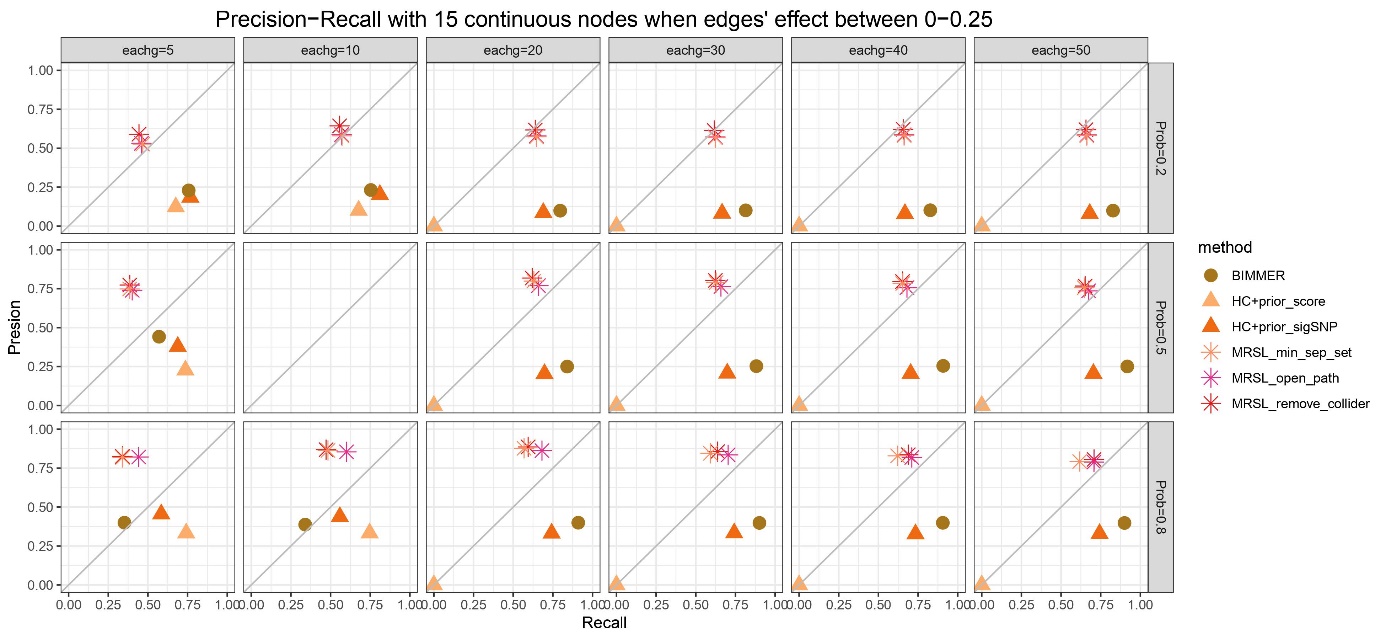

### Supplementary Figure 25. Precision−Recall with 15 continuous nodes when edges' effect between 0−0.25 in simulation study 2

MRPC and cGAUGE are not listed due to their huge time consuming.

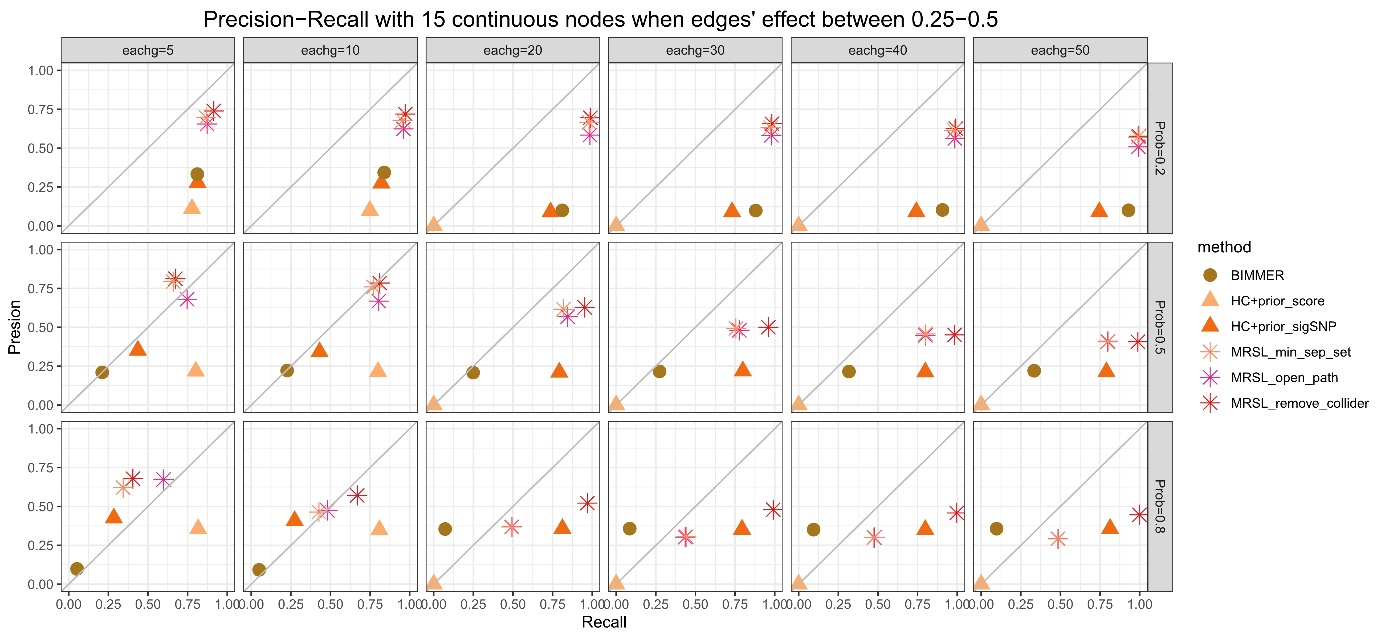

### Supplementary Figure 26. Precision−Recall with 15 continuous nodes when edges' effect between 0.25−0.5 in simulation study 2

MRPC and cGAUGE are not listed due to their huge time consuming.

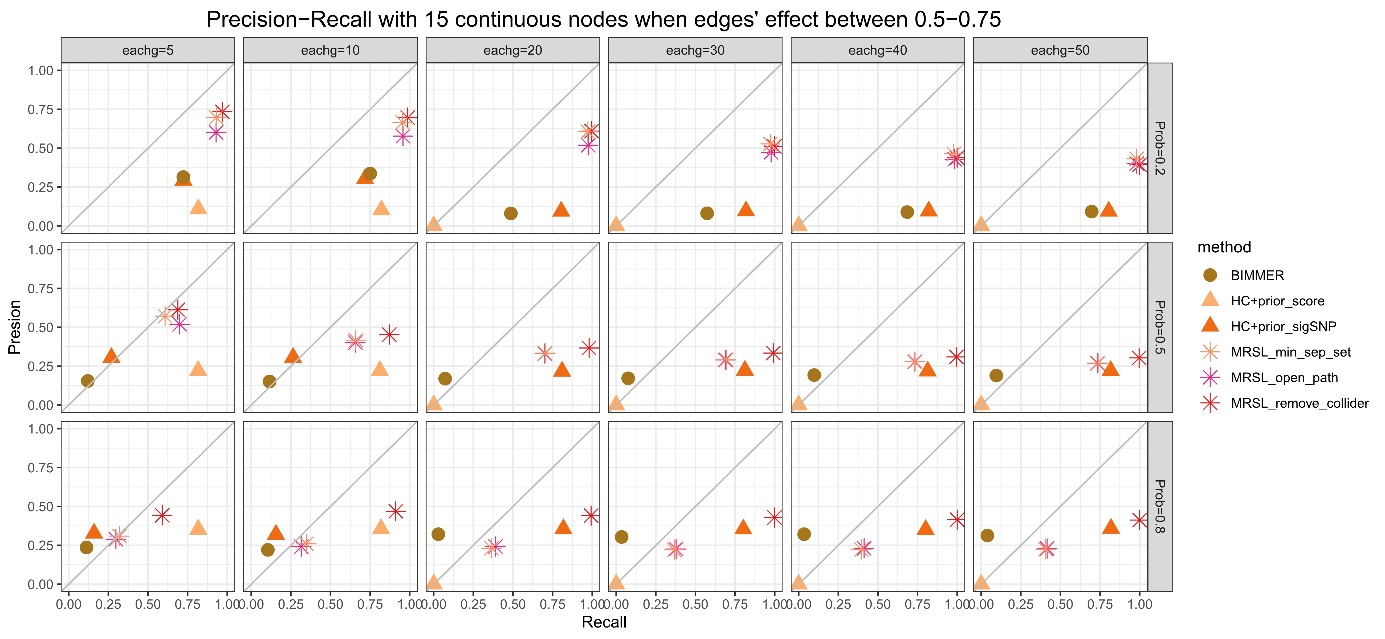

### Supplementary Figure 27. Precision−Recall with 15 continuous nodes when edges' effect between 0.5−0.75 in simulation study 2

MRPC and cGAUGE are not listed due to their huge time consuming.

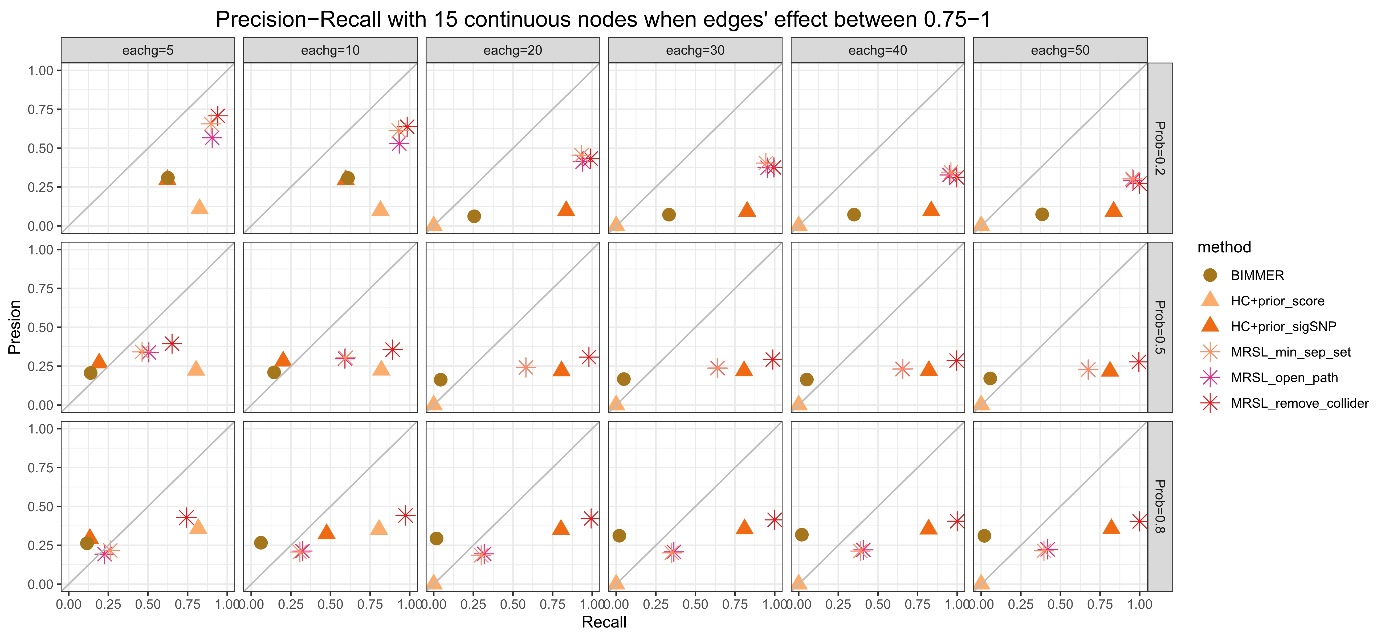

### Supplementary Figure 28. Precision−Recall with 15 continuous nodes when edges' effect between 0.75−1 in simulation study 2

MRPC and cGAUGE are not listed due to their huge time consuming.

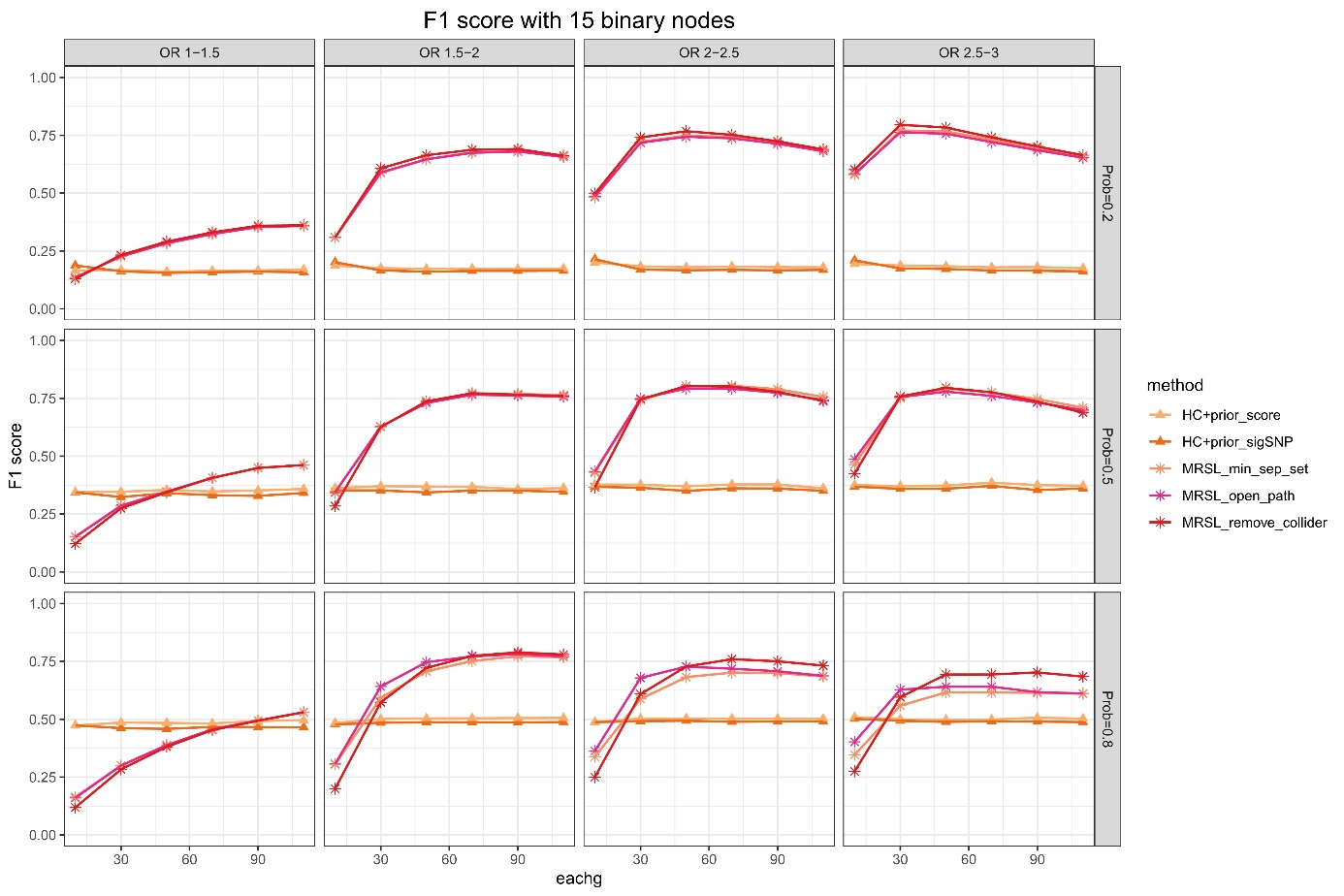

### Supplementary Figure 29. F1 score with 15 binary nodes in simulation study 2

MRPC and cGAUGE are not listed due to their huge time consuming.

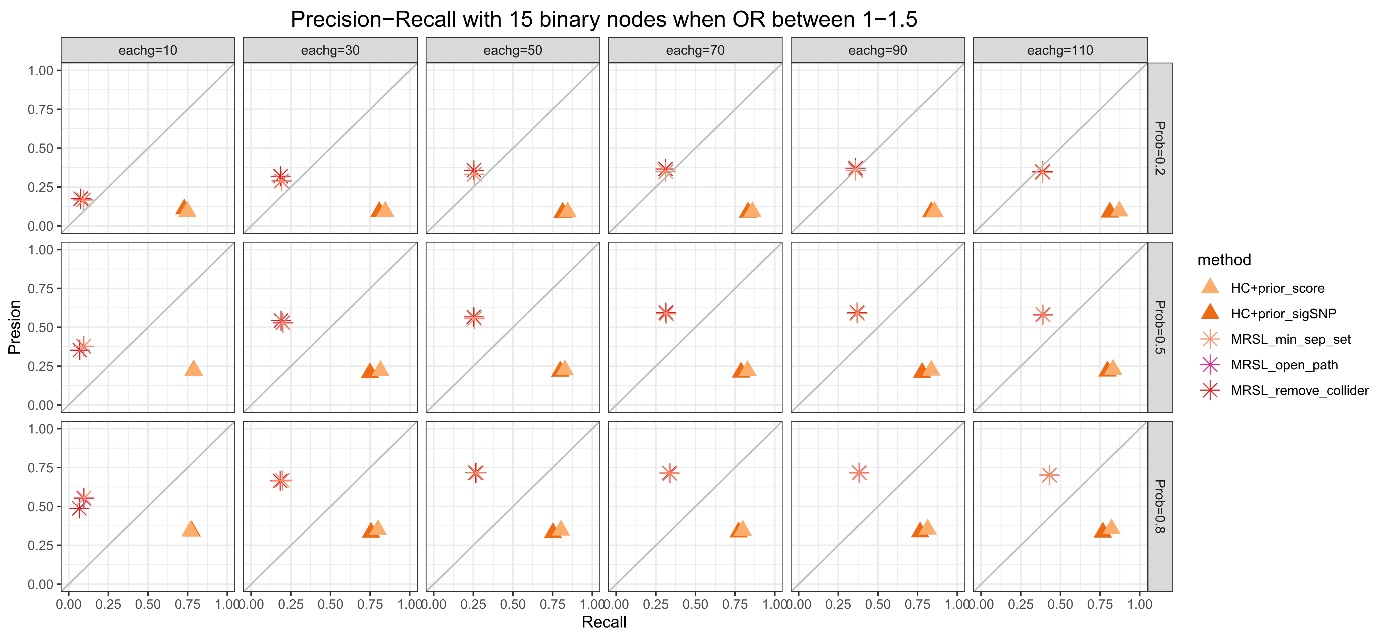

### Supplementary Figure 30. Precision−Recall with 15 binary nodes when OR between 1−1.5 in simulation study 2

MRPC and cGAUGE are not listed due to their huge time consuming.

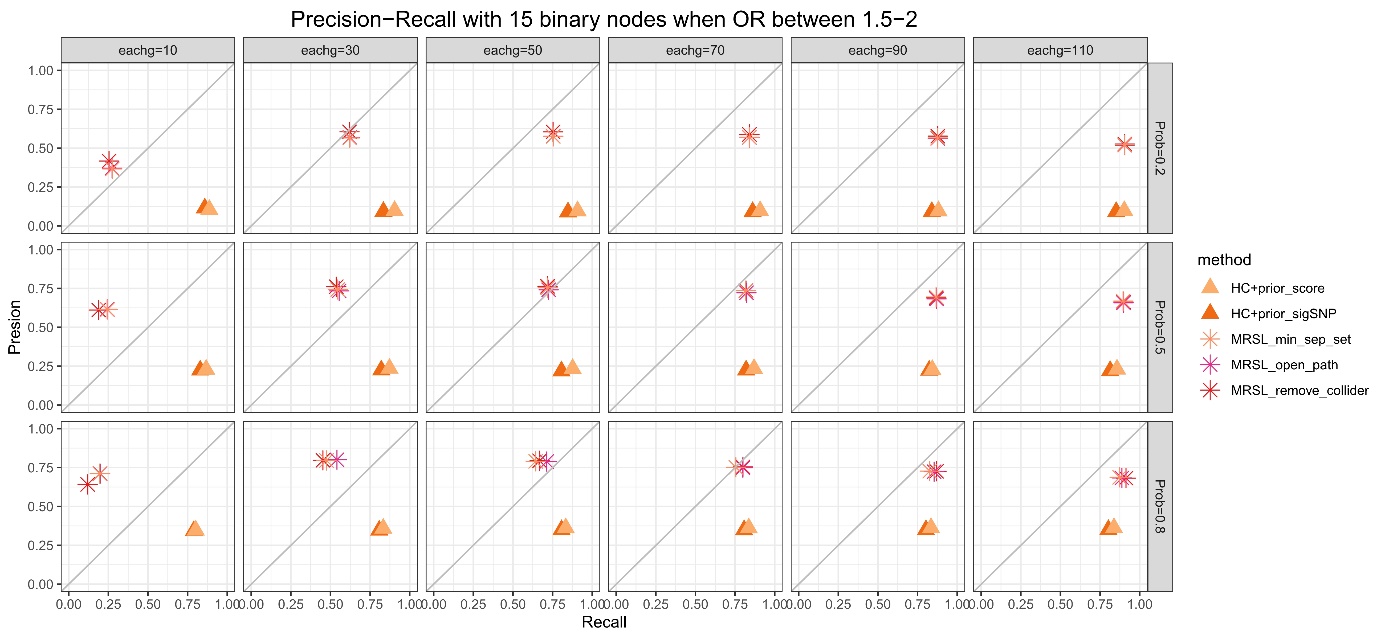

### Supplementary Figure 31. Precision−Recall with 15 binary nodes when OR between 1.5−2 in simulation study 2

MRPC and cGAUGE are not listed due to their huge time consuming.

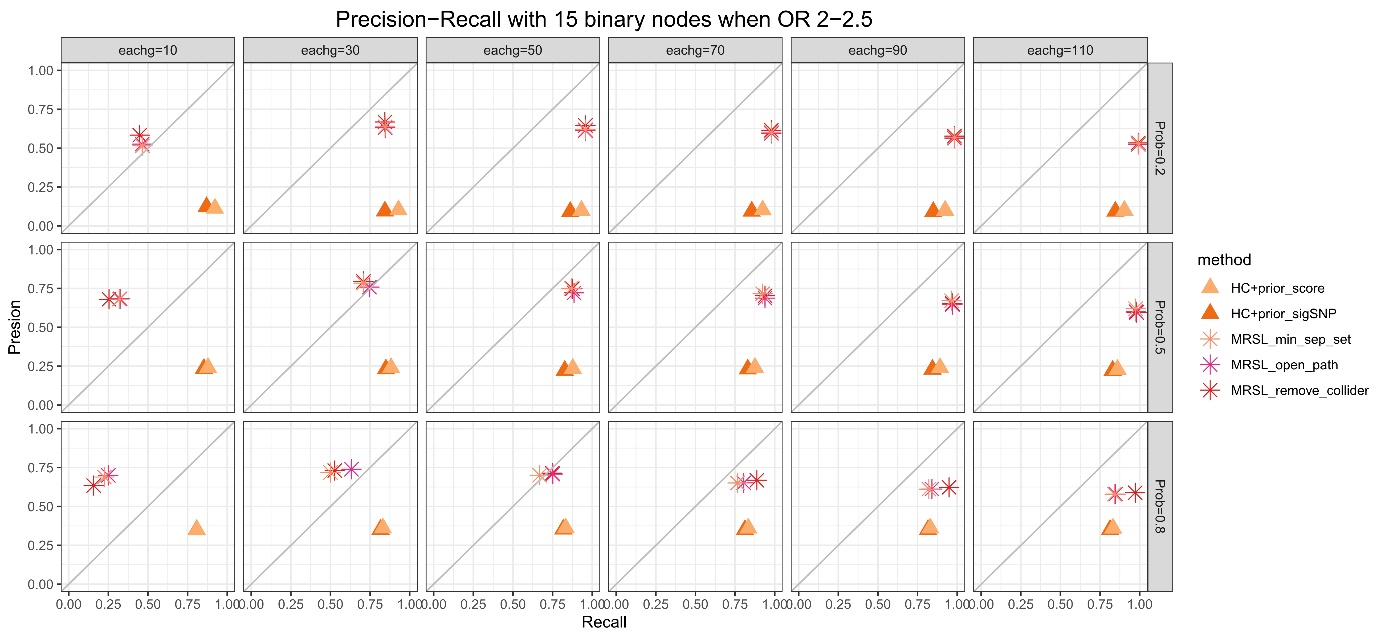

### Supplementary Figure 32. Precision−Recall with 15 binary nodes when OR between 2−2.5 in simulation study 2

MRPC and cGAUGE are not listed due to their huge time consuming.

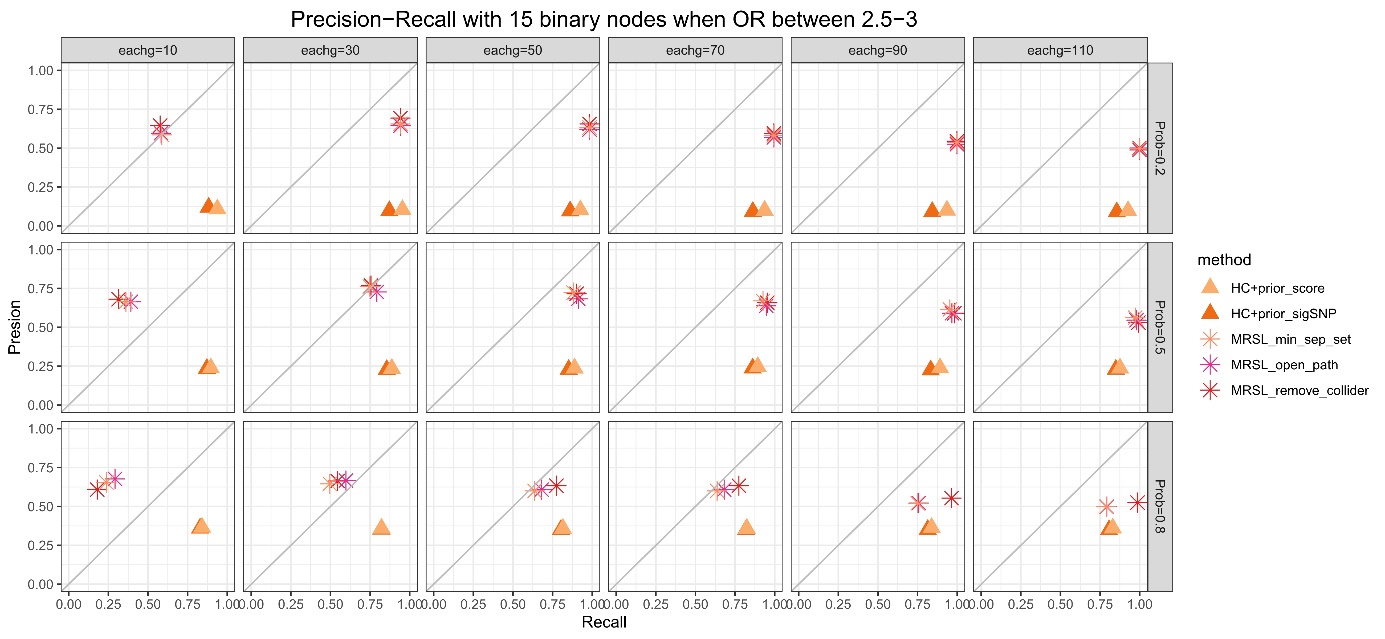

### Supplementary Figure 33. Precision−Recall with 15 binary nodes when OR between 2.5−3 in simulation study 2

MRPC and cGAUGE are not listed due to their huge time consuming.

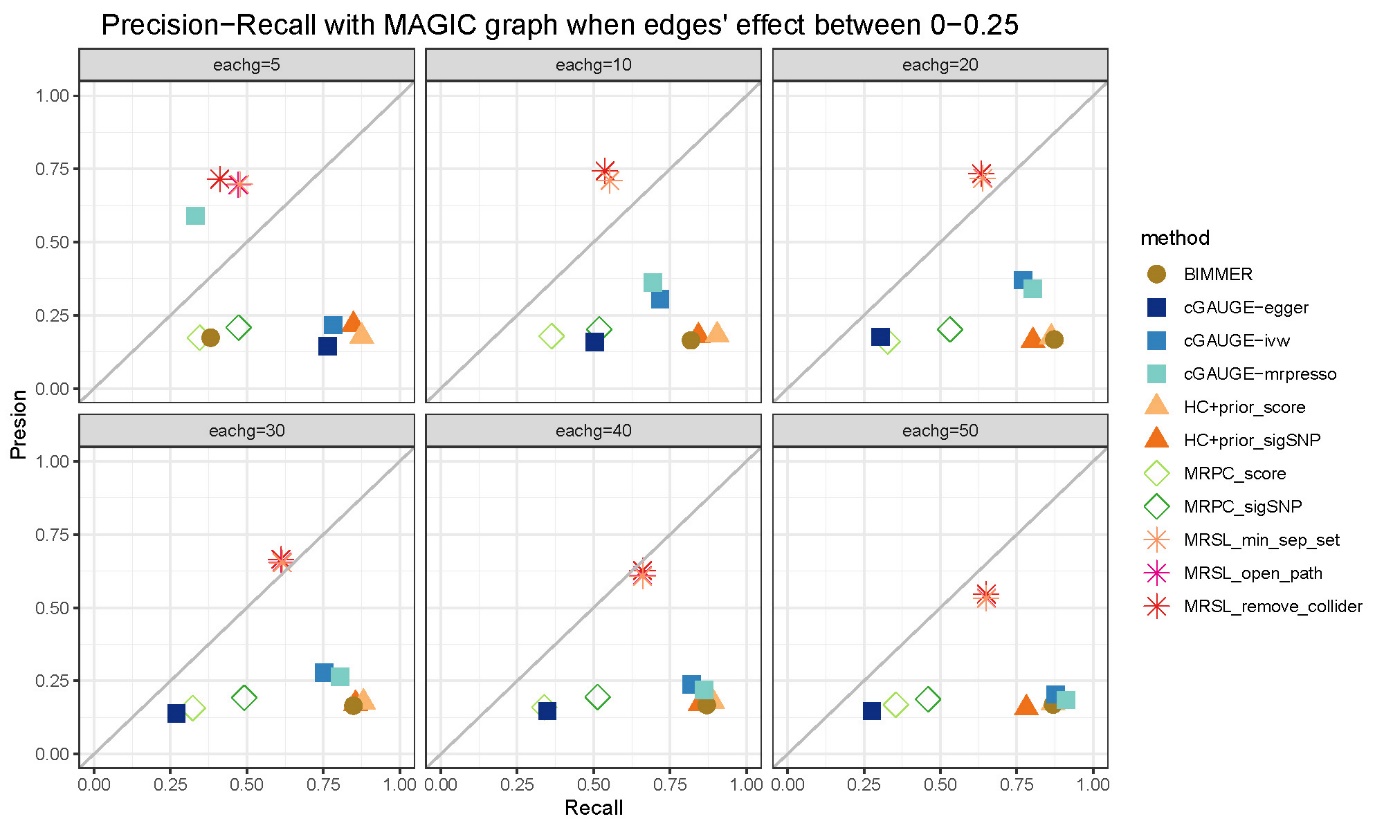

### Supplementary Figure 34. Precision−Recall with MAGIC graph when edges' effect between 0−0.25 in simulation study 3

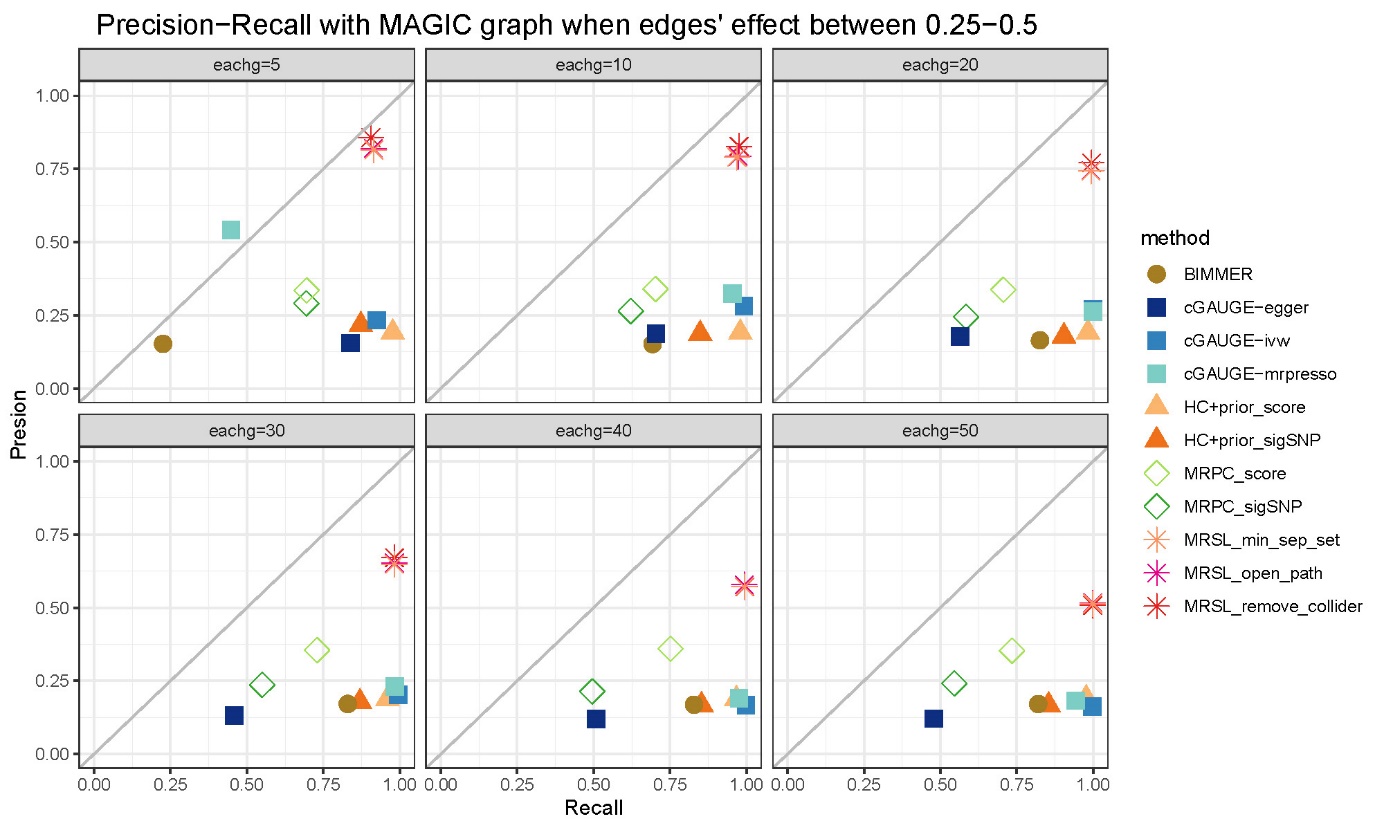

### Supplementary Figure 35. Precision−Recall with MAGIC graph when edges' effect between 0.25−0.5 in simulation study 3

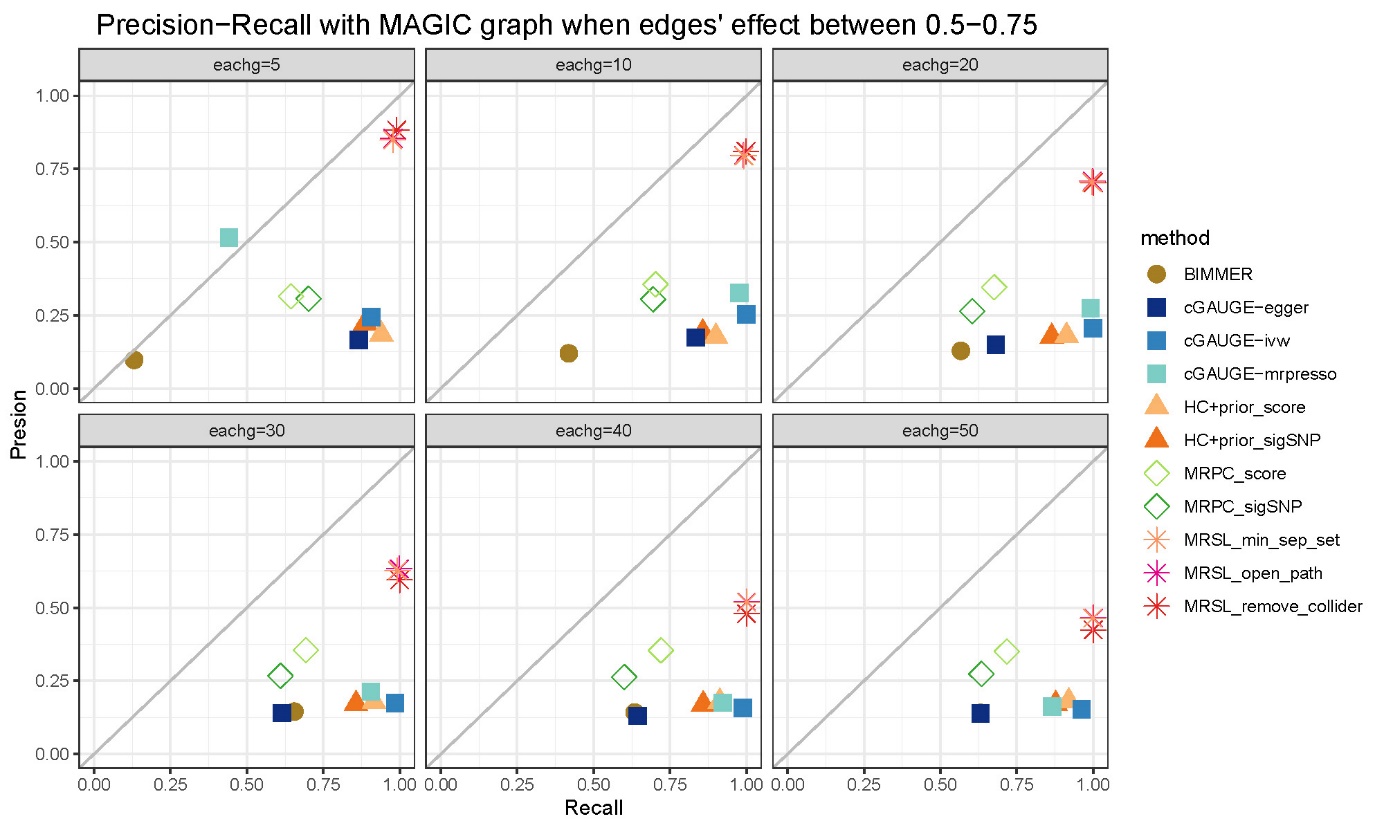

### Supplementary Figure 36. Precision−Recall with MAGIC graph when edges' effect between 0.5−0.75 in simulation study 3

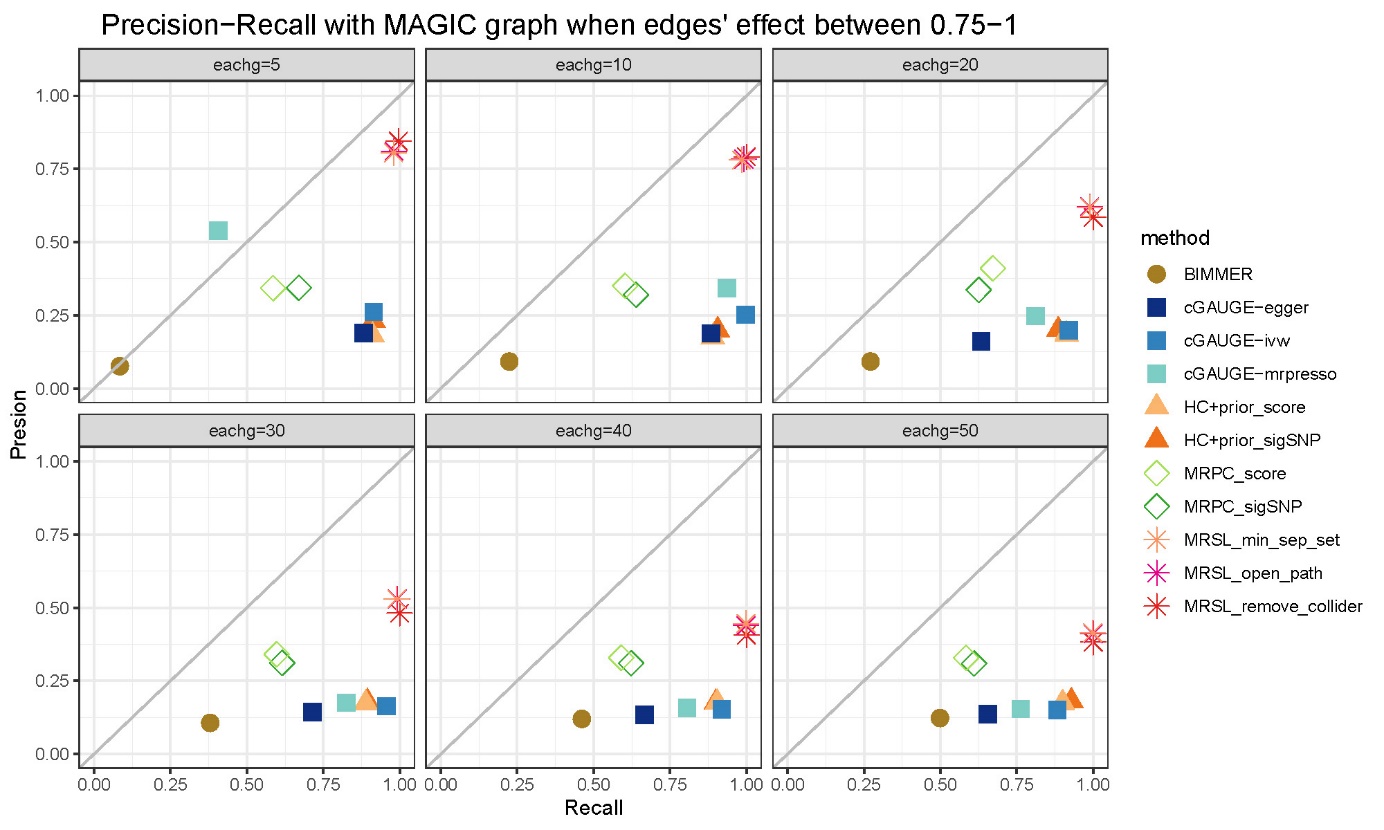

### Supplementary Figure 37. Precision−Recall with MAGIC graph when edges' effect between 0.75−1 in simulation study 3

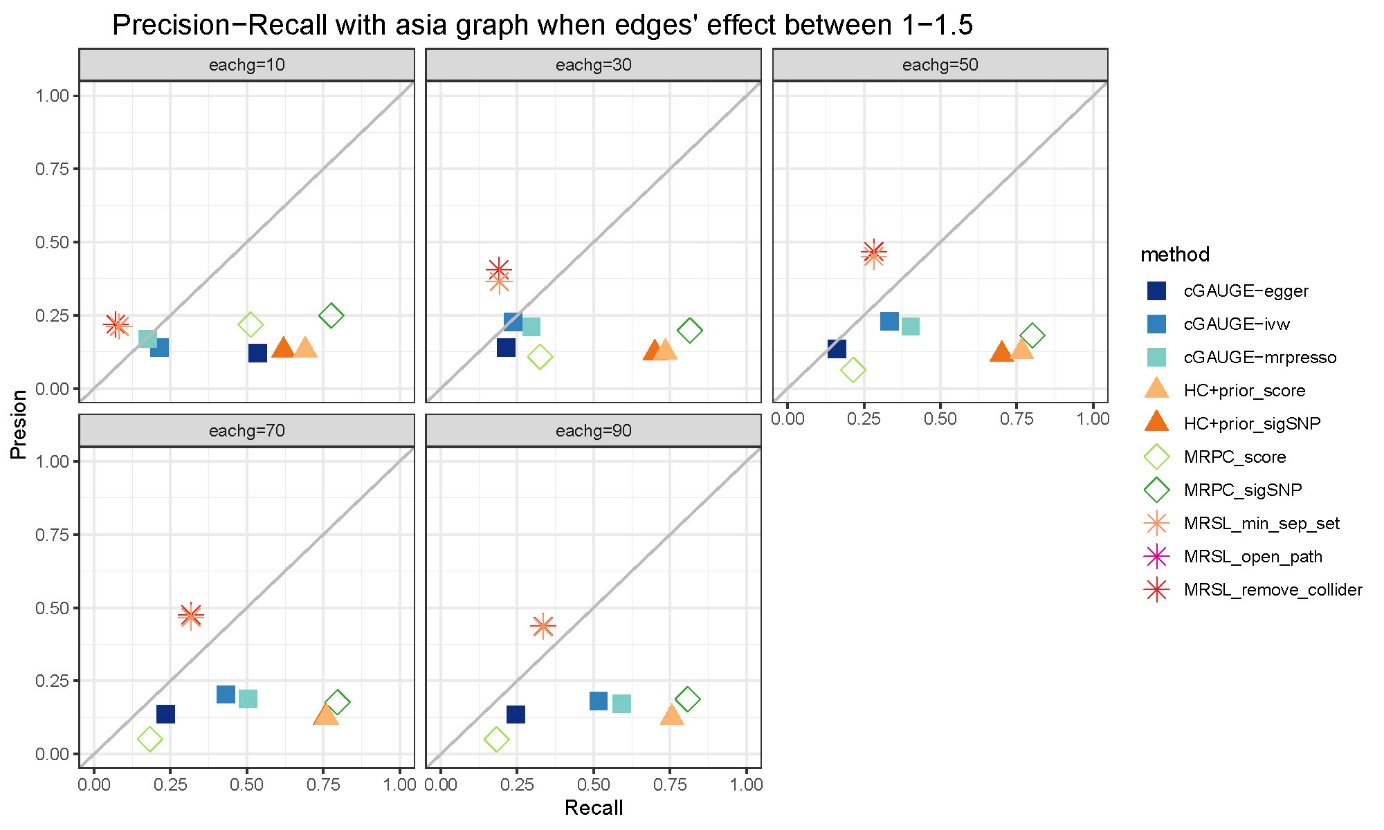

### Supplementary Figure 38. Precision−Recall with ASIA graph when OR between 1−1.5 in simulation study 3

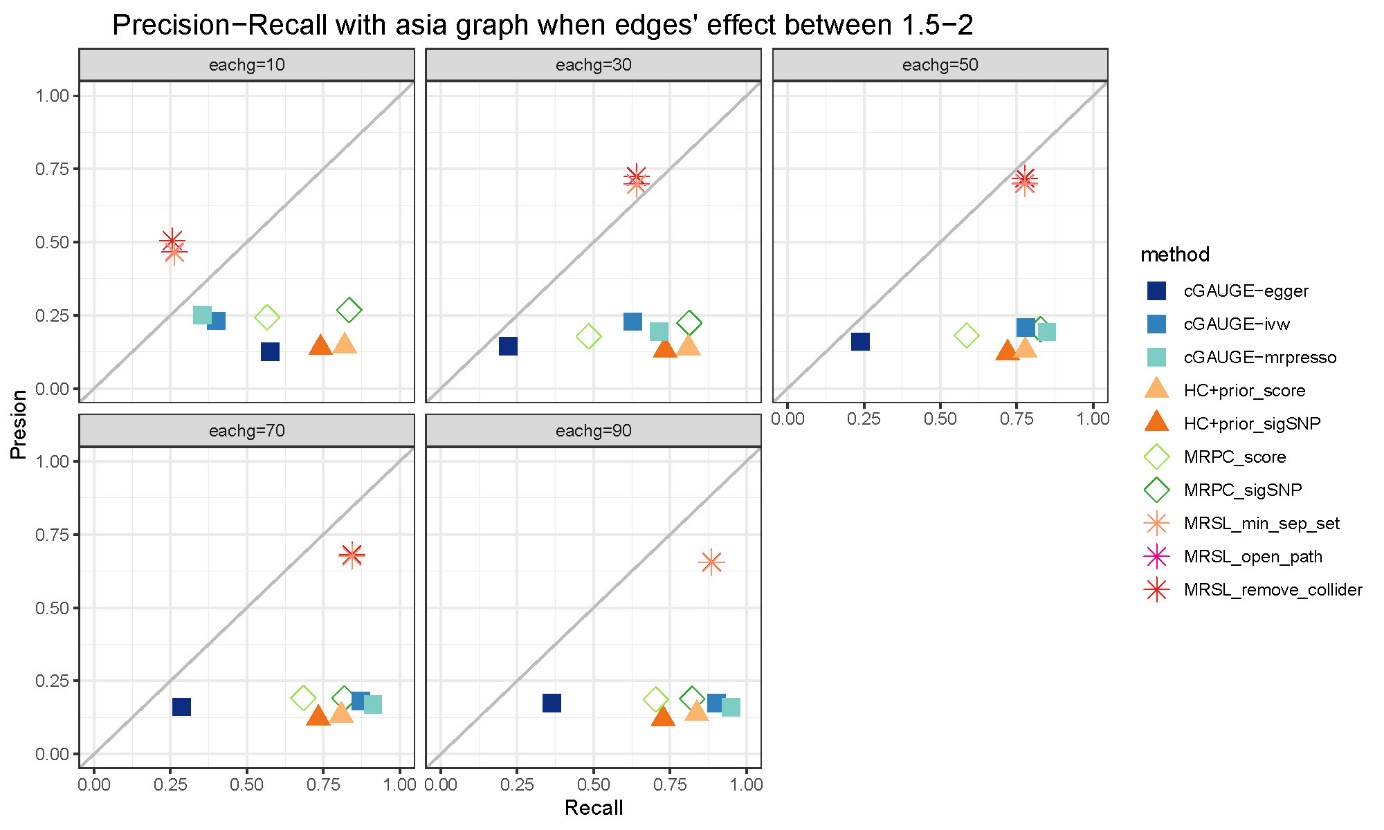

### Supplementary Figure 39. Precision−Recall with ASIA graph when OR between 1.5−2 in simulation study 3

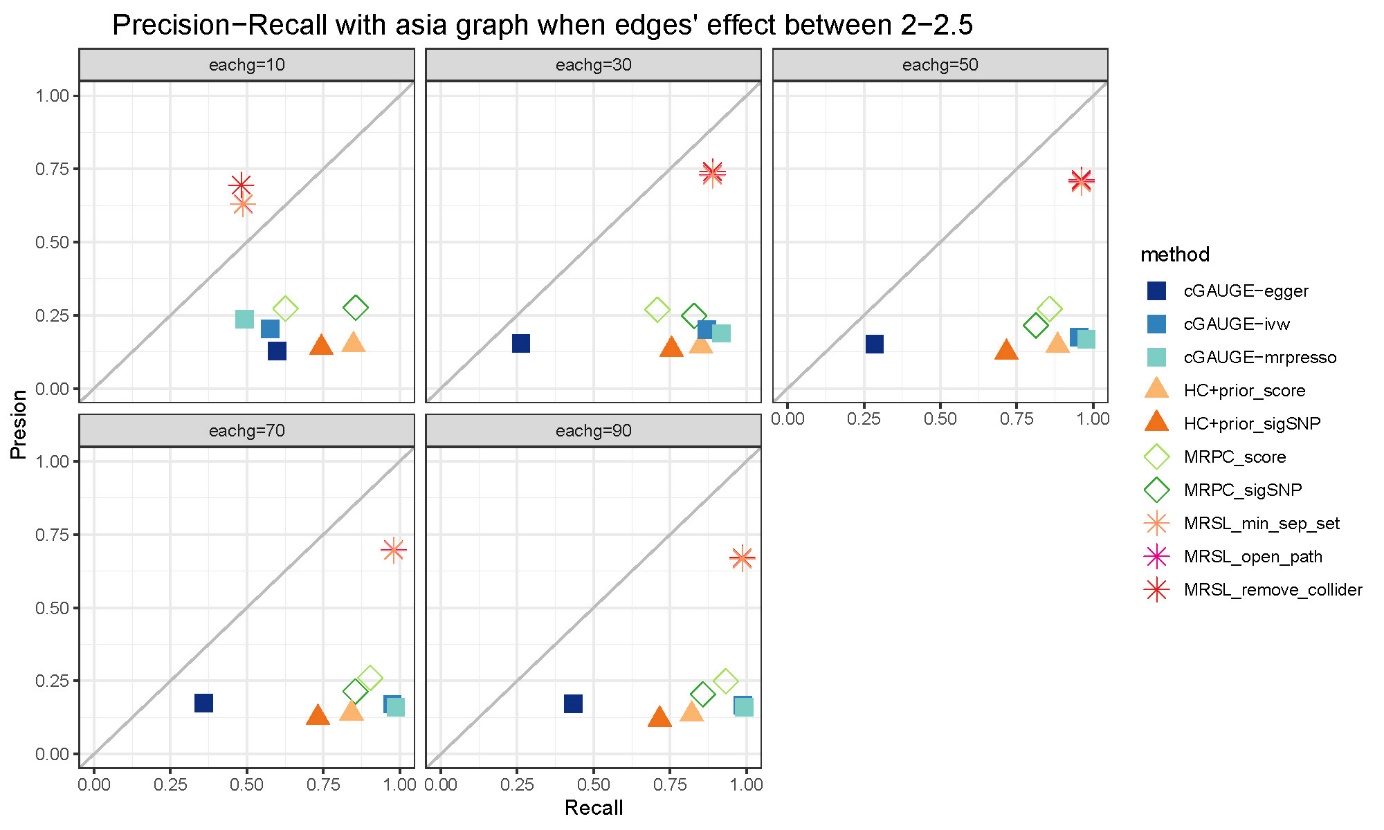

### Supplementary Figure 40. Precision−Recall with ASIA graph when OR between 2−2.5 in simulation study 3

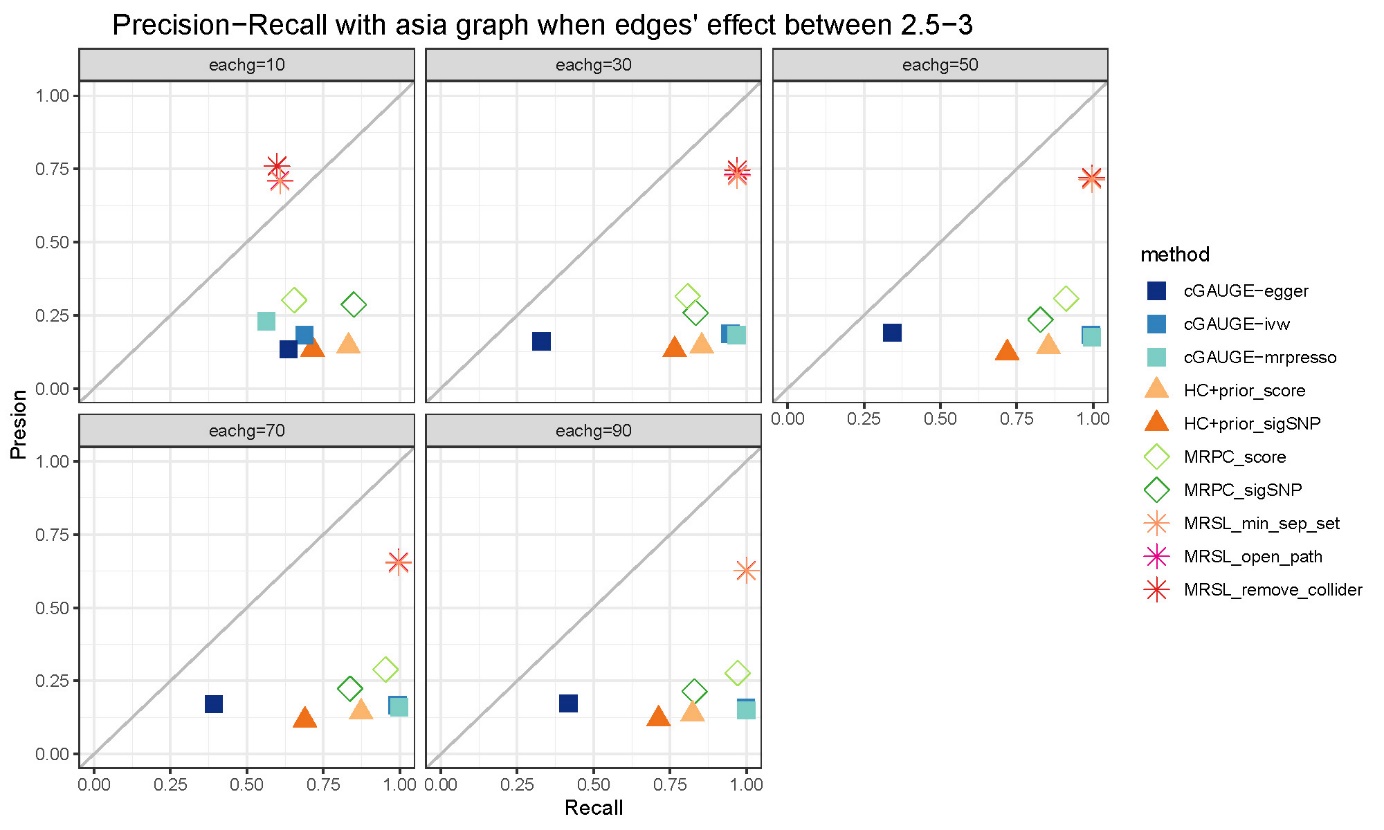

### Supplementary Figure 41. Precision−Recall with ASIA graph when OR between 2.5−3 in simulation study 3

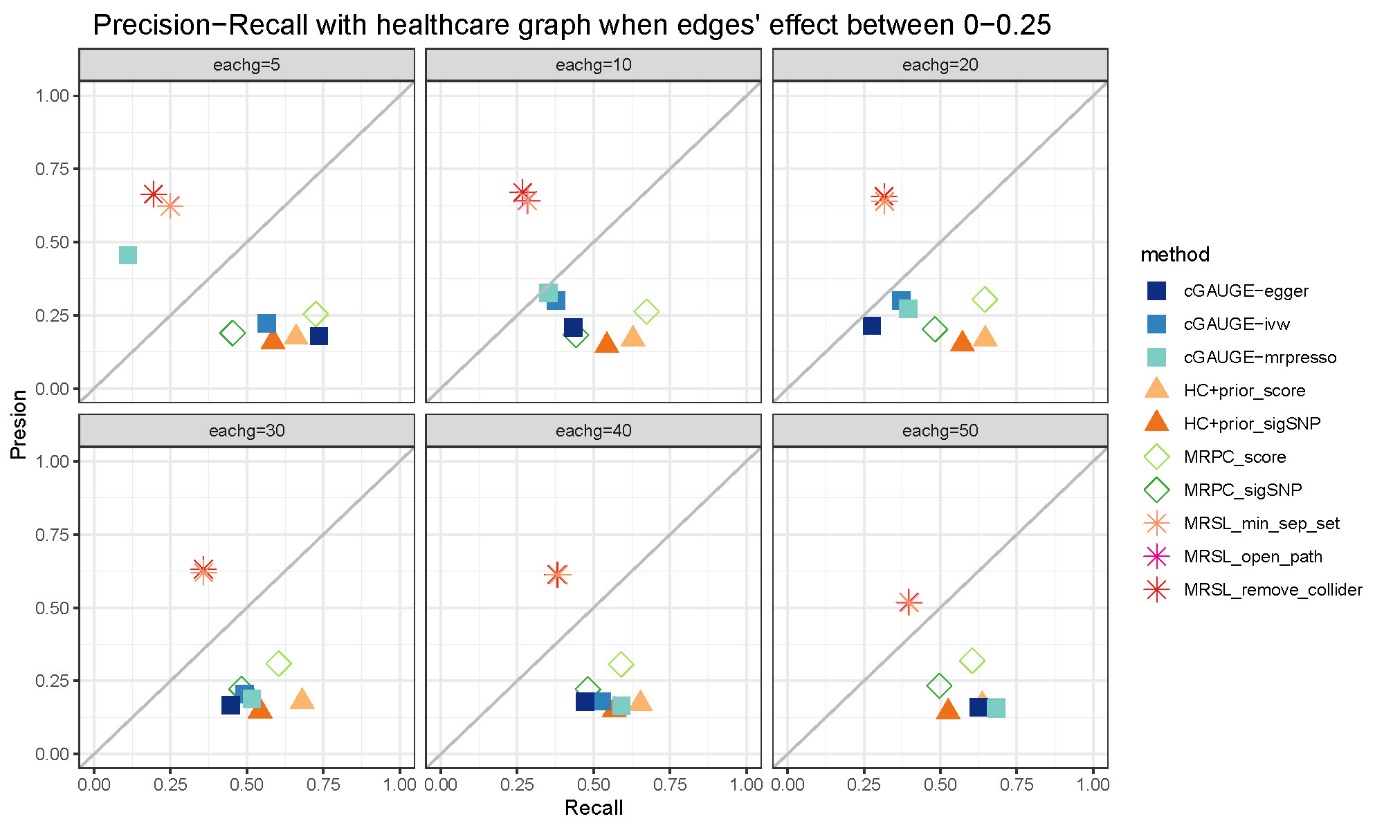

### Supplementary Figure 42. Precision−Recall with Healthcare Cost graph when edges' effect between 0−0.25

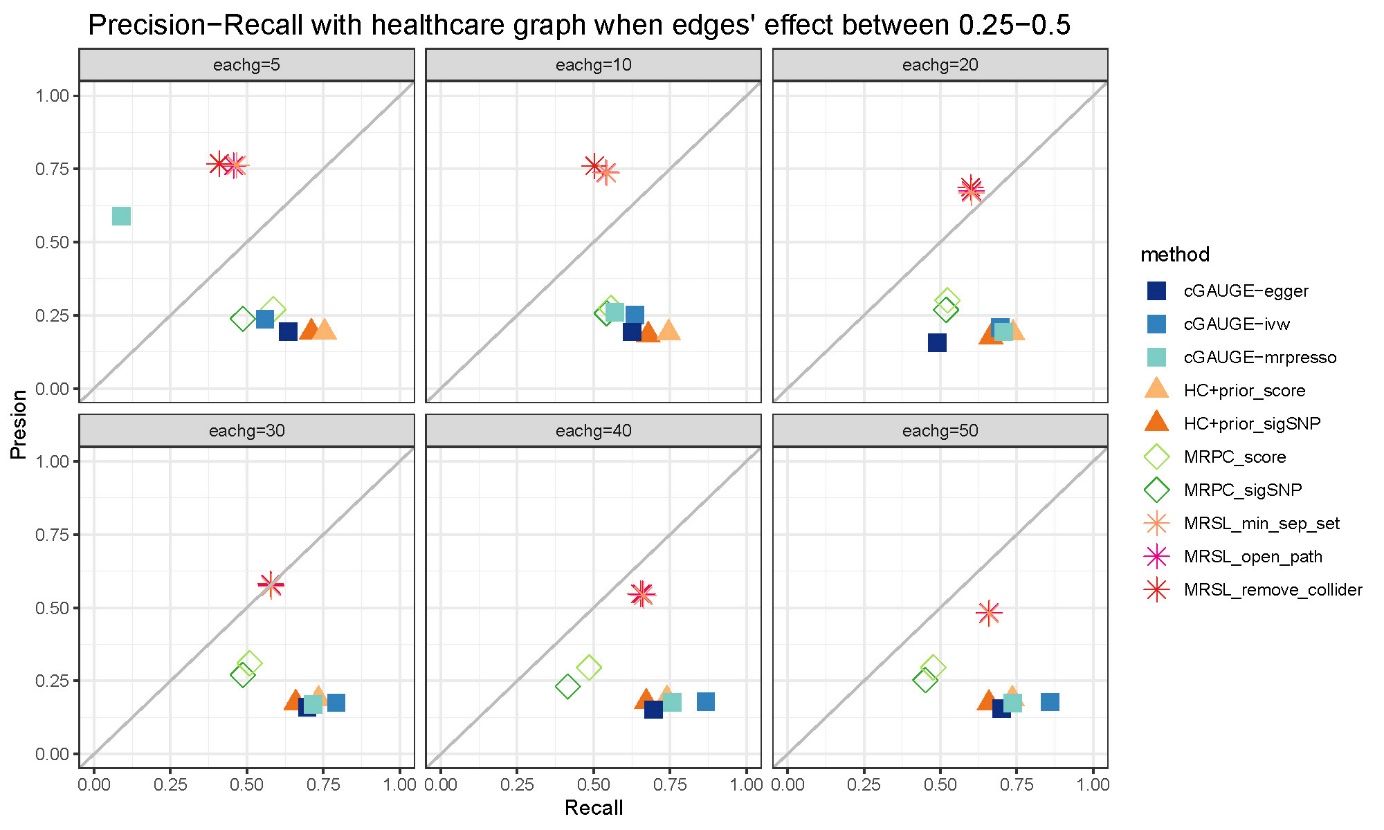

### Supplementary Figure 43. Precision−Recall with Healthcare Cost graph when edges' effect between 0.25−0.5

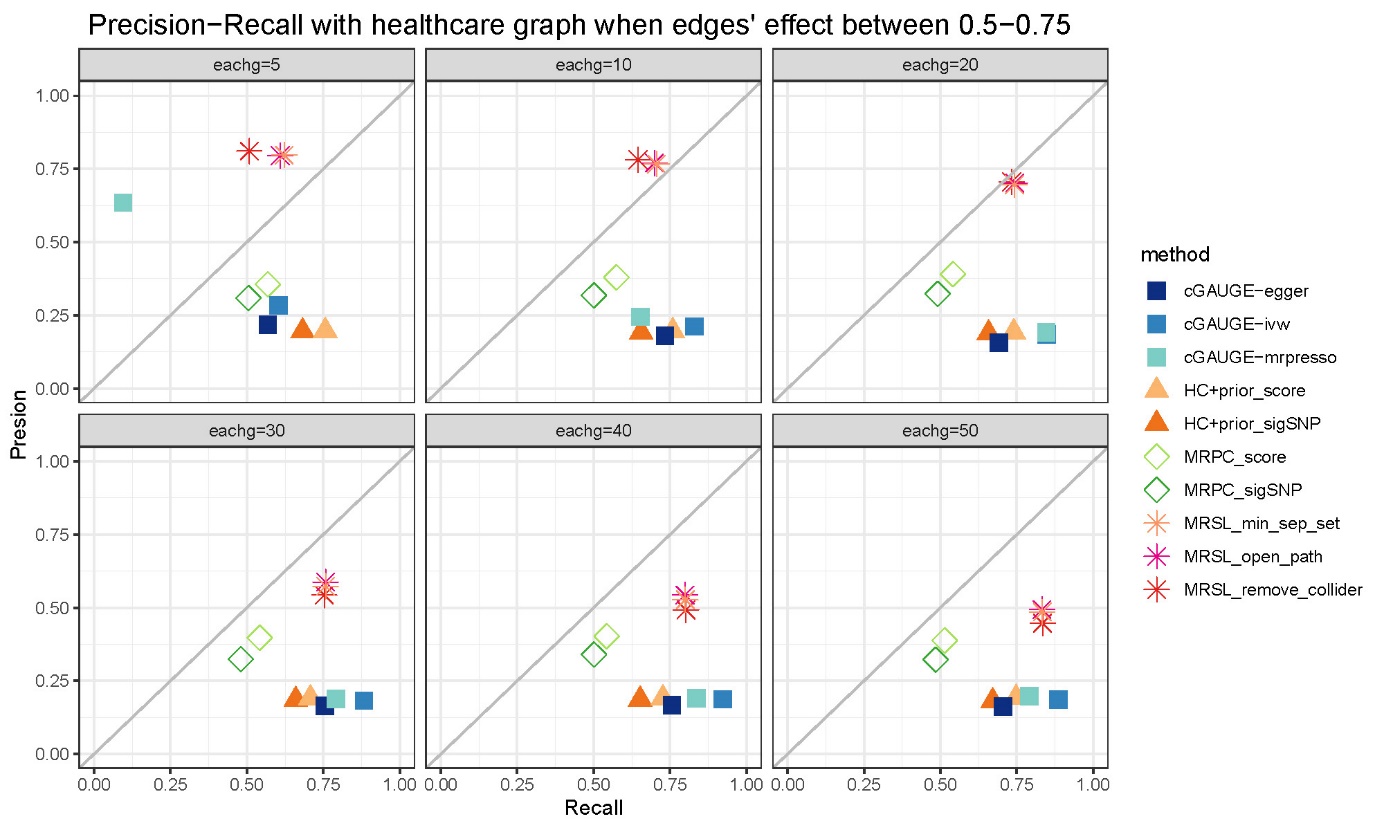

### Supplementary Figure 44. Precision−Recall with Healthcare Cost graph when edges' effect between 0.5−0.75

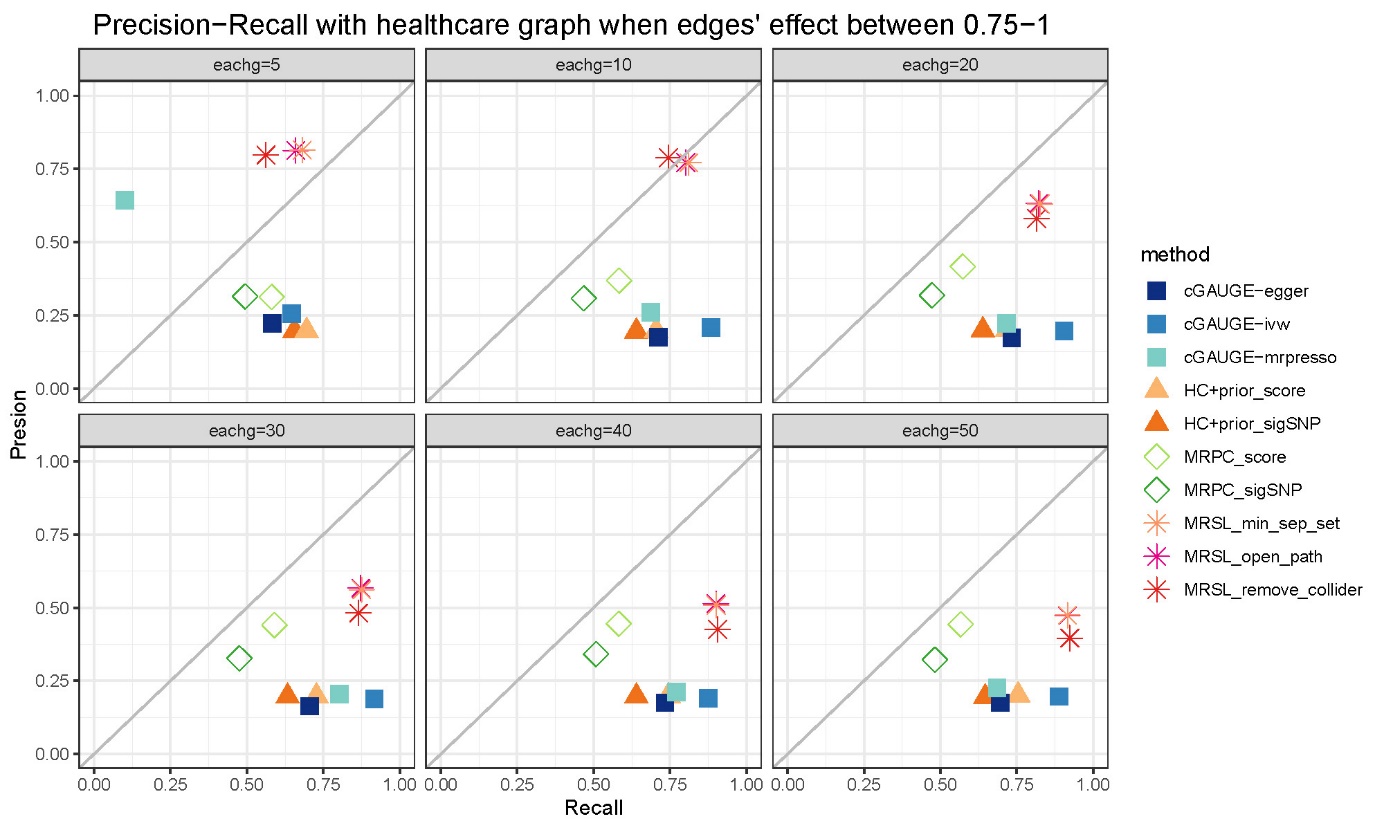

### Supplementary Figure 45. Precision−Recall with Healthcare Cost graph when edges' effect between 0.75−1

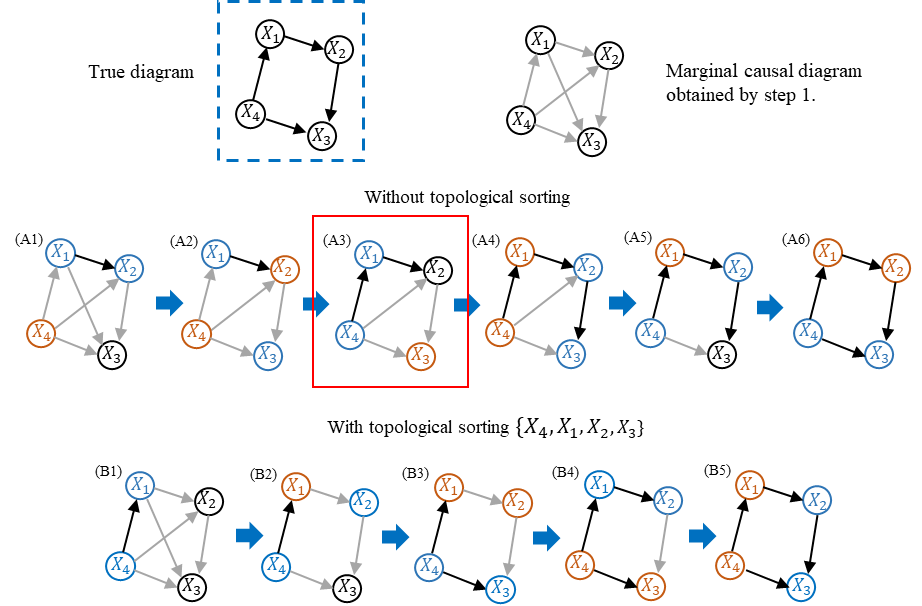

### Supplementary Figure 46. Example for MRSL with and without topological sorting

### The bias formula of causal estimation when adjusting a collider using MVMR

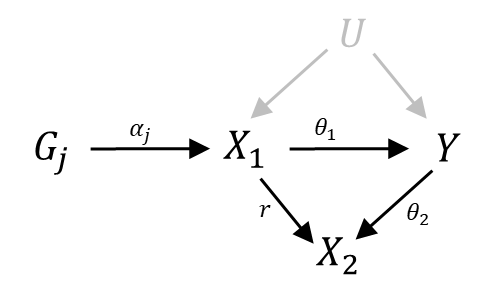

**Supplementary Figure 47.** DAG for MVMR adjusting for a collider

The unbiased estimation for causal effect of $X_{1}$ on Y can be obtained by the following weighted regression:

$\hat{\beta}_{Y_{j}}=\theta_{1}\hat{\beta}_{X_{1j}}+\varepsilon_{j},\varepsilon_{j}\sim N(0,{se(\hat{\beta}_{Y_{j}})}^{2})$.

When we perform MVMR

$\hat{\beta}_{Y_{j}}=\theta_{1}^{'}\hat{\beta}_{X_{1j}}$+$\theta_{2}^{'}\hat{\beta}_{X_{2j}}+\varepsilon_{j},\varepsilon_{j}\sim N(0,{se(\hat{\beta}_{Y_{j}})}^{2})$.

The the estimation of $\theta_{1}^{'}$ is

$\hat{\theta}_{1}^{'}=\frac{\hat{\beta}_{X_{1}}^{T}\hat{\beta}_{Y}\cdot\hat{\beta}_{X_{2}}^{T}\hat{\beta}_{X_{2}}-\hat{\beta}_{X_{2}}^{T}\hat{\beta}_{Y}\cdot\hat{\beta}_{X_{1}}^{T}\hat{\beta}_{X_{2}}}{\hat{\beta}_{X_{1}}^{T}\hat{\beta}_{X_{1}}\cdot\hat{\beta}_{X_{2}}^{T}\hat{\beta}_{X_{2}}-\hat{\beta}_{X_{1}}^{T}\hat{\beta}_{X_{2}}\cdot\hat{\beta}_{X_{2}}^{T}\hat{\beta}_{X_{1}}}$.

The bias is

$$\hat{\theta}_{1}^{'}-\hat{\theta}_{1}=\frac{\hat{\beta}_{X_{1}}^{T}\hat{\beta}_{X_{1}}\cdot\left( \hat{\beta}_{X_{1}}^{T}\hat{\beta}_{Y}\cdot\hat{\beta}_{X_{2}}^{T}\hat{\beta}_{X_{2}}-\hat{\beta}_{X_{2}}^{T}\hat{\beta}_{Y}\cdot\hat{\beta}_{X_{1}}^{T}\hat{\beta}_{X_{2}} \right)-\hat{\beta}_{X_{1}}^{T}\hat{\beta}_{Y}\cdot(\hat{\beta}_{X_{1}}^{T}\hat{\beta}_{X_{1}}\cdot\hat{\beta}_{X_{2}}^{T}\hat{\beta}_{X_{2}}-\hat{\beta}_{X_{1}}^{T}\beta_{X_{2}}\cdot\hat{\beta}_{X_{2}}^{T}\hat{\beta}_{X_{1}})}{\hat{\beta}_{X_{1}}^{T}\hat{\beta}_{X_{1}}\cdot(\hat{\beta}_{X_{1}}^{T}\hat{\beta}_{X_{1}}\cdot\hat{\beta}_{X_{2}}^{T}\hat{\beta}_{X_{2}}-\beta_{X_{1}}^{T}\hat{\beta}_{X_{2}}\cdot\beta_{X_{2}}^{T}\hat{\beta}_{X_{1}})}$$

$$=\frac{\hat{\beta}_{X_{1}}^{T}\hat{\beta}_{X_{1}}\cdot\left( -\hat{\beta}_{X_{2}}^{T}\hat{\beta}_{Y}\cdot\hat{\beta}_{X_{1}}^{T}\hat{\beta}_{X_{2}} \right)-\hat{\beta}_{X_{1}}^{T}\hat{\beta}_{Y}\cdot(-\hat{\beta}_{X_{1}}^{T}\hat{\beta}_{X_{2}}\cdot\hat{\beta}_{X_{2}}^{T}\hat{\beta}_{X_{1}})}{\hat{\beta}_{X_{1}}^{T}\hat{\beta}_{X_{1}}\cdot(\hat{\beta}_{X_{1}}^{T}\hat{\beta}_{X_{1}}\cdot\hat{\beta}_{X_{2}}^{T}\hat{\beta}_{X_{2}}-\hat{\beta}_{X_{1}}^{T}\hat{\beta}_{X_{2}}\cdot\hat{\beta}_{X_{2}}^{T}\hat{\beta}_{X_{1}})}$$

$$=\frac{\hat{\beta}_{X_{1}}^{T}\hat{\beta}_{X_{2}}\cdot(\hat{\beta}_{X_{1}}^{T}\hat{\beta}_{Y}\cdot\hat{\beta}_{X_{2}}^{T}\hat{\beta}_{X_{1}}-\hat{\beta}_{X_{1}}^{T}\hat{\beta}_{X_{1}}\cdot\hat{\beta}_{X_{2}}^{T}\hat{\beta}_{Y})}{\hat{\beta}_{X_{1}}^{T}\hat{\beta}_{X_{1}}\cdot(\hat{\beta}_{X_{1}}^{T}\hat{\beta}_{X_{1}}\cdot\hat{\beta}_{X_{2}}^{T}\hat{\beta}_{X_{2}}-\hat{\beta}_{X_{1}}^{T}\hat{\beta}_{X_{2}}\cdot\hat{\beta}_{X_{2}}^{T}\hat{\beta}_{X_{1}})}$$

### Supplementary Table 1. Computing time with network of 10 continuous nodes in simulation study 2 (seconds).

| edge effect | g | prob | MRSL^1^ | MRSL^2^ | MRSL^3^ | MRPC^1^ | MRPC^2^ | HC^1^ | HC^2^ | BIMMER | cGAUGE^1^ | cGAUGE^2^ | cGAUGE^3^ |
| --- | --- | --- | --- | --- | --- | --- | --- | --- | --- | --- | --- | --- | --- |
| 0-0.25 | 5 | 0.2 | 0.44 | 0.63 | 0.01 | 10.18 | 9.98 | 1.38 | 1.09 | 9.63 | 89.86 | 93.17 | 153.23 |
|  |  | 0.5 | 1.16 | 1.55 | 0.01 | 12.42 | 11.49 | 2.24 | 1.62 | 10.06 | 104.54 | 106.87 | 167.74 |
|  |  | 0.8 | 2.86 | 2.49 | 0.01 | 11.00 | 11.73 | 2.70 | 2.27 | 9.62 | 107.74 | 105.53 | 162.45 |
|  | 10 | 0.2 | 0.49 | 0.70 | 0.01 | 62.28 | 11.50 | 2.28 | 1.22 | 8.00 | 170.85 | 174.88 | 369.29 |
|  |  | 0.5 | 1.40 | 1.51 | 0.01 | 59.68 | 11.10 | 3.03 | 1.58 | 9.05 | 176.03 | 182.82 | 372.44 |
|  |  | 0.8 | 4.18 | 3.30 | 0.02 | 66.91 | 10.58 | 3.17 | 2.08 | 9.35 | 149.90 | 141.86 | 274.87 |
|  | 20 | 0.2 | 0.25 | 0.33 | 0.01 | 318.35 | 6.99 | 2.00 | 0.86 | 3.47 | 187.00 | 180.95 | 374.32 |
|  |  | 0.5 | 0.79 | 0.89 | 0.02 | 524.53 | 5.94 | 2.47 | 1.08 | 4.07 | 188.95 | 185.82 | 375.04 |
|  |  | 0.8 | 1.97 | 1.95 | 0.03 | 225.19 | 5.92 | 2.44 | 1.54 | 5.02 | 189.15 | 186.96 | 377.61 |
|  | 30 | 0.2 | 0.26 | 0.33 | 0.02 | 358.74 | 6.42 | 1.74 | 0.77 | 3.38 | 264.44 | 261.51 | 483.54 |
|  |  | 0.5 | 0.81 | 0.80 | 0.03 | 433.10 | 6.45 | 2.46 | 1.08 | 4.28 | 291.43 | 304.86 | 632.83 |
|  |  | 0.8 | 3.20 | 3.06 | 0.09 | 532.94 | 11.56 | 3.49 | 2.27 | 9.92 | 472.89 | 468.45 | 836.35 |
|  | 40 | 0.2 | 0.41 | 0.54 | 0.06 | 438.62 | 13.90 | 2.84 | 1.46 | 6.45 | 609.54 | 625.28 | 1107.09 |
|  |  | 0.5 | 1.65 | 1.62 | 0.10 | 1159.72 | 11.64 | 3.63 | 1.83 | 7.18 | 602.11 | 565.83 | 1049.76 |
|  |  | 0.8 | 3.53 | 4.00 | 0.16 | 548.12 | 11.62 | 3.61 | 2.52 | 11.34 | 630.67 | 618.76 | 1037.17 |
|  | 50 | 0.2 | 0.43 | 0.54 | 0.08 | 454.50 | 13.33 | 2.69 | 1.45 | 6.33 | 769.21 | 755.44 | 1177.71 |
|  |  | 0.5 | 1.62 | 1.62 | 0.15 | 620.90 | 11.57 | 3.31 | 1.86 | 7.64 | 761.88 | 716.36 | 1161.77 |
|  |  | 0.8 | 3.58 | 3.66 | 0.24 | 556.61 | 11.41 | 3.33 | 2.51 | 11.42 | 740.36 | 716.61 | 1130.72 |
| 0.25-0.5 | 5 | 0.2 | 1.17 | 1.74 | 0.01 | 7.78 | 11.09 | 2.06 | 1.33 | 12.31 | 112.22 | 108.91 | 141.70 |
|  |  | 0.5 | 6.29 | 4.16 | 0.02 | 5.34 | 10.03 | 2.65 | 2.99 | 8.63 | 115.11 | 112.67 | 139.95 |
|  |  | 0.8 | 8.35 | 6.30 | 0.03 | 3.90 | 8.87 | 2.25 | 3.50 | 6.98 | 122.92 | 111.16 | 122.10 |
|  | 10 | 0.2 | 1.02 | 1.31 | 0.01 | 23.29 | 8.86 | 2.48 | 1.19 | 10.84 | 189.62 | 184.54 | 369.08 |
|  |  | 0.5 | 5.43 | 3.77 | 0.03 | 12.91 | 8.99 | 2.59 | 2.78 | 9.34 | 194.07 | 197.01 | 345.75 |
|  |  | 0.8 | 6.93 | 6.53 | 0.04 | 3.99 | 7.54 | 1.90 | 3.09 | 6.72 | 186.45 | 191.20 | 300.31 |
|  | 20 | 0.2 | 1.10 | 1.39 | 0.03 | 110.26 | 9.65 | 2.77 | 1.21 | 8.42 | 338.80 | 332.62 | 610.84 |
|  |  | 0.5 | 6.15 | 5.02 | 0.07 | 33.04 | 8.77 | 2.73 | 3.06 | 11.34 | 362.36 | 353.72 | 571.79 |
|  |  | 0.8 | 8.54 | 10.07 | 0.08 | 5.60 | 7.53 | 2.00 | 3.34 | 7.43 | 344.11 | 339.55 | 523.73 |
|  | 30 | 0.2 | 1.33 | 1.48 | 0.06 | 98.30 | 7.28 | 2.14 | 1.05 | 8.19 | 471.69 | 477.34 | 679.46 |
|  |  | 0.5 | 6.00 | 5.28 | 0.12 | 32.94 | 8.25 | 2.48 | 2.55 | 11.39 | 508.80 | 497.39 | 639.58 |
|  |  | 0.8 | 10.46 | 13.60 | 0.10 | 4.93 | 7.28 | 1.80 | 2.97 | 7.55 | 510.18 | 483.41 | 621.97 |
|  | 40 | 0.2 | 1.24 | 1.47 | 0.10 | 65.86 | 8.39 | 2.12 | 1.23 | 8.66 | 620.99 | 628.76 | 830.62 |
|  |  | 0.5 | 6.51 | 6.63 | 0.20 | 31.50 | 7.80 | 2.26 | 2.69 | 12.14 | 648.79 | 631.50 | 786.62 |
|  |  | 0.8 | 11.71 | 16.66 | 0.11 | 4.86 | 7.49 | 1.85 | 3.15 | 7.60 | 637.01 | 635.31 | 747.73 |
|  | 50 | 0.2 | 1.31 | 1.47 | 0.14 | 56.75 | 8.11 | 1.90 | 1.17 | 8.64 | 743.79 | 757.10 | 960.77 |
|  |  | 0.5 | 7.11 | 7.50 | 0.28 | 25.80 | 8.48 | 2.41 | 2.76 | 14.58 | 791.12 | 733.85 | 942.95 |
|  |  | 0.8 | 15.77 | 18.18 | 0.11 | 4.00 | 7.47 | 1.77 | 3.15 | 7.90 | 773.35 | 755.31 | 895.70 |
| 0.5-0.75 | 5 | 0.2 | 1.28 | 1.81 | 0.01 | 4.77 | 10.17 | 2.06 | 1.53 | 10.40 | 110.09 | 105.68 | 132.13 |
|  |  | 0.5 | 6.99 | 4.42 | 0.02 | 2.86 | 5.44 | 2.43 | 3.53 | 6.77 | 116.25 | 108.11 | 115.02 |
|  |  | 0.8 | 9.75 | 8.56 | 0.03 | 1.91 | 3.25 | 1.85 | 3.15 | 5.42 | 114.58 | 108.85 | 117.62 |
|  | 10 | 0.2 | 1.43 | 1.95 | 0.02 | 12.40 | 8.78 | 2.38 | 1.40 | 11.30 | 186.49 | 183.82 | 332.79 |
|  |  | 0.5 | 6.44 | 4.89 | 0.03 | 2.59 | 4.92 | 2.08 | 3.09 | 6.58 | 180.79 | 167.62 | 251.79 |
|  |  | 0.8 | 11.43 | 13.31 | 0.03 | 1.92 | 3.15 | 1.72 | 2.86 | 5.89 | 197.75 | 191.00 | 238.67 |
|  | 20 | 0.2 | 1.52 | 1.83 | 0.04 | 24.12 | 7.52 | 2.41 | 1.50 | 9.56 | 344.63 | 323.82 | 551.96 |
|  |  | 0.5 | 8.27 | 8.21 | 0.06 | 4.20 | 4.94 | 2.04 | 3.21 | 7.29 | 351.35 | 350.21 | 442.85 |
|  |  | 0.8 | 12.58 | 17.88 | 0.03 | 1.52 | 2.70 | 1.51 | 2.73 | 5.55 | 354.66 | 347.40 | 377.69 |
|  | 30 | 0.2 | 1.73 | 1.95 | 0.07 | 39.08 | 7.15 | 2.08 | 1.36 | 9.95 | 495.09 | 482.30 | 648.55 |
|  |  | 0.5 | 4.95 | 5.99 | 0.05 | 3.29 | 2.31 | 1.15 | 1.80 | 3.76 | 271.43 | 269.86 | 323.41 |
|  |  | 0.8 | 6.93 | 10.06 | 0.02 | 0.75 | 1.36 | 0.84 | 1.58 | 2.90 | 269.35 | 267.84 | 281.50 |
|  | 40 | 0.2 | 0.98 | 1.12 | 0.05 | 18.69 | 4.16 | 1.23 | 0.86 | 5.12 | 341.58 | 340.39 | 443.68 |
|  |  | 0.5 | 5.97 | 7.34 | 0.06 | 1.47 | 2.37 | 1.14 | 1.86 | 3.95 | 346.86 | 345.11 | 399.00 |
|  |  | 0.8 | 7.58 | 10.74 | 0.02 | 0.68 | 1.27 | 0.82 | 1.61 | 2.93 | 344.79 | 343.40 | 358.77 |
|  | 50 | 0.2 | 1.20 | 1.23 | 0.08 | 15.27 | 3.37 | 1.05 | 0.78 | 5.15 | 417.83 | 416.32 | 517.33 |
|  |  | 0.5 | 8.42 | 12.04 | 0.10 | 2.00 | 3.54 | 1.53 | 2.63 | 5.72 | 589.59 | 651.33 | 864.82 |
|  |  | 0.8 | 15.49 | 23.46 | 0.03 | 1.43 | 3.10 | 1.41 | 2.77 | 5.86 | 779.40 | 762.43 | 789.32 |
| 0.75-1 | 5 | 0.2 | 1.38 | 1.68 | 0.01 | 2.50 | 5.55 | 1.86 | 1.55 | 8.89 | 105.81 | 99.97 | 113.52 |
|  |  | 0.5 | 8.34 | 6.12 | 0.02 | 1.71 | 3.30 | 2.30 | 3.33 | 6.15 | 119.01 | 112.45 | 113.43 |
|  |  | 0.8 | 10.01 | 10.06 | 0.02 | 1.22 | 1.30 | 1.41 | 2.20 | 4.24 | 101.53 | 92.05 | 103.46 |
|  | 10 | 0.2 | 1.98 | 2.10 | 0.02 | 9.48 | 6.78 | 2.39 | 1.78 | 10.15 | 187.27 | 182.40 | 285.24 |
|  |  | 0.5 | 10.06 | 9.33 | 0.03 | 1.62 | 3.12 | 1.90 | 3.08 | 6.18 | 188.20 | 180.01 | 228.00 |
|  |  | 0.8 | 13.86 | 17.23 | 0.02 | 1.34 | 1.65 | 1.53 | 2.65 | 5.67 | 194.16 | 189.48 | 214.66 |
|  | 20 | 0.2 | 1.86 | 2.07 | 0.04 | 11.28 | 4.41 | 2.05 | 1.49 | 8.57 | 334.46 | 335.41 | 429.89 |
|  |  | 0.5 | 11.23 | 13.83 | 0.05 | 1.61 | 2.68 | 1.87 | 2.98 | 6.15 | 359.54 | 347.45 | 374.20 |
|  |  | 0.8 | 14.08 | 20.04 | 0.02 | 1.24 | 1.38 | 1.26 | 2.25 | 4.97 | 345.71 | 338.63 | 342.01 |
|  | 30 | 0.2 | 2.70 | 2.70 | 0.08 | 14.25 | 6.77 | 2.12 | 1.76 | 9.16 | 497.69 | 455.60 | 628.40 |
|  |  | 0.5 | 11.16 | 14.84 | 0.06 | 1.52 | 2.82 | 1.74 | 3.04 | 6.61 | 492.69 | 494.02 | 542.91 |
|  |  | 0.8 | 15.16 | 22.26 | 0.02 | 1.11 | 1.50 | 1.35 | 2.47 | 5.07 | 491.96 | 486.01 | 477.01 |
|  | 40 | 0.2 | 2.93 | 3.06 | 0.11 | 12.11 | 6.45 | 1.95 | 1.66 | 8.79 | 628.35 | 613.06 | 774.53 |
|  |  | 0.5 | 12.29 | 16.44 | 0.07 | 1.34 | 2.67 | 1.69 | 3.06 | 5.85 | 633.40 | 639.30 | 693.39 |
|  |  | 0.8 | 14.00 | 20.57 | 0.02 | 1.01 | 1.36 | 1.22 | 2.28 | 4.68 | 613.71 | 637.82 | 641.41 |
|  | 50 | 0.2 | 3.44 | 3.46 | 0.16 | 40.28 | 7.01 | 1.82 | 1.64 | 9.37 | 758.43 | 758.60 | 928.54 |
|  |  | 0.5 | 15.67 | 19.07 | 0.09 | 1.38 | 3.44 | 1.74 | 3.26 | 6.63 | 780.50 | 776.38 | 821.59 |
|  |  | 0.8 | 13.79 | 20.74 | 0.02 | 0.99 | 1.55 | 1.13 | 2.19 | 4.56 | 629.16 | 589.81 | 590.85 |

MRSL^1^, MRSL when adjusting for all nodes on the open paths; MRSL^2^, MRSL when adjusting for minimum separated set; MRSL^3^, MRSL when adjusting for V\{
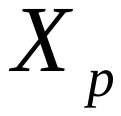
,
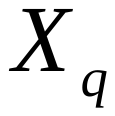
,
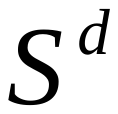
 and U}. MRPC^1^, MRPC algorithm based on the most significant SNP; MRPC^2^, MRPC algorithm based on genetic risk score. HC^1^, HC algorithm incorporating genetic anchors based on the most significant SNP; HC^2^, HC algorithm incorporating genetic anchors based on genetic risk score. cGAUGE^1^, cGAUGE based on IVW; cGAUGE^2^, cGAUGE based on MR Egger; cGAUGE^3^, cGAUGE based on MR PRESSO.

### Supplementary Table 2. Computing time with network of 10 binary nodes in simulation study 2 (seconds).

| edge effect | g | prob | MRSL^1^ | MRSL^2^ | MRSL^3^ | MRPC^1^ | MRPC^2^ | HC^1^ | HC^2^ | cGAUGE^1^ | cGAUGE^2^ | cGAUGE^3^ |
| --- | --- | --- | --- | --- | --- | --- | --- | --- | --- | --- | --- | --- |
| OR 1-1.5 | 10 | 0.2 | 0.11 | 0.13 | 0.00 | 11.75 | 1.75 | 1.38 | 0.67 | 92.53 | 91.45 | 195.02 |
|  |  | 0.5 | 0.13 | 0.16 | 0.00 | 21.69 | 4.26 | 2.15 | 1.24 | 99.18 | 98.52 | 198.48 |
|  |  | 0.8 | 0.16 | 0.21 | 0.00 | 26.13 | 5.71 | 2.27 | 1.50 | 100.97 | 100.50 | 195.60 |
|  | 30 | 0.2 | 0.13 | 0.16 | 0.01 | 195.71 | 6.67 | 2.14 | 1.26 | 251.33 | 255.37 | 591.87 |
|  |  | 0.5 | 0.32 | 0.42 | 0.03 | 164.27 | 11.71 | 3.68 | 2.40 | 452.22 | 451.41 | 1010.35 |
|  |  | 0.8 | 0.45 | 0.58 | 0.03 | 215.97 | 13.17 | 4.21 | 2.94 | 464.24 | 449.07 | 992.08 |
|  | 50 | 0.2 | 0.26 | 0.35 | 0.06 | 566.49 | 14.80 | 4.11 | 2.64 | 726.53 | 720.08 | 1559.85 |
|  |  | 0.5 | 0.39 | 0.53 | 0.07 | 247.24 | 14.79 | 3.94 | 3.12 | 754.02 | 739.02 | 1581.99 |
|  |  | 0.8 | 0.61 | 0.75 | 0.08 | 237.86 | 14.84 | 4.24 | 3.49 | 748.31 | 739.65 | 1580.56 |
|  | 70 | 0.2 | 0.28 | 0.37 | 0.10 | 257.14 | 15.67 | 4.22 | 3.37 | 1014.26 | 1017.65 | 2147.42 |
|  |  | 0.5 | 0.53 | 0.68 | 0.14 | 169.79 | 16.60 | 4.12 | 3.79 | 1014.85 | 1024.53 | 2167.73 |
|  |  | 0.8 | 0.97 | 1.05 | 0.17 | 214.79 | 17.46 | 4.25 | 4.12 | 1025.37 | 1019.90 | 2118.80 |
|  | 90 | 0.2 | 0.31 | 0.42 | 0.16 | 203.81 | 15.19 | 4.22 | 3.67 | 1302.12 | 1302.18 | 2728.89 |
|  |  | 0.5 | 0.69 | 0.80 | 0.25 | 133.96 | 18.72 | 3.95 | 4.22 | 1284.73 | 1300.76 | 2725.47 |
|  |  | 0.8 | 1.30 | 1.30 | 0.32 | 160.22 | 20.46 | 3.93 | 4.31 | 1314.60 | 1324.01 | 2693.18 |
|  | 110 | 0.2 | 0.45 | 0.57 | 0.29 | 120.10 | 17.84 | 4.21 | 4.42 | 1603.72 | 1601.96 | 3201.64 |
|  |  | 0.5 | 0.85 | 0.97 | 0.41 | 110.98 | 20.52 | 3.99 | 4.72 | 1606.83 | 1609.27 | 3170.30 |
|  |  | 0.8 | 1.46 | 1.46 | 0.47 | 107.43 | 21.83 | 3.78 | 4.80 | 1577.56 | 1578.63 | 3136.53 |
|  | 130 | 0.2 | 0.28 | 0.36 | 0.25 | 65.00 | 9.49 | 2.70 | 3.00 | 1002.92 | 993.41 | 2021.41 |
|  |  | 0.5 | 0.61 | 0.63 | 0.36 | 59.38 | 11.36 | 2.50 | 3.15 | 1006.87 | 993.74 | 1953.26 |
|  |  | 0.8 | 1.04 | 1.02 | 0.43 | 47.23 | 13.11 | 2.40 | 3.30 | 1001.13 | 997.17 | 1896.63 |
|  | 150 | 0.2 | 0.39 | 0.43 | 0.41 | 52.86 | 10.88 | 2.61 | 3.20 | 1136.99 | 1135.89 | 2205.91 |
|  |  | 0.5 | 0.85 | 0.82 | 0.55 | 41.99 | 13.00 | 2.46 | 3.30 | 1135.58 | 1134.79 | 2157.91 |
|  |  | 0.8 | 1.40 | 1.41 | 0.68 | 47.00 | 13.19 | 2.38 | 3.44 | 1145.18 | 1142.79 | 2145.34 |
|  | 170 | 0.2 | 0.46 | 0.51 | 0.55 | 51.45 | 11.04 | 2.63 | 3.42 | 1329.08 | 1310.50 | 2444.80 |
|  |  | 0.5 | 1.08 | 1.10 | 0.76 | 43.82 | 13.88 | 2.53 | 3.51 | 1294.91 | 1296.44 | 2440.78 |
|  |  | 0.8 | 1.66 | 1.78 | 0.93 | 42.37 | 15.16 | 2.44 | 3.58 | 1286.82 | 1284.81 | 2341.66 |
|  | 190 | 0.2 | 0.69 | 0.67 | 0.80 | 37.59 | 11.27 | 2.50 | 3.50 | 1441.47 | 1437.31 | 2639.68 |
|  |  | 0.5 | 1.48 | 1.39 | 1.11 | 38.40 | 14.66 | 2.42 | 3.56 | 1440.29 | 1438.36 | 2598.63 |
|  |  | 0.8 | 2.03 | 2.37 | 1.26 | 32.10 | 15.84 | 2.37 | 3.67 | 1441.53 | 1438.41 | 2564.43 |
| OR 1.5-2 | 10 | 0.2 | 0.28 | 0.37 | 0.01 | 28.37 | 3.51 | 2.81 | 1.31 | 179.04 | 173.00 | 359.48 |
|  |  | 0.5 | 0.48 | 0.67 | 0.01 | 36.33 | 9.28 | 3.67 | 2.41 | 192.86 | 185.66 | 352.42 |
|  |  | 0.8 | 0.84 | 1.12 | 0.01 | 30.08 | 13.79 | 3.73 | 2.77 | 195.74 | 192.83 | 309.22 |
|  | 30 | 0.2 | 0.40 | 0.53 | 0.04 | 375.42 | 10.27 | 3.33 | 1.95 | 449.86 | 440.29 | 971.85 |
|  |  | 0.5 | 0.76 | 0.73 | 0.03 | 72.41 | 5.33 | 2.43 | 1.71 | 249.62 | 247.73 | 507.25 |
|  |  | 0.8 | 1.41 | 1.44 | 0.04 | 82.26 | 6.46 | 2.68 | 2.00 | 251.26 | 249.20 | 484.94 |
|  | 50 | 0.2 | 0.26 | 0.35 | 0.04 | 234.44 | 8.13 | 2.46 | 1.73 | 416.40 | 461.57 | 1180.79 |
|  |  | 0.5 | 1.67 | 1.59 | 0.13 | 175.00 | 15.80 | 3.94 | 3.34 | 759.47 | 723.71 | 1518.48 |
|  |  | 0.8 | 3.26 | 3.85 | 0.19 | 116.43 | 16.47 | 4.16 | 3.87 | 760.80 | 755.92 | 1454.25 |
|  | 70 | 0.2 | 0.61 | 0.73 | 0.15 | 322.64 | 19.17 | 3.99 | 3.27 | 972.43 | 1023.67 | 2180.56 |
|  |  | 0.5 | 2.07 | 2.11 | 0.26 | 116.92 | 21.75 | 3.65 | 3.62 | 991.65 | 1029.85 | 2056.59 |
|  |  | 0.8 | 3.56 | 4.59 | 0.30 | 112.96 | 21.39 | 3.91 | 4.21 | 1039.29 | 1032.17 | 1956.21 |
|  | 90 | 0.2 | 0.64 | 0.79 | 0.26 | 170.68 | 24.80 | 4.05 | 3.84 | 1289.43 | 1293.46 | 2711.82 |
|  |  | 0.5 | 2.24 | 2.24 | 0.38 | 116.32 | 30.10 | 3.66 | 4.15 | 1312.18 | 1314.58 | 2541.78 |
|  |  | 0.8 | 4.16 | 5.88 | 0.51 | 92.49 | 28.10 | 3.87 | 4.63 | 1307.34 | 1260.55 | 2441.44 |
|  | 110 | 0.2 | 0.81 | 0.92 | 0.40 | 175.06 | 30.74 | 4.08 | 4.40 | 1545.03 | 1536.41 | 3142.21 |
|  |  | 0.5 | 2.65 | 2.82 | 0.62 | 104.19 | 39.69 | 3.65 | 4.64 | 1593.42 | 1588.40 | 2970.55 |
|  |  | 0.8 | 5.12 | 7.01 | 0.67 | 90.90 | 36.35 | 3.79 | 4.95 | 1597.96 | 1589.77 | 2841.65 |
| OR 2-2.5 | 10 | 0.2 | 0.41 | 0.58 | 0.01 | 23.12 | 2.99 | 2.59 | 1.36 | 178.86 | 175.86 | 373.88 |
|  |  | 0.5 | 0.82 | 1.04 | 0.01 | 33.87 | 8.47 | 3.47 | 2.34 | 192.33 | 186.39 | 344.08 |
|  |  | 0.8 | 1.84 | 1.95 | 0.02 | 20.32 | 12.58 | 3.49 | 2.74 | 198.75 | 191.58 | 299.82 |
|  | 30 | 0.2 | 0.61 | 0.77 | 0.04 | 203.49 | 9.21 | 3.34 | 1.98 | 465.05 | 459.00 | 1005.46 |
|  |  | 0.5 | 2.03 | 1.87 | 0.07 | 69.77 | 9.27 | 3.45 | 2.61 | 444.90 | 454.24 | 937.39 |
|  |  | 0.8 | 5.00 | 5.50 | 0.12 | 60.42 | 12.82 | 4.06 | 3.41 | 480.09 | 477.25 | 859.34 |
|  | 50 | 0.2 | 0.69 | 0.82 | 0.10 | 345.23 | 17.21 | 3.78 | 2.71 | 740.55 | 704.71 | 1479.36 |
|  |  | 0.5 | 3.02 | 3.09 | 0.19 | 76.19 | 15.72 | 3.61 | 3.32 | 757.61 | 757.48 | 1519.41 |
|  |  | 0.8 | 5.49 | 7.20 | 0.25 | 64.63 | 15.36 | 3.81 | 3.85 | 756.66 | 733.77 | 1277.66 |
|  | 70 | 0.2 | 0.60 | 0.68 | 0.14 | 190.89 | 17.79 | 2.97 | 2.61 | 786.02 | 777.08 | 1280.74 |
|  |  | 0.5 | 1.56 | 1.70 | 0.15 | 60.05 | 12.14 | 2.23 | 2.31 | 548.96 | 550.71 | 1050.91 |
|  |  | 0.8 | 3.51 | 4.94 | 0.19 | 28.78 | 10.14 | 2.25 | 2.66 | 548.03 | 551.92 | 965.63 |
|  | 90 | 0.2 | 0.50 | 0.55 | 0.15 | 127.35 | 16.18 | 2.58 | 2.46 | 694.19 | 697.75 | 1409.39 |
|  |  | 0.5 | 1.82 | 2.28 | 0.27 | 41.66 | 16.79 | 2.17 | 2.60 | 691.43 | 692.45 | 1307.32 |
|  |  | 0.8 | 4.16 | 6.25 | 0.29 | 34.26 | 14.28 | 2.27 | 2.90 | 706.69 | 695.48 | 1185.10 |
|  | 110 | 0.2 | 0.60 | 0.61 | 0.26 | 82.48 | 19.15 | 2.47 | 2.72 | 842.77 | 840.28 | 1664.10 |
|  |  | 0.5 | 2.31 | 2.85 | 0.43 | 39.49 | 23.53 | 2.14 | 2.82 | 840.76 | 839.84 | 1525.87 |
|  |  | 0.8 | 4.75 | 6.96 | 0.42 | 26.21 | 17.65 | 2.19 | 3.03 | 840.98 | 840.70 | 1418.54 |
| OR 2.5-3 | 10 | 0.2 | 0.28 | 0.38 | 0.01 | 13.48 | 1.34 | 1.51 | 0.77 | 92.85 | 92.63 | 189.22 |
|  |  | 0.5 | 0.71 | 0.85 | 0.01 | 8.38 | 3.54 | 1.99 | 1.37 | 100.85 | 100.52 | 170.77 |
|  |  | 0.8 | 1.45 | 1.30 | 0.01 | 7.72 | 5.54 | 1.85 | 1.50 | 104.01 | 103.67 | 149.79 |
|  | 30 | 0.2 | 0.39 | 0.48 | 0.02 | 96.83 | 4.19 | 2.00 | 1.14 | 247.08 | 245.59 | 520.49 |
|  |  | 0.5 | 1.85 | 1.75 | 0.04 | 28.92 | 4.33 | 2.16 | 1.69 | 256.30 | 254.18 | 476.39 |
|  |  | 0.8 | 3.29 | 3.40 | 0.06 | 19.28 | 5.74 | 2.23 | 1.95 | 252.41 | 252.40 | 416.91 |
|  | 50 | 0.2 | 0.44 | 0.52 | 0.06 | 158.55 | 7.76 | 2.24 | 1.60 | 403.14 | 397.83 | 819.26 |
|  |  | 0.5 | 1.73 | 1.94 | 0.10 | 28.52 | 7.63 | 2.14 | 1.98 | 415.44 | 413.71 | 782.52 |
|  |  | 0.8 | 3.98 | 5.18 | 0.14 | 19.31 | 7.36 | 2.20 | 2.34 | 412.16 | 427.62 | 706.66 |
|  | 70 | 0.2 | 0.47 | 0.54 | 0.11 | 142.33 | 12.66 | 2.46 | 2.03 | 579.32 | 576.48 | 1188.64 |
|  |  | 0.5 | 3.48 | 3.99 | 0.37 | 56.81 | 18.94 | 2.88 | 3.15 | 890.08 | 888.39 | 1644.10 |
|  |  | 0.8 | 8.04 | 11.15 | 0.36 | 59.62 | 16.64 | 3.12 | 3.65 | 883.90 | 879.40 | 1436.30 |
|  | 90 | 0.2 | 1.02 | 1.09 | 0.35 | 164.93 | 26.69 | 3.37 | 3.28 | 1120.55 | 1114.75 | 2229.67 |
|  |  | 0.5 | 4.06 | 5.19 | 0.61 | 53.85 | 29.52 | 2.95 | 3.46 | 1127.95 | 1124.85 | 2007.56 |
|  |  | 0.8 | 9.86 | 13.85 | 0.59 | 43.05 | 22.88 | 3.00 | 3.86 | 1134.40 | 1114.28 | 1818.57 |
|  | 110 | 0.2 | 1.08 | 1.14 | 0.49 | 152.37 | 33.51 | 3.39 | 3.71 | 1383.01 | 1361.52 | 2642.23 |
|  |  | 0.5 | 5.46 | 7.00 | 0.94 | 56.50 | 37.59 | 2.85 | 3.82 | 1359.51 | 1367.08 | 2404.77 |
|  |  | 0.8 | 9.46 | 12.31 | 0.61 | 30.06 | 24.36 | 2.54 | 3.55 | 1104.28 | 1021.66 | 1630.70 |

MRSL^1^, MRSL when adjusting for all nodes on the open paths; MRSL^2^, MRSL when adjusting for minimum separated set; MRSL^3^, MRSL when adjusting for V\{
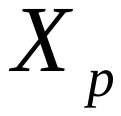
,
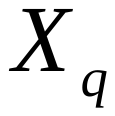
,
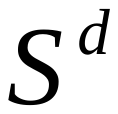
 and U}. MRPC^1^, MRPC algorithm based on the most significant SNP; MRPC^2^, MRPC algorithm based on genetic risk score. HC^1^, HC algorithm incorporating genetic anchors based on the most significant SNP; HC^2^, HC algorithm incorporating genetic anchors based on genetic risk score. cGAUGE^1^, cGAUGE based on IVW; cGAUGE^2^, cGAUGE based on MR Egger; cGAUGE^3^, cGAUGE based on MR PRESSO.

### Supplementary Table 3. Computing time with network of 5 continuous nodes in simulation study 2 (seconds).

| edge effect | g | prob | MRSL^1^ | MRSL^2^ | MRSL^3^ | MRPC^1^ | MRPC^2^ | HC^1^ | HC^2^ | BIMMER | cGAUGE^1^ | cGAUGE^2^ | cGAUGE^3^ |
| --- | --- | --- | --- | --- | --- | --- | --- | --- | --- | --- | --- | --- | --- |
| 0-0.25 | 5 | 0.2 | 0.07 | 0.05 | 0.00 | 0.37 | 0.07 | 0.12 | 0.08 | 3.31 | 7.76 | 8.62 | 20.07 |
|  |  | 0.5 | 0.14 | 0.15 | 0.00 | 0.64 | 0.12 | 0.23 | 0.15 | 5.94 | 11.67 | 11.16 | 25.69 |
|  |  | 0.8 | 0.19 | 0.26 | 0.00 | 0.72 | 0.13 | 0.31 | 0.18 | 6.14 | 12.27 | 11.96 | 26.36 |
|  | 10 | 0.2 | 0.11 | 0.10 | 0.00 | 0.83 | 0.11 | 0.23 | 0.13 | 5.42 | 21.15 | 19.60 | 64.11 |
|  |  | 0.5 | 0.16 | 0.17 | 0.00 | 0.88 | 0.12 | 0.27 | 0.15 | 5.24 | 22.67 | 22.15 | 68.37 |
|  |  | 0.8 | 0.21 | 0.27 | 0.00 | 0.85 | 0.12 | 0.32 | 0.18 | 5.36 | 21.78 | 22.25 | 66.65 |
|  | 20 | 0.2 | 0.09 | 0.09 | 0.00 | 0.81 | 0.11 | 0.27 | 0.15 | 4.63 | 41.99 | 38.01 | 121.43 |
|  |  | 0.5 | 0.20 | 0.19 | 0.00 | 1.13 | 0.16 | 0.34 | 0.17 | 4.87 | 42.20 | 40.19 | 122.75 |
|  |  | 0.8 | 0.19 | 0.25 | 0.00 | 1.12 | 0.11 | 0.38 | 0.19 | 4.50 | 41.88 | 40.19 | 123.28 |
|  | 30 | 0.2 | 0.10 | 0.09 | 0.00 | 1.01 | 0.10 | 0.23 | 0.12 | 3.92 | 56.20 | 51.83 | 129.59 |
|  |  | 0.5 | 0.15 | 0.16 | 0.00 | 1.06 | 0.11 | 0.29 | 0.15 | 5.68 | 58.93 | 58.42 | 153.99 |
|  |  | 0.8 | 0.20 | 0.26 | 0.01 | 1.03 | 0.12 | 0.35 | 0.19 | 4.52 | 60.21 | 58.93 | 152.31 |
|  | 40 | 0.2 | 0.09 | 0.21 | 0.00 | 0.77 | 0.11 | 0.26 | 0.14 | 3.96 | 70.68 | 67.77 | 181.88 |
|  |  | 0.5 | 0.13 | 0.15 | 0.00 | 0.84 | 0.12 | 0.31 | 0.17 | 4.11 | 80.07 | 78.36 | 190.54 |
|  |  | 0.8 | 0.18 | 0.23 | 0.00 | 1.04 | 0.11 | 0.37 | 0.18 | 4.49 | 78.04 | 78.17 | 184.60 |
|  | 50 | 0.2 | 0.10 | 0.09 | 0.00 | 0.81 | 0.12 | 0.25 | 0.15 | 4.20 | 95.38 | 95.89 | 204.09 |
|  |  | 0.5 | 0.14 | 0.16 | 0.00 | 0.88 | 0.12 | 0.28 | 0.16 | 4.16 | 95.30 | 95.40 | 204.14 |
|  |  | 0.8 | 0.24 | 0.28 | 0.01 | 0.84 | 0.13 | 0.33 | 0.19 | 4.39 | 96.17 | 95.37 | 200.28 |
| 0.25-0.5 | 5 | 0.2 | 0.11 | 0.11 | 0.00 | 0.55 | 0.13 | 0.19 | 0.11 | 5.20 | 10.52 | 10.22 | 17.64 |
|  |  | 0.5 | 0.21 | 0.30 | 0.00 | 0.57 | 0.16 | 0.26 | 0.19 | 4.92 | 10.83 | 10.48 | 18.18 |
|  |  | 0.8 | 0.38 | 0.57 | 0.00 | 0.61 | 0.18 | 0.33 | 0.28 | 4.66 | 11.23 | 10.92 | 17.10 |
|  | 10 | 0.2 | 0.11 | 0.13 | 0.00 | 0.60 | 0.10 | 0.21 | 0.11 | 4.78 | 19.84 | 19.99 | 59.86 |
|  |  | 0.5 | 0.25 | 0.35 | 0.00 | 0.57 | 0.15 | 0.28 | 0.17 | 5.61 | 20.67 | 20.96 | 56.28 |
|  |  | 0.8 | 0.38 | 0.54 | 0.00 | 0.55 | 0.19 | 0.31 | 0.28 | 5.38 | 20.67 | 20.47 | 54.54 |
|  | 20 | 0.2 | 0.12 | 0.13 | 0.00 | 0.71 | 0.13 | 0.22 | 0.11 | 4.46 | 38.81 | 35.17 | 98.47 |
|  |  | 0.5 | 0.22 | 0.37 | 0.00 | 0.65 | 0.12 | 0.24 | 0.15 | 4.14 | 33.82 | 36.63 | 96.05 |
|  |  | 0.8 | 0.35 | 0.58 | 0.00 | 0.77 | 0.16 | 0.30 | 0.27 | 5.38 | 39.01 | 38.78 | 96.83 |
|  | 30 | 0.2 | 0.12 | 0.13 | 0.00 | 0.69 | 0.08 | 0.16 | 0.09 | 4.46 | 58.80 | 57.24 | 117.06 |
|  |  | 0.5 | 0.22 | 0.28 | 0.00 | 0.76 | 0.15 | 0.27 | 0.17 | 4.81 | 57.99 | 57.65 | 110.07 |
|  |  | 0.8 | 0.36 | 0.51 | 0.00 | 0.64 | 0.18 | 0.29 | 0.28 | 6.11 | 57.99 | 57.46 | 105.00 |
|  | 40 | 0.2 | 0.11 | 0.12 | 0.00 | 0.56 | 0.09 | 0.16 | 0.10 | 4.24 | 76.44 | 75.64 | 142.21 |
|  |  | 0.5 | 0.23 | 0.29 | 0.01 | 0.60 | 0.13 | 0.24 | 0.17 | 4.55 | 76.56 | 75.77 | 134.84 |
|  |  | 0.8 | 0.41 | 0.67 | 0.01 | 1.15 | 0.15 | 0.28 | 0.26 | 5.30 | 77.71 | 77.59 | 131.61 |
|  | 50 | 0.2 | 0.14 | 0.15 | 0.01 | 0.75 | 0.10 | 0.16 | 0.11 | 4.53 | 94.94 | 94.60 | 160.73 |
|  |  | 0.5 | 0.26 | 0.33 | 0.01 | 0.77 | 0.14 | 0.24 | 0.18 | 4.94 | 94.77 | 95.21 | 156.77 |
|  |  | 0.8 | 0.44 | 0.57 | 0.01 | 0.83 | 0.18 | 0.29 | 0.28 | 5.84 | 96.73 | 96.00 | 153.13 |
| 0.5-0.75 | 5 | 0.2 | 0.14 | 0.16 | 0.00 | 0.50 | 0.12 | 0.20 | 0.13 | 5.14 | 10.58 | 10.40 | 18.38 |
|  |  | 0.5 | 0.31 | 0.46 | 0.00 | 0.52 | 0.19 | 0.26 | 0.22 | 4.89 | 11.07 | 10.71 | 15.55 |
|  |  | 0.8 | 0.47 | 0.69 | 0.00 | 0.49 | 0.19 | 0.25 | 0.32 | 4.53 | 11.22 | 11.25 | 14.36 |
|  | 10 | 0.2 | 0.14 | 0.16 | 0.00 | 0.56 | 0.09 | 0.17 | 0.10 | 4.23 | 17.89 | 17.81 | 39.85 |
|  |  | 0.5 | 0.28 | 0.45 | 0.00 | 0.51 | 0.13 | 0.21 | 0.17 | 4.33 | 16.18 | 17.12 | 41.77 |
|  |  | 0.8 | 0.50 | 0.76 | 0.00 | 0.60 | 0.21 | 0.28 | 0.34 | 5.90 | 22.90 | 23.12 | 43.92 |
|  | 20 | 0.2 | 0.14 | 0.17 | 0.00 | 0.69 | 0.11 | 0.21 | 0.12 | 4.91 | 40.71 | 39.65 | 95.23 |
|  |  | 0.5 | 0.31 | 0.50 | 0.00 | 0.73 | 0.17 | 0.26 | 0.21 | 5.33 | 41.59 | 41.06 | 90.25 |
|  |  | 0.8 | 0.48 | 0.70 | 0.00 | 0.53 | 0.18 | 0.26 | 0.31 | 5.19 | 41.83 | 40.48 | 80.84 |
|  | 30 | 0.2 | 0.17 | 0.21 | 0.00 | 0.73 | 0.12 | 0.22 | 0.14 | 5.13 | 61.33 | 60.14 | 118.54 |
|  |  | 0.5 | 0.34 | 0.47 | 0.01 | 0.59 | 0.17 | 0.25 | 0.22 | 5.13 | 60.80 | 59.35 | 108.93 |
|  |  | 0.8 | 0.59 | 0.80 | 0.01 | 0.54 | 0.18 | 0.25 | 0.32 | 5.17 | 60.61 | 59.59 | 99.13 |
|  | 40 | 0.2 | 0.14 | 0.16 | 0.00 | 1.89 | 0.10 | 0.17 | 0.11 | 4.19 | 76.58 | 73.12 | 143.46 |
|  |  | 0.5 | 0.40 | 0.51 | 0.01 | 1.37 | 0.18 | 0.26 | 0.23 | 5.45 | 80.72 | 78.96 | 117.79 |
|  |  | 0.8 | 0.82 | 1.13 | 0.01 | 1.47 | 0.21 | 0.27 | 0.35 | 6.17 | 79.68 | 77.39 | 126.45 |
|  | 50 | 0.2 | 0.14 | 0.17 | 0.01 | 0.64 | 0.14 | 0.18 | 0.12 | 5.03 | 97.04 | 89.79 | 147.11 |
|  |  | 0.5 | 0.40 | 0.50 | 0.01 | 0.65 | 0.20 | 0.25 | 0.22 | 5.45 | 97.90 | 97.18 | 159.58 |
|  |  | 0.8 | 0.91 | 1.12 | 0.01 | 0.59 | 0.23 | 0.25 | 0.33 | 5.86 | 100.29 | 97.54 | 153.60 |
| 0.75-1 | 5 | 0.2 | 0.12 | 0.14 | 0.00 | 0.49 | 0.11 | 0.17 | 0.11 | 5.09 | 10.14 | 9.34 | 14.21 |
|  |  | 0.5 | 0.22 | 0.31 | 0.00 | 0.36 | 0.12 | 0.18 | 0.18 | 3.42 | 9.59 | 9.10 | 11.75 |
|  |  | 0.8 | 0.53 | 0.62 | 0.00 | 0.42 | 0.15 | 0.20 | 0.26 | 3.53 | 10.01 | 9.55 | 11.12 |
|  | 10 | 0.2 | 0.14 | 0.17 | 0.00 | 0.50 | 0.12 | 0.20 | 0.12 | 4.85 | 21.09 | 20.62 | 53.80 |
|  |  | 0.5 | 0.28 | 0.38 | 0.00 | 0.53 | 0.20 | 0.24 | 0.22 | 4.82 | 20.96 | 20.94 | 41.33 |
|  |  | 0.8 | 0.59 | 0.89 | 0.00 | 0.47 | 0.19 | 0.21 | 0.30 | 4.38 | 21.45 | 21.20 | 34.69 |
|  | 20 | 0.2 | 0.20 | 0.23 | 0.01 | 0.76 | 0.11 | 0.22 | 0.13 | 6.28 | 45.56 | 42.37 | 78.88 |
|  |  | 0.5 | 0.38 | 0.56 | 0.00 | 0.63 | 0.16 | 0.23 | 0.21 | 5.40 | 41.63 | 40.99 | 70.59 |
|  |  | 0.8 | 0.96 | 1.26 | 0.01 | 0.55 | 0.20 | 0.21 | 0.29 | 4.93 | 41.54 | 39.26 | 58.86 |
|  | 30 | 0.2 | 0.14 | 0.16 | 0.00 | 0.75 | 0.10 | 0.17 | 0.12 | 5.20 | 60.08 | 60.31 | 119.79 |
|  |  | 0.5 | 0.36 | 0.50 | 0.01 | 0.63 | 0.15 | 0.19 | 0.19 | 4.63 | 57.84 | 61.16 | 94.72 |
|  |  | 0.8 | 0.90 | 1.28 | 0.01 | 0.54 | 0.17 | 0.19 | 0.27 | 4.59 | 58.79 | 58.36 | 84.75 |
|  | 40 | 0.2 | 0.18 | 0.34 | 0.01 | 1.50 | 0.12 | 0.18 | 0.13 | 5.23 | 84.65 | 81.94 | 140.78 |
|  |  | 0.5 | 0.52 | 0.72 | 0.01 | 1.31 | 0.17 | 0.23 | 0.22 | 5.27 | 81.44 | 81.45 | 127.30 |
|  |  | 0.8 | 1.10 | 1.32 | 0.01 | 0.47 | 1.27 | 0.23 | 0.29 | 4.84 | 80.08 | 80.63 | 109.58 |
|  | 50 | 0.2 | 0.13 | 0.15 | 0.00 | 0.60 | 0.10 | 0.14 | 0.10 | 4.02 | 86.30 | 91.06 | 141.97 |
|  |  | 0.5 | 0.50 | 0.66 | 0.01 | 0.58 | 0.19 | 0.21 | 0.23 | 5.08 | 90.40 | 94.33 | 146.76 |
|  |  | 0.8 | 1.07 | 1.31 | 0.01 | 0.39 | 0.17 | 0.17 | 0.26 | 4.15 | 94.83 | 94.91 | 125.10 |

MRSL^1^, MRSL when adjusting for all nodes on the open paths; MRSL^2^, MRSL when adjusting for minimum separated set; MRSL^3^, MRSL when adjusting for V\{
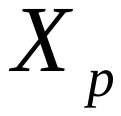
,
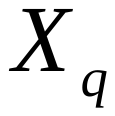
,
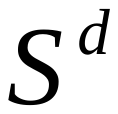
 and U}. MRPC^1^, MRPC algorithm based on the most significant SNP; MRPC^2^, MRPC algorithm based on genetic risk score. HC^1^, HC algorithm incorporating genetic anchors based on the most significant SNP; HC^2^, HC algorithm incorporating genetic anchors based on genetic risk score. cGAUGE^1^, cGAUGE based on IVW; cGAUGE^2^, cGAUGE based on MR Egger; cGAUGE^3^, cGAUGE based on MR PRESSO.

### Supplementary Table 4. Computing time with network of 5 binary nodes in simulation study 2 (seconds).

| edge effect | g | prob | MRSL^1^ | MRSL^2^ | MRSL^3^ | MRPC^1^ | MRPC^2^ | HC^1^ | HC^2^ | cGAUGE^1^ | cGAUGE^2^ | cGAUGE^3^ |
| --- | --- | --- | --- | --- | --- | --- | --- | --- | --- | --- | --- | --- |
| OR 1-1.5 | 10 | 0.2 | 0.04 | 0.03 | 0.00 | 0.40 | 0.05 | 0.12 | 0.06 | 12.37 | 11.70 | 37.54 |
|  |  | 0.5 | 0.06 | 0.04 | 0.00 | 0.50 | 0.07 | 0.20 | 0.10 | 14.22 | 13.09 | 37.84 |
|  |  | 0.8 | 0.06 | 0.04 | 0.00 | 0.59 | 0.07 | 0.24 | 0.14 | 14.53 | 12.33 | 35.63 |
|  | 30 | 0.2 | 0.05 | 0.04 | 0.00 | 0.69 | 0.08 | 0.26 | 0.15 | 36.75 | 33.27 | 104.10 |
|  |  | 0.5 | 0.05 | 0.04 | 0.00 | 0.47 | 0.07 | 0.24 | 0.13 | 34.90 | 32.54 | 100.97 |
|  |  | 0.8 | 0.06 | 0.06 | 0.00 | 0.46 | 0.06 | 0.22 | 0.15 | 35.63 | 33.05 | 101.78 |
|  | 50 | 0.2 | 0.05 | 0.04 | 0.00 | 0.50 | 0.07 | 0.27 | 0.16 | 59.52 | 54.32 | 162.89 |
|  |  | 0.5 | 0.06 | 0.06 | 0.00 | 0.43 | 0.07 | 0.25 | 0.17 | 55.05 | 53.19 | 163.48 |
|  |  | 0.8 | 0.07 | 0.07 | 0.00 | 0.42 | 0.07 | 0.24 | 0.19 | 54.59 | 52.47 | 158.40 |
|  | 70 | 0.2 | 0.05 | 0.05 | 0.00 | 0.45 | 0.07 | 0.24 | 0.19 | 74.27 | 73.04 | 219.72 |
|  |  | 0.5 | 0.06 | 0.06 | 0.00 | 0.44 | 0.07 | 0.24 | 0.20 | 74.03 | 72.94 | 217.48 |
|  |  | 0.8 | 0.08 | 0.08 | 0.00 | 0.42 | 0.07 | 0.24 | 0.21 | 74.67 | 73.40 | 216.92 |
|  | 90 | 0.2 | 0.06 | 0.05 | 0.00 | 0.43 | 0.07 | 0.24 | 0.22 | 95.72 | 94.40 | 273.26 |
|  |  | 0.5 | 0.07 | 0.08 | 0.01 | 0.45 | 0.07 | 0.24 | 0.21 | 97.24 | 94.69 | 270.18 |
|  |  | 0.8 | 0.12 | 0.13 | 0.01 | 0.67 | 0.12 | 0.32 | 0.30 | 141.07 | 135.85 | 379.21 |
|  | 110 | 0.2 | 0.06 | 0.06 | 0.01 | 0.42 | 0.07 | 0.23 | 0.24 | 115.19 | 117.11 | 344.73 |
|  |  | 0.5 | 0.10 | 0.11 | 0.01 | 0.56 | 0.12 | 0.30 | 0.30 | 132.30 | 115.65 | 325.04 |
|  |  | 0.8 | 0.14 | 0.17 | 0.01 | 0.59 | 0.14 | 0.31 | 0.31 | 151.68 | 147.34 | 419.72 |
| OR 1.5-2 | 10 | 0.2 | 0.08 | 0.07 | 0.00 | 0.61 | 0.06 | 0.18 | 0.09 | 18.33 | 17.59 | 55.72 |
|  |  | 0.5 | 0.10 | 0.09 | 0.00 | 0.70 | 0.08 | 0.28 | 0.15 | 19.38 | 18.43 | 55.69 |
|  |  | 0.8 | 0.10 | 0.11 | 0.00 | 0.65 | 0.10 | 0.35 | 0.21 | 18.89 | 18.08 | 53.46 |
|  | 30 | 0.2 | 0.09 | 0.08 | 0.00 | 0.76 | 0.11 | 0.32 | 0.18 | 54.29 | 51.49 | 161.79 |
|  |  | 0.5 | 0.12 | 0.14 | 0.00 | 0.67 | 0.10 | 0.30 | 0.19 | 49.65 | 48.36 | 145.43 |
|  |  | 0.8 | 0.19 | 0.25 | 0.00 | 0.71 | 0.13 | 0.35 | 0.26 | 54.02 | 52.63 | 152.10 |
|  | 50 | 0.2 | 0.11 | 0.12 | 0.00 | 0.76 | 0.13 | 0.38 | 0.24 | 91.02 | 88.70 | 274.20 |
|  |  | 0.5 | 0.14 | 0.17 | 0.00 | 0.64 | 0.12 | 0.28 | 0.22 | 69.43 | 59.44 | 154.00 |
|  |  | 0.8 | 0.15 | 0.18 | 0.00 | 0.45 | 0.10 | 0.23 | 0.20 | 54.32 | 52.52 | 154.52 |
|  | 70 | 0.2 | 0.07 | 0.07 | 0.00 | 0.47 | 0.08 | 0.24 | 0.19 | 74.69 | 72.84 | 219.48 |
|  |  | 0.5 | 0.11 | 0.13 | 0.01 | 0.49 | 0.10 | 0.24 | 0.21 | 74.36 | 72.92 | 215.04 |
|  |  | 0.8 | 0.16 | 0.20 | 0.01 | 0.46 | 0.13 | 0.25 | 0.23 | 74.68 | 72.83 | 210.15 |
|  | 90 | 0.2 | 0.07 | 0.07 | 0.01 | 0.42 | 0.09 | 0.23 | 0.22 | 95.10 | 93.02 | 270.65 |
|  |  | 0.5 | 0.12 | 0.15 | 0.01 | 0.43 | 0.13 | 0.24 | 0.23 | 94.49 | 93.08 | 266.61 |
|  |  | 0.8 | 0.17 | 0.21 | 0.01 | 0.45 | 0.15 | 0.23 | 0.25 | 95.08 | 93.02 | 261.28 |
|  | 110 | 0.2 | 0.08 | 0.09 | 0.01 | 0.45 | 0.09 | 0.25 | 0.24 | 114.74 | 112.38 | 323.67 |
|  |  | 0.5 | 0.13 | 0.15 | 0.01 | 0.45 | 0.13 | 0.24 | 0.24 | 114.94 | 112.46 | 316.55 |
|  |  | 0.8 | 0.19 | 0.25 | 0.01 | 0.44 | 0.17 | 0.23 | 0.25 | 114.73 | 112.32 | 309.46 |
| OR 2-2.5 | 10 | 0.2 | 0.06 | 0.05 | 0.00 | 0.38 | 0.04 | 0.12 | 0.06 | 12.04 | 11.52 | 35.21 |
|  |  | 0.5 | 0.08 | 0.08 | 0.00 | 0.39 | 0.04 | 0.18 | 0.10 | 12.49 | 11.81 | 33.99 |
|  |  | 0.8 | 0.10 | 0.13 | 0.00 | 0.39 | 0.06 | 0.23 | 0.15 | 12.62 | 12.02 | 32.85 |
|  | 30 | 0.2 | 0.07 | 0.07 | 0.00 | 0.47 | 0.06 | 0.22 | 0.12 | 33.78 | 32.49 | 102.06 |
|  |  | 0.5 | 0.11 | 0.13 | 0.00 | 0.45 | 0.07 | 0.21 | 0.14 | 34.02 | 32.58 | 98.14 |
|  |  | 0.8 | 0.16 | 0.21 | 0.00 | 0.45 | 0.08 | 0.23 | 0.18 | 33.75 | 32.38 | 95.12 |
|  | 50 | 0.2 | 0.07 | 0.07 | 0.00 | 0.47 | 0.08 | 0.26 | 0.16 | 54.55 | 52.58 | 161.98 |
|  |  | 0.5 | 0.12 | 0.15 | 0.00 | 0.46 | 0.11 | 0.24 | 0.18 | 54.43 | 52.49 | 156.28 |
|  |  | 0.8 | 0.17 | 0.22 | 0.00 | 0.46 | 0.13 | 0.24 | 0.22 | 54.17 | 52.57 | 151.85 |
|  | 70 | 0.2 | 0.08 | 0.08 | 0.00 | 0.49 | 0.09 | 0.25 | 0.19 | 74.33 | 72.85 | 218.08 |
|  |  | 0.5 | 0.12 | 0.15 | 0.01 | 0.47 | 0.13 | 0.23 | 0.21 | 74.41 | 72.91 | 211.78 |
|  |  | 0.8 | 0.19 | 0.24 | 0.01 | 0.45 | 0.17 | 0.22 | 0.23 | 74.92 | 72.87 | 205.96 |
|  | 90 | 0.2 | 0.08 | 0.08 | 0.01 | 0.44 | 0.10 | 0.24 | 0.22 | 95.51 | 93.14 | 272.50 |
|  |  | 0.5 | 0.14 | 0.17 | 0.01 | 0.44 | 0.15 | 0.23 | 0.23 | 95.54 | 93.24 | 262.27 |
|  |  | 0.8 | 0.20 | 0.25 | 0.01 | 0.44 | 0.20 | 0.24 | 0.25 | 95.44 | 93.35 | 255.26 |
|  | 110 | 0.2 | 0.08 | 0.09 | 0.01 | 0.46 | 0.10 | 0.25 | 0.23 | 114.87 | 112.47 | 320.31 |
|  |  | 0.5 | 0.14 | 0.17 | 0.01 | 0.44 | 0.16 | 0.22 | 0.24 | 115.83 | 112.48 | 312.84 |
|  |  | 0.8 | 0.21 | 0.26 | 0.01 | 0.45 | 0.21 | 0.23 | 0.27 | 116.24 | 112.44 | 305.82 |
| OR 2.5-3 | 10 | 0.2 | 0.07 | 0.07 | 0.00 | 0.38 | 0.04 | 0.13 | 0.06 | 12.23 | 11.57 | 35.98 |
|  |  | 0.5 | 0.10 | 0.10 | 0.00 | 0.43 | 0.06 | 0.19 | 0.11 | 12.53 | 11.86 | 33.70 |
|  |  | 0.8 | 0.14 | 0.18 | 0.00 | 0.48 | 0.08 | 0.24 | 0.17 | 12.26 | 11.94 | 31.97 |
|  | 30 | 0.2 | 0.07 | 0.08 | 0.00 | 0.47 | 0.07 | 0.21 | 0.12 | 33.79 | 32.52 | 100.45 |
|  |  | 0.5 | 0.12 | 0.16 | 0.00 | 0.46 | 0.08 | 0.21 | 0.14 | 34.03 | 32.68 | 96.01 |
|  |  | 0.8 | 0.17 | 0.22 | 0.00 | 0.45 | 0.11 | 0.24 | 0.18 | 33.91 | 32.53 | 92.97 |
|  | 50 | 0.2 | 0.07 | 0.07 | 0.00 | 0.48 | 0.07 | 0.24 | 0.15 | 54.62 | 52.54 | 160.31 |
|  |  | 0.5 | 0.12 | 0.15 | 0.00 | 0.45 | 0.11 | 0.23 | 0.17 | 54.88 | 52.56 | 153.76 |
|  |  | 0.8 | 0.18 | 0.24 | 0.00 | 0.44 | 0.14 | 0.23 | 0.21 | 54.57 | 52.66 | 147.50 |
|  | 70 | 0.2 | 0.08 | 0.08 | 0.00 | 0.47 | 0.09 | 0.25 | 0.19 | 74.28 | 72.30 | 217.10 |
|  |  | 0.5 | 0.13 | 0.16 | 0.01 | 0.44 | 0.13 | 0.23 | 0.21 | 74.88 | 72.33 | 209.29 |
|  |  | 0.8 | 0.20 | 0.25 | 0.01 | 0.44 | 0.17 | 0.23 | 0.23 | 74.68 | 72.91 | 202.08 |
|  | 90 | 0.2 | 0.08 | 0.09 | 0.01 | 0.45 | 0.10 | 0.24 | 0.22 | 94.39 | 93.08 | 270.03 |
|  |  | 0.5 | 0.14 | 0.17 | 0.01 | 0.46 | 0.15 | 0.23 | 0.22 | 94.69 | 93.23 | 260.03 |
|  |  | 0.8 | 0.21 | 0.26 | 0.01 | 0.43 | 0.20 | 0.21 | 0.25 | 94.74 | 93.22 | 252.15 |
|  | 110 | 0.2 | 0.08 | 0.09 | 0.01 | 0.44 | 0.11 | 0.23 | 0.24 | 114.97 | 112.41 | 319.35 |
|  |  | 0.5 | 0.16 | 0.19 | 0.01 | 0.46 | 0.17 | 0.22 | 0.25 | 115.92 | 112.65 | 309.02 |
|  |  | 0.8 | 0.23 | 0.29 | 0.01 | 0.42 | 0.23 | 0.22 | 0.27 | 115.85 | 112.60 | 298.49 |

MRSL^1^, MRSL when adjusting for all nodes on the open paths; MRSL^2^, MRSL when adjusting for minimum separated set; MRSL^3^, MRSL when adjusting for V\{
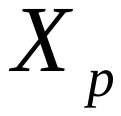
,
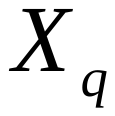
,
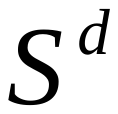
 and U}. MRPC^1^, MRPC algorithm based on the most significant SNP; MRPC^2^, MRPC algorithm based on genetic risk score. HC^1^, HC algorithm incorporating genetic anchors based on the most significant SNP; HC^2^, HC algorithm incorporating genetic anchors based on genetic risk score. cGAUGE^1^, cGAUGE based on IVW; cGAUGE^2^, cGAUGE based on MR Egger; cGAUGE^3^, cGAUGE based on MR PRESSO.

### Supplementary Table 5. Computing time with network of 15 continuous nodes in simulation study 2 (seconds).

| edge effect | g | prob | MRSL^1^ | MRSL^2^ | MRSL^3^ | HC^1^ | HC^2^ | BIMMER |
| --- | --- | --- | --- | --- | --- | --- | --- | --- |
| 0-0.25 | 5 | 0.2 | 0.17 | 0.83 | 0.91 | 19.11 | 1.22 | 0.70 |
|  |  | 0.5 | 0.10 | 0.90 | 0.96 | 50.75 | 2.33 | 1.94 |
|  |  | 0.8 | 0.12 | 0.88 | 0.95 | 77.52 | 3.11 | 5.75 |
|  | 10 | 0.2 | 0.37 | 0.63 | 0.85 | 17.27 | 2.26 | 3.63 |
|  |  | 0.35 | 0.30 | 0.70 | 0.91 | 31.57 | 2.88 | 6.43 |
|  |  | 0.8 | 0.19 | 0.81 | 0.98 | 75.55 | 3.67 | 7.38 |
|  | 20 | 0.2 | 1.00 | 0.89 | 0.05 | 7.93 | 2.53 | 5.37 |
|  |  | 0.5 | 3.10 | 2.41 | 0.08 | 8.32 | 3.68 | 10.16 |
|  |  | 0.8 | 4.55 | 5.34 | 0.10 | 6.92 | 4.66 | 19.08 |
|  | 30 | 0.2 | 0.98 | 0.87 | 0.09 | 7.50 | 3.01 | 5.30 |
|  |  | 0.5 | 3.03 | 2.57 | 0.14 | 7.90 | 4.82 | 13.01 |
|  |  | 0.8 | 4.70 | 6.38 | 0.19 | 6.37 | 6.69 | 23.32 |
|  | 40 | 0.2 | 1.05 | 0.90 | 0.14 | 7.30 | 3.17 | 5.20 |
|  |  | 0.5 | 5.40 | 2.79 | 0.25 | 7.55 | 5.48 | 14.30 |
|  |  | 0.8 | 4.99 | 7.78 | 0.31 | 5.99 | 7.15 | 24.15 |
|  | 50 | 0.2 | 1.04 | 0.90 | 0.22 | 6.48 | 3.26 | 5.90 |
|  |  | 0.5 | 6.55 | 2.98 | 0.39 | 6.89 | 6.15 | 15.58 |
|  |  | 0.8 | 5.35 | 9.27 | 0.49 | 5.84 | 7.69 | 25.32 |
| 0.25-0.5 | 5 | 0.2 | 0.21 | 0.79 | 0.81 | 17.16 | 1.02 | 4.74 |
|  |  | 0.5 | 0.41 | 0.59 | 0.74 | 39.00 | 1.94 | 13.78 |
|  |  | 0.8 | 0.68 | 0.32 | 0.72 | 58.12 | 2.82 | 24.78 |
|  | 10 | 0.2 | 0.24 | 0.76 | 0.98 | 20.35 | 1.78 | 1.57 |
|  |  | 0.5 | 0.31 | 0.69 | 0.93 | 46.00 | 2.28 | 5.55 |
|  |  | 0.8 | 0.64 | 0.36 | 0.78 | 60.58 | 2.92 | 22.31 |
|  | 20 | 0.2 | 4.97 | 1.73 | 0.07 | 6.05 | 2.05 | 9.20 |
|  |  | 0.5 | 18.64 | 20.25 | 0.10 | 4.72 | 2.73 | 6.07 |
|  |  | 0.8 | 12.03 | 28.61 | 0.04 | 3.23 | 3.46 | 3.67 |
|  | 30 | 0.2 | 3.16 | 1.89 | 0.12 | 5.04 | 2.29 | 13.91 |
|  |  | 0.5 | 7.90 | 39.14 | 0.16 | 4.03 | 3.61 | 7.97 |
|  |  | 0.8 | 13.80 | 21.15 | 0.06 | 2.99 | 4.93 | 3.98 |
|  | 40 | 0.2 | 3.77 | 2.06 | 0.21 | 4.73 | 2.71 | 14.93 |
|  |  | 0.5 | 9.33 | 23.66 | 0.23 | 4.17 | 4.09 | 9.43 |
|  |  | 0.8 | 14.93 | 23.50 | 0.07 | 3.01 | 5.59 | 4.34 |
|  | 50 | 0.2 | 5.25 | 2.66 | 0.34 | 4.27 | 2.83 | 17.84 |
|  |  | 0.5 | 11.03 | 23.07 | 0.30 | 4.07 | 4.64 | 9.92 |
|  |  | 0.8 | 16.71 | 25.29 | 0.08 | 2.93 | 5.96 | 4.44 |
| 0.5-0.75 | 5 | 0.2 | 0.39 | 0.61 | 0.57 | 11.94 | 3.19 | 8.82 |
|  |  | 0.5 | 0.71 | 0.29 | 0.55 | 28.32 | 5.55 | 24.08 |
|  |  | 0.8 | 0.84 | 0.16 | 0.50 | 39.90 | 7.21 | 42.73 |
|  | 10 | 0.2 | 0.30 | 0.70 | 0.91 | 18.03 | 3.65 | 2.62 |
|  |  | 0.5 | 0.65 | 0.35 | 0.67 | 32.77 | 6.41 | 19.54 |
|  |  | 0.8 | 0.84 | 0.16 | 0.52 | 41.05 | 6.80 | 42.16 |
|  | 20 | 0.2 | 9.86 | 2.51 | 0.07 | 4.73 | 4.17 | 8.71 |
|  |  | 0.5 | 10.82 | 28.46 | 0.06 | 3.96 | 8.58 | 3.78 |
|  |  | 0.8 | 14.87 | 26.58 | 0.02 | 2.78 | 8.19 | 3.03 |
|  | 30 | 0.2 | 10.64 | 3.37 | 0.14 | 4.39 | 4.93 | 10.49 |
|  |  | 0.5 | 13.40 | 27.54 | 0.07 | 3.83 | 10.01 | 4.08 |
|  |  | 0.8 | 16.17 | 25.81 | 0.03 | 2.74 | 11.51 | 3.07 |
|  | 40 | 0.2 | 11.13 | 4.41 | 0.23 | 4.27 | 6.00 | 13.16 |
|  |  | 0.5 | 14.77 | 22.06 | 0.10 | 3.88 | 10.79 | 4.23 |
|  |  | 0.8 | 17.57 | 27.53 | 0.03 | 2.74 | 9.66 | 3.23 |
|  | 50 | 0.2 | 12.06 | 6.82 | 0.52 | 4.74 | 6.18 | 19.12 |
|  |  | 0.5 | 20.16 | 23.10 | 0.13 | 3.82 | 9.79 | 4.35 |
|  |  | 0.8 | 26.26 | 39.84 | 0.05 | 3.32 | 11.85 | 4.32 |
| 0.75-1 | 5 | 0.2 | 0.65 | 0.35 | 0.35 | 7.24 | 17.97 | 13.46 |
|  |  | 0.5 | 0.84 | 0.16 | 0.34 | 17.27 | 4.07 | 35.40 |
|  |  | 0.8 | 0.92 | 0.08 | 0.29 | 23.74 | 3.02 | 59.57 |
|  | 10 | 0.2 | 0.47 | 0.53 | 0.72 | 14.65 | 9.73 | 6.52 |
|  |  | 0.5 | 0.75 | 0.25 | 0.50 | 24.52 | 4.98 | 27.99 |
|  |  | 0.8 | 7.61 | 14.05 | 0.19 | 15.24 | 1.93 | 27.84 |
|  | 20 | 0.2 | 398.11 | 6.95 | 0.09 | 4.85 | 7.15 | 6.64 |
|  |  | 0.5 | 14.58 | 77.58 | 0.04 | 3.98 | 3.49 | 3.25 |
|  |  | 0.8 | 15.75 | 25.04 | 0.02 | 3.00 | 1.68 | 2.77 |
|  | 30 | 0.2 | 564.77 | 5.11 | 0.14 | 3.99 | 7.39 | 6.42 |
|  |  | 0.5 | 21.81 | 96.78 | 0.08 | 4.72 | 3.10 | 4.24 |
|  |  | 0.8 | 35.77 | 56.04 | 0.04 | 3.55 | 1.78 | 3.49 |
|  | 40 | 0.2 | 905.81 | 17.45 | 0.45 | 7.20 | 7.60 | 15.01 |
|  |  | 0.5 | 32.68 | 156.18 | 0.13 | 6.58 | 2.43 | 6.57 |
|  |  | 0.8 | 37.13 | 58.64 | 0.04 | 5.08 | 1.39 | 5.33 |
|  | 50 | 0.2 | 1433.09 | 58.58 | 0.70 | 6.46 | 5.13 | 16.48 |
|  |  | 0.5 | 33.12 | 246.11 | 0.16 | 6.32 | 2.94 | 6.70 |
|  |  | 0.8 | 36.91 | 58.31 | 0.05 | 4.71 | 1.54 | 5.88 |

MRSL^1^, MRSL when adjusting for all nodes on the open paths; MRSL^2^, MRSL when adjusting for minimum separated set; MRSL^3^, MRSL when adjusting for V\{
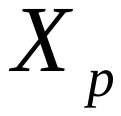
,
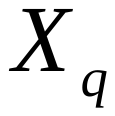
,
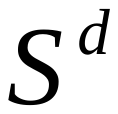
 and U}. HC^1^, HC algorithm incorporating genetic anchors based on the most significant SNP; HC^2^, HC algorithm incorporating genetic anchors based on genetic risk score. MRPC and cGAUGE are not listed due to their huge time consuming.

### Supplementary Table 6. Computing time with network of 15 binary nodes in simulation study 2 (seconds).

| edge effect | g | prob | MRSL^1^ | MRSL^2^ | MRSL^3^ | HC^1^ | HC^2^ |
| --- | --- | --- | --- | --- | --- | --- | --- |
| OR 1-1.5 | 10 | 0.2 | 0.63 | 0.93 | 0.04 | 11.18 | 5.88 |
|  |  | 0.5 | 0.79 | 1.11 | 0.04 | 14.91 | 9.63 |
|  |  | 0.8 | 1.01 | 1.28 | 0.04 | 14.90 | 10.34 |
|  | 30 | 0.2 | 0.86 | 1.00 | 0.20 | 14.01 | 7.96 |
|  |  | 0.5 | 1.66 | 1.62 | 0.26 | 16.01 | 10.54 |
|  |  | 0.8 | 2.62 | 2.31 | 0.33 | 16.68 | 11.52 |
|  | 50 | 0.2 | 0.90 | 1.12 | 0.49 | 15.13 | 10.40 |
|  |  | 0.5 | 2.76 | 2.11 | 0.70 | 16.60 | 12.53 |
|  |  | 0.8 | 4.73 | 3.06 | 0.83 | 15.86 | 12.90 |
|  | 70 | 0.2 | 1.55 | 1.53 | 1.19 | 15.70 | 13.02 |
|  |  | 0.5 | 3.36 | 2.78 | 1.49 | 16.61 | 14.90 |
|  |  | 0.8 | 7.69 | 5.83 | 1.95 | 16.13 | 15.44 |
|  | 90 | 0.2 | 1.80 | 1.79 | 2.08 | 15.97 | 15.19 |
|  |  | 0.5 | 5.98 | 3.44 | 2.55 | 15.70 | 16.35 |
|  |  | 0.8 | 15.24 | 7.89 | 3.24 | 16.27 | 17.46 |
|  | 110 | 0.2 | 2.36 | 2.09 | 2.95 | 15.42 | 16.47 |
|  |  | 0.5 | 6.53 | 5.32 | 4.13 | 15.54 | 18.29 |
|  |  | 0.8 | 9.52 | 11.13 | 4.77 | 15.70 | 18.99 |
| OR 1.5-2 | 10 | 0.2 | 1.11 | 1.47 | 0.05 | 11.91 | 6.69 |
|  |  | 0.5 | 2.39 | 2.25 | 0.06 | 12.63 | 8.79 |
|  |  | 0.8 | 4.11 | 3.23 | 0.06 | 12.05 | 9.53 |
|  | 30 | 0.2 | 2.68 | 2.27 | 0.31 | 13.71 | 7.99 |
|  |  | 0.5 | 7.93 | 7.09 | 0.49 | 15.28 | 10.52 |
|  |  | 0.8 | 20.12 | 14.43 | 0.76 | 13.93 | 11.12 |
|  | 50 | 0.2 | 3.45 | 3.01 | 0.86 | 14.52 | 10.06 |
|  |  | 0.5 | 10.89 | 11.15 | 1.45 | 14.20 | 11.93 |
|  |  | 0.8 | 15.16 | 28.04 | 2.06 | 14.09 | 12.58 |
|  | 70 | 0.2 | 4.32 | 3.78 | 1.76 | 14.52 | 12.34 |
|  |  | 0.5 | 22.28 | 17.82 | 3.04 | 13.70 | 13.22 |
|  |  | 0.8 | 22.84 | 48.81 | 4.30 | 12.91 | 13.08 |
|  | 90 | 0.2 | 4.18 | 4.31 | 2.98 | 13.82 | 14.30 |
|  |  | 0.5 | 12.23 | 27.04 | 5.08 | 13.55 | 15.20 |
|  |  | 0.8 | 30.78 | 56.72 | 5.73 | 13.05 | 14.81 |
|  | 110 | 0.2 | 6.05 | 5.99 | 4.47 | 14.28 | 16.05 |
|  |  | 0.5 | 233.05 | 30.45 | 8.11 | 12.56 | 15.21 |
|  |  | 0.8 | 44.24 | 76.69 | 7.86 | 13.35 | 17.02 |
| OR 2-2.5 | 10 | 0.2 | 1.98 | 2.22 | 0.05 | 11.05 | 6.36 |
|  |  | 0.5 | 4.76 | 3.59 | 0.07 | 11.19 | 8.34 |
|  |  | 0.8 | 9.72 | 5.25 | 0.08 | 9.59 | 7.75 |
|  | 30 | 0.2 | 4.40 | 3.55 | 0.36 | 12.11 | 7.38 |
|  |  | 0.5 | 20.22 | 12.27 | 0.65 | 12.57 | 9.57 |
|  |  | 0.8 | 22.76 | 27.92 | 0.96 | 11.38 | 9.53 |
|  | 50 | 0.2 | 4.91 | 4.08 | 1.02 | 13.39 | 9.79 |
|  |  | 0.5 | 12.42 | 21.41 | 1.78 | 12.17 | 10.73 |
|  |  | 0.8 | 28.83 | 48.78 | 2.49 | 11.00 | 10.42 |
|  | 70 | 0.2 | 6.84 | 5.22 | 2.24 | 12.92 | 11.65 |
|  |  | 0.5 | 50.89 | 30.63 | 3.71 | 11.74 | 12.35 |
|  |  | 0.8 | 38.05 | 79.61 | 4.12 | 11.44 | 12.59 |
|  | 90 | 0.2 | 9.08 | 6.42 | 3.37 | 13.60 | 13.70 |
|  |  | 0.5 | 21.87 | 39.90 | 6.80 | 11.90 | 14.10 |
|  |  | 0.8 | 49.61 | 75.93 | 4.97 | 11.50 | 13.99 |
|  | 110 | 0.2 | 8.25 | 9.77 | 5.75 | 13.37 | 15.13 |
|  |  | 0.5 | 27.96 | 47.61 | 9.47 | 11.68 | 15.05 |
|  |  | 0.8 | 62.09 | 207.54 | 5.55 | 10.92 | 14.53 |
| OR 2.5-3 | 10 | 0.2 | 2.60 | 2.61 | 0.06 | 10.12 | 5.97 |
|  |  | 0.5 | 7.17 | 4.72 | 0.08 | 9.71 | 7.70 |
|  |  | 0.8 | 15.07 | 6.77 | 0.10 | 8.01 | 7.18 |
|  | 30 | 0.2 | 5.23 | 4.32 | 0.40 | 12.36 | 7.56 |
|  |  | 0.5 | 18.10 | 15.85 | 0.74 | 11.48 | 9.20 |
|  |  | 0.8 | 32.43 | 38.08 | 1.08 | 9.37 | 8.40 |
|  | 50 | 0.2 | 9.25 | 5.25 | 1.03 | 12.37 | 9.20 |
|  |  | 0.5 | 120.64 | 25.00 | 2.02 | 11.30 | 10.21 |
|  |  | 0.8 | 39.76 | 155.11 | 2.42 | 9.55 | 9.60 |
|  | 70 | 0.2 | 15.97 | 6.87 | 2.27 | 12.59 | 11.12 |
|  |  | 0.5 | 107.27 | 52.24 | 4.29 | 11.32 | 11.93 |
|  |  | 0.8 | 38.05 | 79.61 | 4.12 | 9.94 | 11.50 |
|  | 90 | 0.2 | 18.68 | 8.27 | 4.07 | 12.94 | 12.81 |
|  |  | 0.5 | 27.58 | 46.18 | 6.74 | 10.67 | 12.65 |
|  |  | 0.8 | 57.47 | 206.58 | 3.39 | 9.65 | 12.03 |
|  | 110 | 0.2 | 24.67 | 10.34 | 5.71 | 12.40 | 14.49 |
|  |  | 0.5 | 35.71 | 60.29 | 8.44 | 10.65 | 14.26 |
|  |  | 0.8 | 71.87 | 121.28 | 3.70 | 9.66 | 13.42 |

MRSL^1^, MRSL when adjusting for all nodes on the open paths; MRSL^2^, MRSL when adjusting for minimum separated set; MRSL^3^, MRSL when adjusting for V\{
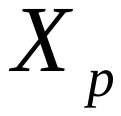
,
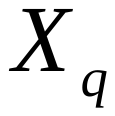
,
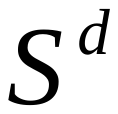
 and U}. HC^1^, HC algorithm incorporating genetic anchors based on the most significant SNP; HC^2^, HC algorithm incorporating genetic anchors based on genetic risk score. MRPC and cGAUGE are not listed due to their huge time consuming.

### Supplementary Table 7. 44 diseases and 26 biomarkers in the applied example.

| Description | Variable type | Source | N_non_missing | N_missing | N_controls | N_cases |
| --- | --- | --- | --- | --- | --- | --- |
| Alanine aminotransferase (quantile) | continuous_irnt | biomarkers | 344136 | 17003 | NA | NA |
| Albumin (quantile) | continuous_irnt | biomarkers | 315268 | 45871 | NA | NA |
| Alkaline phosphatase (quantile) | continuous_irnt | biomarkers | 344292 | 16847 | NA | NA |
| Apoliprotein A (quantile) | continuous_irnt | biomarkers | 313387 | 47752 | NA | NA |
| Apoliprotein B (quantile) | continuous_irnt | biomarkers | 342590 | 18549 | NA | NA |
| Aspartate aminotransferase (quantile) | continuous_irnt | biomarkers | 342990 | 18149 | NA | NA |
| C-reactive protein (quantile) | continuous_irnt | biomarkers | 343524 | 17615 | NA | NA |
| Calcium (quantile) | continuous_irnt | biomarkers | 315153 | 45986 | NA | NA |
| Cholesterol (quantile) | continuous_irnt | biomarkers | 344278 | 16861 | NA | NA |
| Creatinine (quantile) | continuous_irnt | biomarkers | 344104 | 17035 | NA | NA |
| Cystatin C (quantile) | continuous_irnt | biomarkers | 344264 | 16875 | NA | NA |
| Gamma glutamyltransferase (quantile) | continuous_irnt | biomarkers | 344104 | 17035 | NA | NA |
| Glucose (quantile) | continuous_irnt | biomarkers | 314916 | 46223 | NA | NA |
| Glycated haemoglobin (quantile) | continuous_irnt | biomarkers | 344182 | 16957 | NA | NA |
| HDL cholesterol (quantile) | continuous_irnt | biomarkers | 315133 | 46006 | NA | NA |
| IGF-1 (quantile) | continuous_irnt | biomarkers | 342439 | 18700 | NA | NA |
| LDL direct (quantile) | continuous_irnt | biomarkers | 343621 | 17518 | NA | NA |
| Phosphate (quantile) | continuous_irnt | biomarkers | 314658 | 46481 | NA | NA |
| SHBG (quantile) | continuous_irnt | biomarkers | 312215 | 48924 | NA | NA |
| Testosterone (quantile) | continuous_irnt | biomarkers | 312102 | 49037 | NA | NA |
| Total bilirubin (quantile) | continuous_irnt | biomarkers | 342829 | 18310 | NA | NA |
| Total protein (quantile) | continuous_irnt | biomarkers | 314921 | 46218 | NA | NA |
| Triglycerides (quantile) | continuous_irnt | biomarkers | 343992 | 17147 | NA | NA |
| Urate (quantile) | continuous_irnt | biomarkers | 343836 | 17303 | NA | NA |
| Urea (quantile) | continuous_irnt | biomarkers | 344052 | 17087 | NA | NA |
| Vitamin D (quantile) | continuous_irnt | biomarkers | 329247 | 31892 | NA | NA |
| B37 Candidiasis | binary | icd10 | 361194 | 0 | 360942 | 252 |
| C16 Malignant neoplasm of stomach | binary | icd10 | 361194 | 0 | 360806 | 388 |
| D04 Carcinoma in situ of skin | binary | icd10 | 361194 | 0 | 360792 | 402 |
| D11 Benign neoplasm of major salivary glands | binary | icd10 | 361194 | 0 | 360818 | 376 |
| F31 Bipolar affective disorder | binary | icd10 | 361194 | 0 | 360823 | 371 |
| F33 Recurrent depressive disorder | binary | icd10 | 361194 | 0 | 360901 | 293 |
| F43 Reaction to severe stress, and adjustment disorders | binary | icd10 | 361194 | 0 | 360967 | 227 |
| G81 Hemiplegia | binary | icd10 | 361194 | 0 | 360903 | 291 |
| H60 Otitis externa | binary | icd10 | 361194 | 0 | 360892 | 302 |
| H81 Disorders of vestibular function | binary | icd10 | 361194 | 0 | 360853 | 341 |
| H92 Otalgia and effusion of ear | binary | icd10 | 361194 | 0 | 360943 | 251 |
| I25 Chronic ischaemic heart disease | binary | icd10 | 361194 | 0 | 348425 | 12769 |
| I48 Atrial fibrillation and flutter | binary | icd10 | 361194 | 0 | 354838 | 6356 |
| J03 Acute tonsillitis | binary | icd10 | 361194 | 0 | 360789 | 405 |
| J31 Chronic rhinitis, nasopharyngitis and pharyngitis | binary | icd10 | 361194 | 0 | 360903 | 291 |
| K07 Dentofacial anomalies [including malocclusion] | binary | icd10 | 361194 | 0 | 360784 | 410 |
| K12 Stomatitis and related lesions | binary | icd10 | 361194 | 0 | 360923 | 271 |
| K40 Inguinal hernia | binary | icd10 | 361194 | 0 | 348047 | 13147 |
| K41 Femoral hernia | binary | icd10 | 361194 | 0 | 360728 | 466 |
| K74 Fibrosis and cirrhosis of liver | binary | icd10 | 361194 | 0 | 360942 | 252 |
| K90 Intestinal malabsorption | binary | icd10 | 361194 | 0 | 360272 | 922 |
| L40 Psoriasis | binary | icd10 | 361194 | 0 | 360720 | 474 |
| L43 Lichen planus | binary | icd10 | 361194 | 0 | 360820 | 374 |
| M22 Disorders of patella | binary | icd10 | 361194 | 0 | 360792 | 402 |
| M72 Fibroblastic disorders | binary | icd10 | 361194 | 0 | 358001 | 3193 |
| M76 Enthesopathies of lower limb, excluding foot | binary | icd10 | 361194 | 0 | 360905 | 289 |
| M81 Osteoporosis without pathological fracture | binary | icd10 | 361194 | 0 | 360457 | 737 |
| N62 Hypertrophy of breast | binary | icd10 | 361194 | 0 | 360645 | 549 |
| R14 Flatulence and related conditions | binary | icd10 | 361194 | 0 | 360863 | 331 |
| R15 Faecal incontinence | binary | icd10 | 361194 | 0 | 360284 | 910 |
| R18 Ascites | binary | icd10 | 361194 | 0 | 360835 | 359 |
| R25 Abnormal involuntary movements | binary | icd10 | 361194 | 0 | 360961 | 233 |
| R26 Abnormalities of gait and mobility | binary | icd10 | 361194 | 0 | 360864 | 330 |
| R30 Pain associated with micturition | binary | icd10 | 361194 | 0 | 360835 | 359 |
| S30 Superficial injury of abdomen, lower back and pelvis | binary | icd10 | 361194 | 0 | 360948 | 246 |
| S64 Injury of nerves at wrist and hand level | binary | icd10 | 361194 | 0 | 360822 | 372 |
| S80 Superficial injury of lower leg | binary | icd10 | 361194 | 0 | 360963 | 231 |
| S81 Open wound of lower leg | binary | icd10 | 361194 | 0 | 360907 | 287 |
| T17 Foreign body in respiratory tract | binary | icd10 | 361194 | 0 | 360957 | 237 |
| Z11 Special screening examination for infectious and parasitic diseases | binary | icd10 | 361194 | 0 | 360943 | 251 |
| Z50 Care involving use of rehabilitation procedures | binary | icd10 | 361194 | 0 | 360976 | 218 |
| Z52 Donors of organs and tissues | binary | icd10 | 361194 | 0 | 360917 | 277 |
| Z80 Family history of malignant neoplasm | binary | icd10 | 361194 | 0 | 360817 | 377 |
| Z85 Personal history of malignant neoplasm | binary | icd10 | 361194 | 0 | 360956 | 238 |
